# Modeling geographic vaccination strategies for COVID-19 in Norway

**DOI:** 10.1101/2023.08.16.23294112

**Authors:** Louis Yat Hin Chan, Gunnar Rø, Jørgen Eriksson Midtbø, Francesco Di Ruscio, Sara Sofie Viksmoen Watle, Lene Kristine Juvet, Jasper Littmann, Preben Aavitsland, Karin Maria Nygård, Are Stuwitz Berg, Geir Bukholm, Anja Bråthen Kristoffersen, Kenth Engø-Monsen, Solveig Engebretsen, David Swanson, Alfonso Diz-Lois Palomares, Jonas Christoffer Lindstrøm, Arnoldo Frigessi, Birgitte Freiesleben de Blasio

## Abstract

1

Vaccination was a key intervention in controlling the COVID-19 pandemic globally. In early 2021, Norway faced significant regional variations in COVID-19 incidence and prevalence, with large differences in population density, necessitating efficient vaccine allocation to reduce infections and severe outcomes. This study explored alternative vaccination strategies to minimize health outcomes (infections, hospitalizations, ICU admissions, deaths) by varying regions prioritized, extra doses prioritized, and implementation start time.

Using two models (individual-based and meta-population), we simulated COVID-19 transmission during the primary vaccination period in Norway, covering the first 7 months of 2021. We investigated alternative strategies to allocate more vaccine doses to regions with a higher force of infection. We also examined the robustness of our results and highlighted potential structural differences between the two models.

Our findings suggest that early vaccine prioritization could reduce COVID-19 related health outcomes by 8% to 20% compared to a baseline strategy without geographic prioritization. For minimizing infections, hospitalizations, or ICU admissions, the best strategy was to initially allocate all available vaccine doses to fewer high-risk municipalities, comprising approximately one-fourth of the population. For minimizing deaths, a moderate level of geographic prioritization, with approximately one-third of the population receiving doubled doses, gave the best outcomes by balancing the trade-off between vaccinating younger people in high-risk areas and older people in low-risk areas.

The actual strategy implemented in Norway was a two-step moderate level aimed at maintaining the balance and ensuring ethical considerations and public trust. However, it did not offer significant advantages over the baseline strategy without geographic prioritization. Earlier implementation of geographic prioritization could have more effectively addressed the main wave of infections, substantially reducing the national burden of the pandemic.

**Author summary:** We utilized two geographic-age-structured models (an individual-based model and a meta-population model) to conduct a scenario-based analysis aimed at evaluating strategies for geographic prioritization of COVID-19 vaccines in Norway. By reconstructing the dynamics of COVID-19 transmission from January to July of 2021, we compared various alternative vaccination strategies through model simulations, given the limited number of vaccine doses. We found that prioritization of vaccines based on geographic location, alongside considering age, was preferable to a baseline strategy without geographic prioritization. We assessed the selection of which municipalities to prioritize and the degree of prioritization they should receive. Our findings indicated that optimal strategies depended on whether the aim was to minimize infections, hospitalizations, ICU admissions, or deaths. Trade-offs in infection growth between municipalities and subsequent risk-class allocations (such as age groups) were the primary factors influencing optimal vaccine allocation. Furthermore, we found that earlier implementation of most geographic prioritization strategies was advantageous in reducing the overall burden of COVID-19.

## 3 Introduction

The COVID-19 pandemic has placed an enormous burden on public health systems globally. Since the first detection of the SARS-CoV-2 virus in Wuhan, China, in late December 2019 [1], it rapidly spread to other countries and triggered a worldwide health crisis [2]. The first imported case in Norway was confirmed on the 26th of February 2020 [3]. On the 12th of March 2020, the Norwegian government implemented a national lockdown that involved closure of schools and non-essential businesses, border controls, travel restrictions and social distancing measures [4–6]. The measures were gradually scaled back, and from the autumn of 2020, the government shifted its policy to regional differentiation based on local transmission levels [7, 8]. Until the start of 2021, the government maintained a strict mitigation strategy to avoid overloading the healthcare system, prevent deaths, and focus on early testing and isolation of cases, along with a locally-based contact tracing policy.

At the turn of 2020/2021, in the face of the emergence of the Alpha variant and limited global and domestic vaccine production and supply, Norway launched a mass COVID-19 vaccination program [9], mainly distributing the two messenger RNA (mRNA) vaccines, Comirnaty (Pfizer-BioNTech) and Spikevax (Moderna), to the population. The government initially prioritized vaccines to protect individuals most at risk for severe illness, vaccinating those aged 65 years and above, along with people with underlying medical conditions [10–12]. Additionally, healthcare workers were given priority due to their high exposure and risk of transmitting the virus to vulnerable patients.

Due to substantial variations in infection levels across the country, with urban areas, notably in and around the capital city of Oslo, experiencing particularly high infection rates, the Norwegian government decided to implement a geographic prioritization of vaccines. Initially, on the 9th of March 2021, a moderate geographic prioritization of vaccines was introduced in Oslo and several neighboring municipalities [13]. Subsequently, on the 19th of May 2021, a more significant redistribution was implemented, which both increased the amount of the prioritization and broadened the geographic scope of vaccine distribution [14]. By the 31st of July 2021, around 3.6 million (68% of the total population) and 1.8 million (34% of the total population) had received their first and second doses, respectively [15].

Selecting a strategy for distributing a vaccine in limited supply during an ongoing pandemic is a complicated task. Particularly, there is a trade-off between direct protection of vulnerable groups versus indirect protection and overall reduction of transmission. Additionally, effective vaccination strategies must account for local COVID-19 epidemiology. Geographic prioritization of COVID-19 vaccines can be guided by several factors, including infection rates, hospitalization rates, and mortality rates, while also accounting for the ability to control transmission over time without resorting to lockdown or other measures with significant societal consequences and costs. For this reason, the optimal strategy may vary depending on the objective and aim of the vaccination program.

Some modeling studies have explored vaccination strategies, focusing on age, health condition and occupation as the primary targets considered for prioritization [16–27]. Most studies concluded that prioritizing high-risk individuals such as the elderly, those with underlying diseases and healthcare workers is preferred for minimizing deaths. On the other hand, prioritizing younger people who have higher contact rates is better for reducing transmission and hence infections. A few modeling studies have investigated geographic allocation of vaccines taking into account spatial heterogeneities as the secondary consideration on top of age [28–31]. Bertsimas et al. optimized the location of vaccination sites to minimize deaths in the US [28]. Lemaitre et al. and Molla et al. optimized by age and region simultaneously based on optimal control theory and showed that vaccine allocation to high incidence areas is the optimal strategy to reduce deaths in Italy and Finland, respectively [29, 30]. Grauer et al. found that deaths could be significantly reduced if vaccines were distributed focusing on one region at a time, using a computational model with Brownian agents [31].

In this study, we document the modeling approach used by the Norwegian Institute of Public Health to support policymakers with the COVID-19 vaccination strategy in Norway [32–35]. The approach includes an individual-based model (IBM) and a geographic-age-stratified meta-population model (MPM). Our aim is to perform a retrospective evaluation of the early Norwegian vaccination strategy and examine the effectiveness of alternative geographic prioritization strategies. Our mathematical models are calibrated to historical epidemiological and vaccination uptake data to capture the local COVID-19 transmission dynamics. To compare geographic prioritization strategies, we vary three key factors: (i) the number of vaccine doses redistributed, (ii) the number of municipalities prioritized, and (iii) the timing of the start of geographic prioritization. These strategies, along with the actual vaccination strategy, are evaluated relative to a national roll-out without geographic prioritization. To assess the effectiveness of these strategies, we explore separately outcomes of infections, hospitalizations, ICU admissions and deaths by calculating the relative risk reduction (RRR) of cumulative outcomes during the first seven months of 2021. Results of the two models are presented side-by-side to examine the robustness of results and highlight any potential structural biases.

## 4 Materials and methods

### 4.1. Data sources

The health data used in this study, covering the period from the 1st of January to the 31st of July 2021, were provided by the Norwegian emergency preparedness register for COVID-19 (Beredt C19) [36]. The database contains individually merged data collected from national health and other administrative registers. Data including positive tests, importations and deaths were obtained from the Norwegian Surveillance System for Communicable Diseases (MSIS) [37]. Additionally, data on hospitalizations and ICU admissions were collected from the Norwegian Intensive Care and Pandemic Registry (NIPaR) [38, 39]. Finally, the vaccination data were obtained from the Norwegian Immunization Registry (SYSVAK), a national electronic immunization registry that records vaccination status of all individuals [40, 41].

The data were pre-processed and aggregated by date, municipality, age, and risk status. The risk status, or medical risk group, refers to individuals with one or more defined diseases or conditions (e.g. organ transplantation, immunodeficiency, diabetes, etc.) that increase the risk of severe disease and death from COVID-19 [42]. Further details about the data used in this study can be found in S1 Appendix.

Ethics approval has been obtained for the use of data in this article, authorized through the Norwegian Health Preparedness Act, paragraphs 2-4. More information regarding this approval can be found at https://www.fhi.no/en/id/infectious-diseases/coronavirus/emergency-preparedness-register-for-covid-19.

### 4.2. The two geographic-age-structured models

Our study employs two stochastic mathematical models, an individual-based model (IBM) and a meta-population model (MPM), to conduct analyses of COVID-19 vaccine distribution scenarios in Norway. The models were developed to simulate the Norwegian population of approximately 5.4 million individuals, taking into account their demographics in terms of age and geographic location. Both models consider the actual age profile and geographic distribution of the population, and simulate the spread of SARS-CoV-2 from January to July 2021, when the majority of the first doses of vaccines were delivered to the population. The distribution of population size and density across the Norwegian municipalities is illustrated in Fig S9A and B in S1 Appendix. Moreover, we rely on unpublished social contact studies to determine the heterogeneity of contact patterns between individuals of different ages, while the contacts between and within regions are based on mobility data [43].

To capture the transmission dynamics of the virus, we employ a Susceptible-Exposed-Infectious-Removed (SEIR) transmission model that accounts for symptomatic individuals who may require hospitalization and have a risk of mortality due to severe infection. The models also incorporate interventions such as vaccination and restrictions of social contacts. The distribution of vaccines follows the priority order established by the Norwegian government, while the changes in non-pharmaceutical interventions (NPIs) and population contact behavior are captured by the transmission rates, as described in the model calibration section below.

#### 4.2.1. The individual-based model (IBM)

The individual-based model (IBM), also known as an agent-based model (ABM), captures the complex individual variability and local interactions involved in the transmission dynamics of COVID-19 in Norway. The 356 municipalities in Norway are subdivided into 13,521 cells, and individuals within each cell are assigned ages ranging from 0 to 100 years old. To account for different types of contacts, we construct several contact routes, including community, household, school, university, and workplace.

The community contacts are modeled as random processes based on the age of potential contacts and physical distance between cells. Relative contact rates among different age groups are also considered, with the highest contact rate observed among those aged 20 to 29 years according to the unpublished social contact studies. To reflect real-world mobility patterns, we employ a heavy-tailed distribution function based on mobility data from the largest telecommunication company in Norway, Telenor [43, 44]. This function results in a greater likelihood of short-distance transmission compared to long-distance transmission.

Contacts within households, schools, universities, and workplaces are uniformly distributed among all individuals belonging to those institutions. The IBM’s structure was originally developed to study the transmission dynamics of MRSA in Norway [45] and has been adapted to model COVID-19 transmission, as presented in this paper.

The code for the IBM is available at https://github.com/folkehelseinstituttet/COVID19 vaccination-IBM or https://github.com/imlouischan/corona-no. More details on the IBM can be found in S1 Appendix.

#### 4.2.2. The meta-population model (MPM)

The meta-population model (MPM) implements the same epidemiological model as the IBM, but with sub-populations defined by 9 age groups, 2 risk groups, and a varying number of geographic areas based on the prioritization strategy. Contact between age groups is based on Norwegian survey data, while contacts between and within areas are based on the mobility data from Telenor [43, 44].

The model is implemented in R [46] using the odin [47] and mcstate [48] packages.

The MPM impelementation is available at https://github.com/folkehelseinstituttet/COVID19 vaccination-MPM or https://github.com/Gulfa/regional vaccination. More details on the MPM can be found in S1 Appendix.

### 4.3. SEIR transmission and hospitalization model

We employed an SEIR transmission model, which includes pre-symptomatic, symptomatic and asymptomatic infectious compartments, as illustrated in Fig S3 in S1 Appendix. In brief, individuals who have not been exposed to the virus or the vaccine are susceptible (*S*). Upon exposure, susceptible individuals (*S*) enter a latent, non-infectious state (*E*_*a*_ or *E*_*s*_), before becoming infectious. Symptomatic individuals first enter a pre-symptomatic state (*I*_*p*_) before developing symptoms (*I*_*s*_), whereas those with asymptomatic infections (*I*_*a*_) are assumed to recover (*R*). Individuals with a symptomatic infection (*I*_*s*_) may recover (*R*), or develop severe illness requiring hospitalization (*H*). Hospitalization can result in discharge and recovery (*R*), or it can lead to ICU admission (*U*). ICU admissions (*U*) are followed by a second stay in the hospital ward (*H*), before discharge and recovery (*R*). Deaths (*D*) are assumed to occur from the symptomatic state or during hospitalization. The time spent in each compartment follows gamma distributions in the IBM and exponential distributions in the MPM, and the parameters are shown in Table S1 in S1 Appendix.

The probabilities of hospitalization and death depend on age and risk status, whereas the probabilities of ICU admissions depend solely on age. We consider that older people have a higher probability of hospitalization compared to younger people, and the risk increases exponentially with age, as reported by another study [49]. Similarly, people belonging to the risk group for severe illness have a higher probability of hospital admission and death than those who do not. For simplicity, we assume that the probability of ICU admission is independent of risk status. The probabilities of hospitalization, ICU admissions, and death are reported in Tables S3, S7, and S8 in S1 Appendix.

To account for regional differences during the simulation period, we scaled the transmission rate parameter *β* by a scaling factor for each municipality. These scaling factors, or relative reproduction numbers, were assumed to be constant and calculated based on the proportion of cumulative number of reported cases and population size between February 2020 and May 2021 in each municipality, and are provided in Section S2.2 in S1 Appendix. A map showing the distribution of regional scaling factors is also included in Fig S9C in S1 Appendix.

We assume a seasonal effect on transmission across all municipalities, with a relative difference of 50% between the maximum (in winter) and minimum (in summer). The seasonality is calculated based on the mean daily temperature for Norway and illustrated in Fig S2E in S1 Appendix.

We consider cases imported from abroad and calculate the daily number of importations using monthly data that are aggregated by age and county, as shown in Fig S2F in S1 Appendix.

The simulations begin on the 1st of January 2021 concurrent to the vaccine roll-out. To initialize the number of individuals in each compartment, we rely on the results of a separate model used by the Norwegian Institute of Public Health (NIPH) for situational awareness and forecasting of the COVID-19 pandemic in weekly runs, enabling the monitoring of the pandemic in Norway since March 2020 [43, 50, 51]. The situational awareness model provides the total numbers of all age groups and risk factors per county. Within each county, we assume a uniform distribution across municipalities, age groups, and risk factors. This ensures that the total population count is divided among all subpopulations based on their respective population sizes.

### 4.4. Vaccination model

Based on observational studies during the pandemic, which suggest similar real-world effectiveness, we assume that both vaccines (Comirnaty and Spikevax) are equally effective in our models [52].

We vaccinate individuals who are not infected at the time of vaccination and assume that the vaccines are “leaky”, meaning that the vaccine effect of both the first and second doses is imperfect [16]. This implies that vaccinated individuals have a lower risk of infection upon exposure compared to unvaccinated individuals. The vaccine effectiveness (VE) of the two doses is assumed to be a two-step monotonically increasing function over time, as shown in Fig S2D in S1 Appendix. For simplicity, the MPM incorporates the leaky scheme by utilizing separate compartments. We assume that people receive their second dose 12 weeks after the first dose. We also assume that the time to reach full effect is 14 days after the second dose vaccination, which is consistent with another study [53]. Given the relatively short simulation period of seven months, we decided not to consider waning effects of vaccine-derived immunity in this study.

We characterize vaccine effectiveness by five types of protection, namely against (i) symptomatic infection, (ii) asymptomatic infection, (iii) hospitalization, (iv) death, and (v) transmissibility. The first two effects against infection represent a reduction in susceptibility to the virus, while the last effect against transmissibility is translated into a reduction in infectiousness for infected people. Vaccine effectiveness parameters are provided in Table S2 and detailed information with references on the assumptions of vaccine effectiveness can be found in S1 Appendix.

### 4.5. The actual Norwegian vaccination strategy

The national government established guidelines for prioritization and administration of the vaccine program, while the municipalities were responsible for vaccinating their residents [10].

During the initial roll-out at the turn of 2020/2021, vaccines were allocated to municipalities based on the number of people above 65 years old exclusively. Elderly people including residents in nursing homes and those with underlying medical conditions were given priority to protect those at greatest risk of severe illness and death from SARS-CoV-2 infection [54]. In practice, healthcare workers were also given priority. In our models, we positioned healthcare workers after the elderly population, and the distribution of vaccines to municipalities was based on the size of at-risk populations, with priority given in age-descending order (refer to Table S10 in S1 Appendix for more information):

1. Elderly aged above 65 years (divided into three age categories)
2. Healthcare workers
3. Individuals aged 18-64 years with underlying medical conditions (divided into three age categories)
4. Adult population aged above 18 years without underlying medical conditions (divided into four age categories)

The geographic prioritization of COVID-19 vaccines in Norway was carried out in two steps. The initial step involved making a priority ranking of the 356 municipalities based on the cumulative hospitalization rates per resident population. On the 9th of March 2021, around the time most individuals above 85 years old had been vaccinated, the government decided to implement moderate geographic prioritization, whereby 20% additional vaccine doses were allocated to 10 municipalities, including the capital Oslo and four municipalities in the surrounding Viken county that had experienced consistently high infection levels over time [13]. These 10 municipalities, referred to as the *Plus* group, received vaccines at the expense of 330 municipalities, referred to as the *Minus* group, while the remaining 16 municipalities received the same number of vaccine doses and were referred to as the *Neutral* group. Later in March, the municipality distribution key was changed to population proportions of adults above 18 years old. The second phase began on the 19th of May 2021, around the time most individuals above 65 years old had been vaccinated, with a stronger geographic targeting of vaccines, where 60% extra doses were allocated to the prioritized regions [14]. This strategy involved adding 19 municipalities to the *Plus* group, primarily major cities and municipalities in Viken, while 319 and 13 municipalities were placed in the *Minus* and *Neutral* groups, respectively. The prioritization continued until all eligible adult individuals in the *Plus* group aged above 18 years had been offered vaccination.

To simulate the vaccination strategy used in Norway, we used vaccination data recorded by SYSVAK from January to July 2021. Real data from SYSVAK revealed that the prioritization plan was not consistently followed considering vaccine deliveries to each municipality and the vaccination of age groups within the municipalities, possibly due to vaccine hesitancy, logistical challenges, and unforeseen events. In our simulations, we proceeded by extracting the daily count per municipality, age, and risk status of individuals who received their first dose of the vaccine and assumed that they would all receive their second dose 12 weeks later. This assumption was supported by the high uptake rate of 90% among the adult population as of July 2021 (see Fig S10 in S1 Appendix), as well as the fact that 93% of those who received the first dose also received the second dose [36].

### 4.6. Model calibration

We calibrated our models to fit the national registry data between the 1st of January and the 31st of July 2021. The calibration process involved two steps, with the first step focusing on fitting the trend and age distributions of hospitalizations, and the second step focusing on fitting the age distributions of ICU admissions and deaths.

In March, the sharp increase in hospitalizations due to the third wave of COVID-19 infections and the arrival of the Alpha variant prompted the government to impose restrictions on social gatherings and travel. To account for changes in NPIs, we implemented two change points for the national transmission rate on the 28th of January and 11th of March 2021.

The calibration involved 12 free parameters including transmission rate parameters and susceptibility to infection parameters. The transmission rate parameters *β* controlled the time-varying behavior (contact rate) in three time periods, while the susceptibility to infection parameters described the relative probability of being infected upon exposure in each of the nine 10-year age group. Due to computational costs, the models were calibrated to fit the national daily hospitalization data by time and age, and we used a Latin hypercube sampling (LHS) to explore the parameter space. The goodness-of-fit was evaluated using a least squares method by the IBM, while the MPM used a Poisson likelihood.

The IBM was additionally calibrated to fit the proportions of the four transmission routes based on information provided by general practitioners. The data collected from general practitioners about the likely transmission revealed that 45.1%, 9.9%, 6.2%, and 38.6% of cases were infected within households, workplaces, schools, and other settings, respectively.

After calibrating the trend of hospitalizations and transmission, we estimated the probabilities of ICU admission given hospitalization and the probabilities of death given infection using age-stratified numbers of infections, hospitalizations, ICU admissions, and deaths over the entire simulation period. The fitted IBM and MPM to epidemiological data are shown in Fig 1. Both models estimate a significantly higher number (on the order of a factor two) of infections than reported cases, particularly in the younger age groups, indicating a high degree of under-ascertainment of cases [55]. Further details about the calibration of the two models can be found in S1 Appendix.

**Fig 1.**
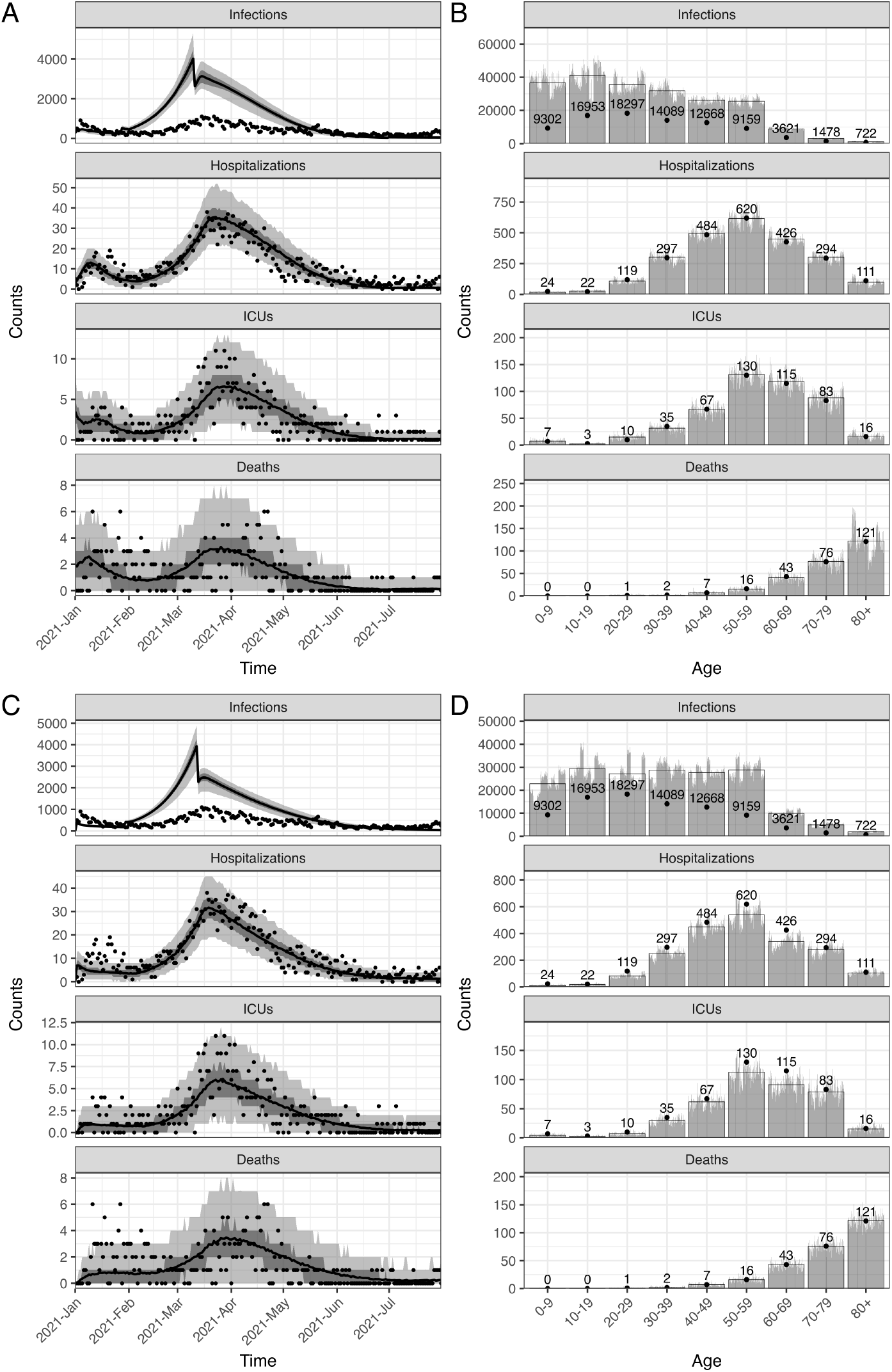
The calibrated individual-based model (IBM) and meta-population model (MPM) with data in the actual vaccination strategy. The upper and lower panels show the IBM and MPM, respectively. A and C: The time series data of all ages. The lines show the model fits with their 50 and 95% prediction intervals represented by gray areas. The dots show the observed data. B and D: The age distribution of total counts. The gray bars show the counts of each simulation and the full bars with black borders show the mean of all simulations. The data are shown in dots with their exact numbers. During the whole period, there were 86289 confirmed cases, 2397 hospital admissions, 466 ICU admissions, and 266 deaths registered.

#### 4.6.1. Sensitivity analysis on the hospitalization probabilities

Due to the lack of seroprevalence data in Norway, the precise number of infected individuals remains uncertain. To address the uncertainty surrounding the total number of infected individuals in Norway, we conducted a sensitivity analysis by doubling the probabilities of hospitalization given infection used in our main analysis. As a result of this sensitivity analysis, the model produced a number of infections that closely matched the reported cases. However, it is important to note that there may still be additional unreported cases that were not accounted for [55].

Nonetheless, in our main analysis, we incorporated the expected under-detection of cases by halving the probabilities of hospitalization given infection based on international literature sources [49, 56]. For a more detailed explanation of our assumptions, please refer to S1 Appendix.

### 4.7. Alternative strategies of geographic prioritization

We developed alternative geographic prioritization strategies for vaccine distribution based on the actual Norwegian vaccination strategy. We ensured the comparability of our models by keeping the calibrated parameters and setting the daily available doses identical across all strategies. As we distributed vaccine doses in pairs (first and second doses), all the numbers of doses refer to the first doses shown in Fig S11A in S1 Appendix.

We categorized the 356 municipalities into three groups: *Plus, Minus*, and *Neutral*, which corresponded to receiving extra doses, fewer doses, and the same number of doses, respectively. We simplified the geographic prioritization process into a single step and assumed that all municipalities followed the national guidelines for age and risk group prioritization. These strategies were defined by three key parameters:

- ∆*p*: The proportion of extra vaccine doses provided to the *Plus* group. The proportion received by each municipality is calculated relative to a national baseline strategy without geographic prioritization of eligible adult individuals aged above 18 years. We consider up to a maximum of 300% additional doses, corresponding to the municipalities in the *Plus* group receiving quadruple the number of doses relative to the adult population fraction.
- ∆*n*: The shift of municipality priority towards the *Plus* group (∆*n >* 0) or *Minus* group (∆*n <* 0). The selection of municipalities is based on the relative reproduction numbers of the regions. The shifts in either direction lead to a decreased population in the *Neutral* group, and the proportion of the population in the three groups changes accordingly. Geographic maps illustrating the distribution of various alternatives is presented in Fig 2, and more detailed descriptions of the shifts are provided in the following.

**Fig 2.**
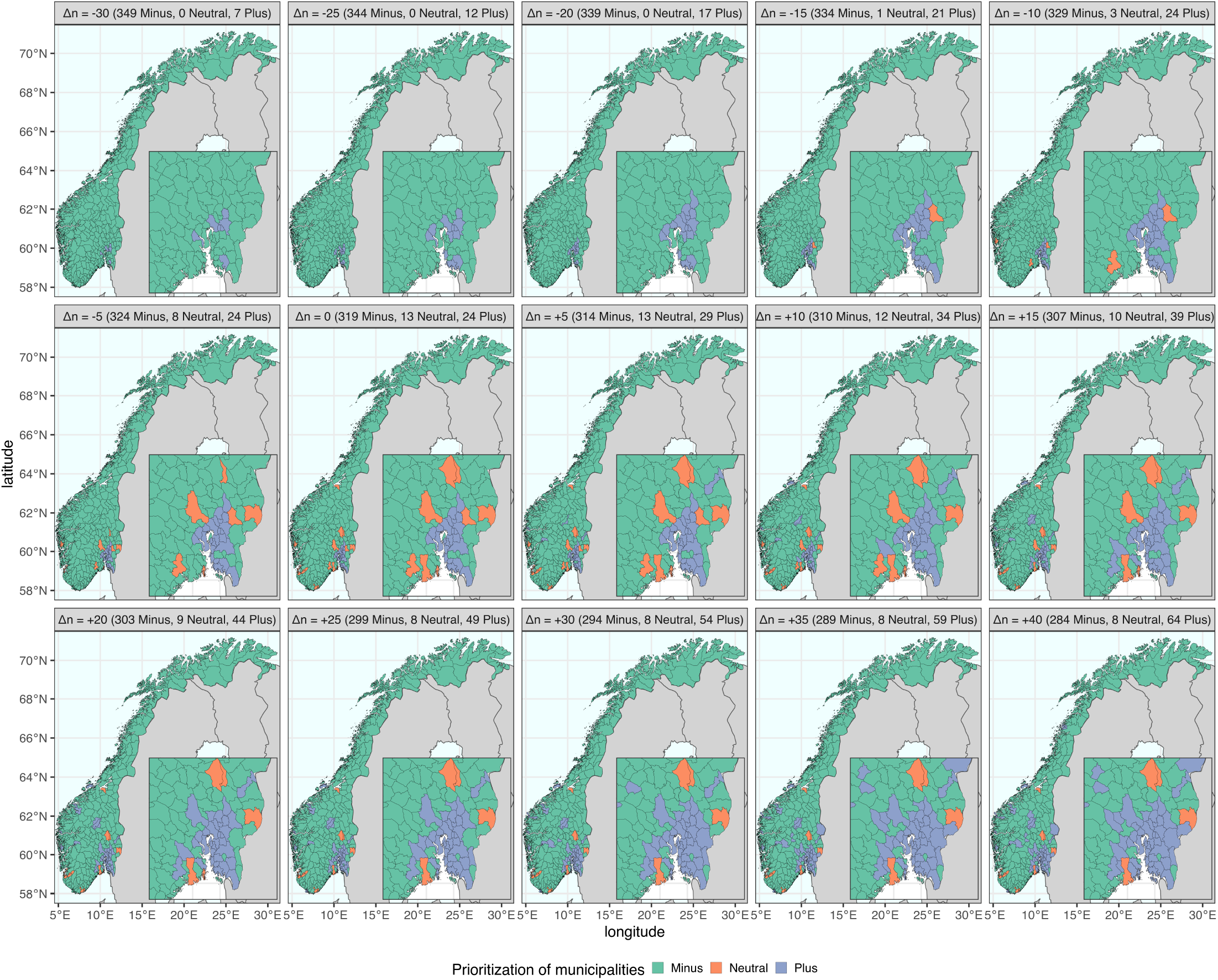
The geographic distribution of municipality prioritization in alternative strategies. The 356 municipalities are classified into three groups, represented by the colors green (*Minus*), orange (*Neutral*), and blue (*Plus*). Each strategy comprises a different number of municipalities in each group. The baseline strategy (∆*n* = 0) represents the selection made in the real-world implementation. The shifts of municipality priority ∆*n* are indicated in brackets, and the number of municipalities in each group is shown accordingly in each panel title.
- ∆*t*: The timing of the start of the vaccination program measured in months. The geographic prioritization strategy is assumed to be implemented on the first day of the months, i.e., on the 1st of January, February, March, April, May, June and July 2021.

To derive alternative strategies, we employed the same grouping of municipalities as the original selection implemented in the second step of the actual strategy, with 24, 13, and 319 municipalities in the *Plus, Neutral*, and *Minus* groups, respectively. Given the large differences in sizes of municipalities, such as Oslo accounting for 12% of the Norwegian population, while other municipalities may contain only less than a thousand people, it is essential to consider the proportion of the population that is prioritized. In original selection (∆*n* = 0), the *Plus, Neutral*, and *Minus* municipalities represent approximately 50%, 20% and 30% of the Norwegian population, respectively.

The alternative strategies were selected in the following way:

1. We rank each municipality according to their relative reproduction numbers, as described in Section S2.2 and Fig S12 in S1 Appendix. Alternative target areas are chosen by shifting the priority of five municipalities at a time, considering the ranking and their priority group.
2. To increase the prioritized geographic region relative to the adopted strategy (∆*n >* 0), we add municipalities to the *Plus* group (originally 24 municipalities) in successive steps. The top five highest ranked municipalities in the *Neutral* and *Minus* groups are moved to the *Plus* group. This procedure is repeated until 40 additional municipalities are shifted to the *Plus* group, i.e., 29, 34, …, and 64 municipalities are in the *Plus* group.
3. To increase the non-prioritized geographic region relative to the adopted strategy (∆*n <* 0), we add municipalities to the *Minus* group (originally 319 municipalities) in successive steps. The bottom five lowest ranked municipalities in the *Plus* and *Neutral* groups are moved to the *Minus* group. This procedure is repeated until 30 additional municipalities are shifted to the *Minus* group, i.e., 324, 329, …, and 349 municipalities are in the *Minus* group.
4. For each of the 14 selected geographic regions in Step 2 (∆*n >* 0) and Step 3 (∆*n <* 0), as well as the baseline strategy (∆*n* = 0), we select a value for prioritization of vaccine doses ∆*p* ranging from 0% up to a maximum of 300%, and a date of implementation ∆*t* from January to July 2021. The maximum value of ∆*p* is determined by the population ratio between *Minus* and *Plus*, which is presented in Table S9 in S1 Appendix. The number of extra vaccine doses provided to the *Plus* group for each strategy combination of ∆*n* and ∆*p* is shown in Fig S13 in S1 Appendix.
5. As a baseline for comparison of the alternative strategies, we consider a national strategy without geographic prioritization (i.e. ∆*p* = 0).

To evaluate the strategies, we conducted 1000 simulations in both IBM and MPM. We used 100 stochastic seeds for each of 10 calibrated parameter sets. Each simulation in the IBM took approximately 3 minutes, while each simulation in the MPM took approximately 20 seconds. All simulations were run in parallel on large computer clusters. We compared each iterated simulation of the alternative strategies to the national baseline strategy without geographic prioritization. To assess the reduction in health outcomes, we calculated the relative risk reduction (RRR) using the following formula:

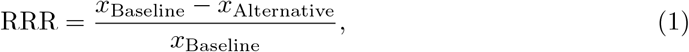

where *x* represents the cumulative infections, hospitalizations, ICU admissions or deaths over the entire simulation period. The RRR values indicate the amount by which the alternative strategies reduce the health outcomes compared to the baseline strategy. An RRR = 0 means that the alternative is the same as the baseline, while a positive RRR = 50% means that the alternative strategy reduces the cumulative health outcome by 50% compared to the baseline strategy. The higher RRR indicates the more effective the strategy to reduce the burden, and an RRR = 1 represents a disease-free situation. In contrast, a negative RRR = *−*100% or *−*200% means doubling or tripling of the outcomes, respectively, compared to the baseline strategy. The following results shows the mean values and their 95% confidence intervals (95%CIs) using 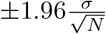, where *σ* is standard deviation of RRR and *N* = 1000 is the number of simulations.

In addition, to evaluate the actual strategy as a special case, we generated another national strategy without geographic prioritization, using the real vaccination data. We adjusted the vaccine distribution starting from the implementation of the first step of geographic prioritization (i.e., 9th of March 2021) based on the adult population (aged 18 years or older) in each municipality, while keeping all the data prior to that day. We generated 100 realizations corresponding to the 100 stochastic seeds for each of 10 calibrated parameter sets. We chose to compare the actual strategy with this national strategy instead of the fully controlled baseline strategy above for several reasons. Firstly, healthcare workers were given a high priority in the actual strategy, while we vaccinated healthcare workers after the elderly in the baseline strategy. Secondly, the actual strategy in January and February prioritized based on the population of people aged 65 years or older, while we considered all adults throughout the entire period in the baseline. This resulted in prioritized municipalities receiving more doses than in the actual strategy, primarily due to more younger people living in Oslo and its surrounding areas. Thirdly, both the proposed geographic and age prioritization was not strictly followed, especially during the initial phase, in the actual strategy as we could control in our baseline simulations.

## 5 Results

### 5.1. Evaluating the actual strategy

Table 1 presents the total mean number of infections, hospitalizations, ICU admissions, and deaths from the 1st of January to the 31st of July 2021 under the baseline strategy without geographic prioritization, along with the relative risk reduction (RRR) of the actual strategy and best strategies from both models. The RRR for infections, hospitalizations, ICU admissions, and deaths resulting from the actual strategy were nearly zero, implying that the actual strategy was similar to the baseline without geographic prioritization.

**Table 1.** The health outcomes and relative risk reduction (RRR) of different strategies compared to the baseline strategy without geographic prioritization.

| Model | Outcome | Baseline strategy | Actual strategy | Best strategy |  |  |
| --- | --- | --- | --- | --- | --- | --- |
| | | Count<br>(95% CI) | RRR, %<br>(95% CI) | RRR, %<br>(95% CI) | $\Delta p$, % | $\Delta n$ (Population, % in<br>*Minus*, *Neutral*, and *Plus*) |
| IBM | Infections, $10^3$ | 242.5 (241.2, 243.8) | -0.1 (-0.5, 0.3) | 16.4 (16.1, 16.7) | 300 | -25 (76.3, 0.0, 23.7) |
|  | Hospitalizations | 2688 (2676, 2699) | 0.3 (-0.1, 0.7) | 19.4 (19.0, 19.7) | 250 | -20 (72.7, 0.0, 27.3) |
|  | ICUs | 522 (520, 525) | 0.0 (-0.6, 0.5) | 19.2 (18.8, 19.6) | 250 | -20 (72.7, 0.0, 27.3) |
|  | Deaths | 235 (234, 237) | -0.7 (-1.3, 0.0) | 8.0 ( 7.5, 8.6) | 100 | +10 (46.6, 19.9, 33.4) |
| MPM | Infections, $10^3$ | 195.9 (194.9, 197.0) | -0.2 (-1.1, 0.6) | 16.0 (15.7, 16.3) | 300 | -25 (76.3, 0.0, 23.7) |
|  | Hospitalizations | 2188 (2177, 2199) | 0.1 (-0.7, 0.8) | 15.4 (15.0, 15.7) | 300 | -25 (76.3, 0.0, 23.7) |
|  | ICUs | 419 (416, 421) | 0.1 (-0.8, 0.9) | 15.6 (15.1, 16.1) | 300 | -25 (76.3, 0.0, 23.7) |
|  | Deaths | 240 (239, 241) | 0.2 (-0.5, 0.9) | 5.5 ( 4.9, 6.2) | 70 | -15 (69.5, 0.4, 30.1) |

The third column shows the total mean number (and their 95% confidence intervals) of infections, hospitalizations, ICU admissions, and deaths in the two models. The mean RRR (and their 95% confidence intervals) of the actual strategy and the best strategies are shown in the next two columns. The best strategies, selected separately for each health outcome, are shown in the last two columns, and were based on the starting time ( $\Delta t$ ) of the 1st of January 2021. The brackets in the last column indicate the population fraction of three groups (*Minus*, *Neutral*, and *Plus*).

### 5.2. Alternative municipality priority

In Fig 3, we present the national RRR from two models for various combinations of proportions of extra doses (∆*p*) and shifts in municipality priority (∆*n*) for alternative strategies, given the starting time (∆*t*) of geographic prioritization on the 1st of January 2021. The leftmost column shows the baseline strategy without geographic prioritization (∆*p* = 0%) does not minimize any health outcome.

**Fig 3.**
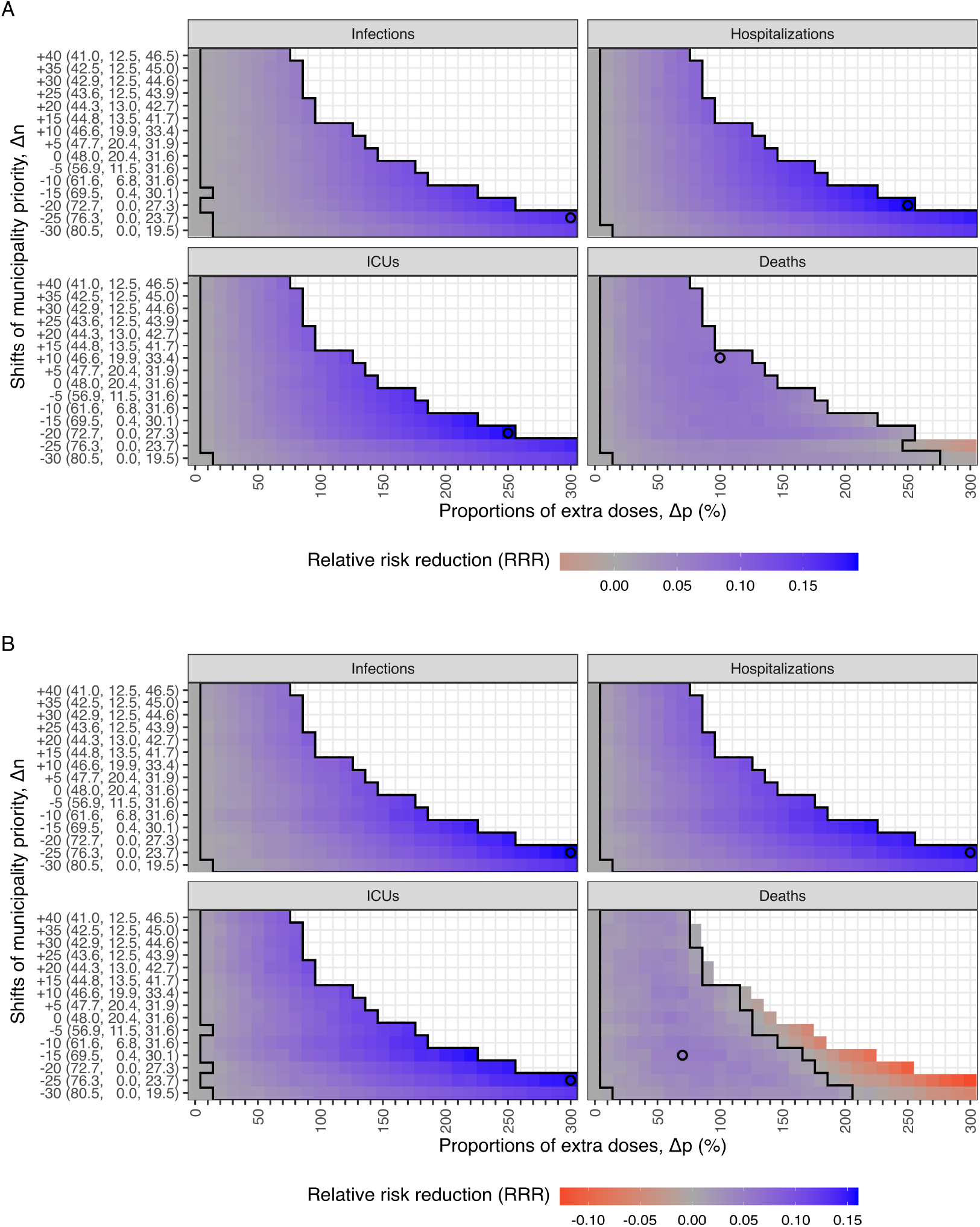
The mean relative risk reduction (RRR) of health outcomes for the alternative strategies modeled with (A) IBM and (B) MPM. The mean RRR of infections, hospitalizations, ICU admissions, and deaths is represented by color, ranging from red (negative RRR) to blue (positive RRR). The black empty circles and larger areas illustrate the strategies with the highest mean RRR and positive 95% confidence intervals for each outcome. To minimize infections, hospitalizations, or ICU admissions, the strategies with higher priority levels lead to higher mean RRR. However, for minimizing deaths, the highest level of prioritization has a negative impact, while medium levels of prioritization lead to the highest mean RRR. The y-axis shows the population fractions (%) of three groups (*Minus, Neutral*, and *Plus*) for each shift in municipality priority (∆*n*). The geographic distribution of municipality priority (∆*n*) can be found in Fig 2.

To minimize infections, a strong geographic prioritization to a small number of municipalities with high transmission levels is optimal. The best strategy involves reducing the priority by moving 25 municipalities to the *Minus* group (∆*n* = − 25) and giving 300% extra doses to the *Plus* group (∆*p* = 300%), resulting in the highest mean RRR (and their 95% CIs) of 16.4 (16.1 to 16.7)% and 16.0 (15.7 to 16.3)%, which is equivalent to saving 40,063 (39,190 to 40,936) and 31,563 (30,905 to 32,221) infections in the IBM and MPM, respectively. Similar strategies also minimize hospitalizations and ICU admissions. In the IBM, the optimal strategy involves slightly shifting the priority to ∆*n* = − 20 and ∆*p* = 250%, which produces the highest mean RRR (and their 95% CIs) of 19.4 (19.0 to 19.7)% and 19.2 (18.8 to 19.6)%, resulting in saving 524 (514 to 534) hospitalizations and 102 (99 to 104) ICU admissions, respectively. In the MPM, the optimal strategy is ∆*n* = − 25 and ∆*p* = 300%, with the mean RRR (and their 95% CIs)) of 15.4 (15.0 to 15.7)% and 15.6 (15.1 to 16.1)% for hospitalizations and ICU admissions, respectively, corresponding to saving 342 (334 to 351) hospitalizations and 67 (65 to 69) ICU admissions.

To minimize deaths, a moderate level of geographic prioritization to a larger number of municipalities is likely optimal, unlike the strategies that minimize infections, hospitalizations, or ICU admissions. In the IBM, increasing the priority by moving 10 municipalities to the *Plus* group (∆*n* = +10) and giving 100% extra doses (∆*p* = 100%) is the optimal strategy, which can avoid 20 (95%CI: 18 to 21) deaths with a mean RRR of 8.0 (95%CI: 7.5 to 8.6)%. In the MPM, the strategy that minimizes deaths is ∆*n* = − 15 and ∆*p* = 70%, with a mean RRR of 5.5 (95%CI: 4.9 to 6.2)%, corresponding to avoiding 15 (95%CI: 13 to 16) deaths. However, choosing the prioritization strategy that minimizes infections at ∆*n* = − 25 and ∆*p* = 300% results in a negative effect on deaths with mean RRR of -4.3 (95%CI: -5.0 to -3.6)% and -12.7 (95%CI: -13.4 to -12.0)%, leading to 10 (95%CI: 8 to 11) and 29 (95%CI: 28 to 31) extra lives lost compared to the baseline strategy in the IBM and MPM, respectively.

The gains in the *Plus* group are similar across all health outcomes, but the losses in the *Minus* group can be substantial under higher level of prioritization strategies. Fig S15 in S1 Appendix shows the trade-off between municipalities, i.e. the *Plus* and *Minus* groups. Similarly, Fig S16 in S1 Appendix shows the trade-off between age groups. When more doses are prioritized, the protection of the younger population increases, resulting in a higher average age of infections.

Taking the optimal strategy for minimizing infections in the IBM as an example, Fig 4 shows the trade-off between municipalities by plotting the RRR of each municipality on a map. The municipality-specific RRR of infections, hospitalizations, ICU admissions, and deaths are positive in Oslo and its surroundings because they receive more doses and hence prevent larger local outbreaks. Although the national benefit is larger than the losses in terms of infections, hospitalizations, and ICU admissions, it is not for deaths. Non-prioritized municipalities experience a significant negative impact under higher level of geographic prioritization, creating a large trade-off between age groups. Fig S14 in S1 Appendix shows the trade-off by age groups under different geographic prioritization strategies.

**Fig 4.**
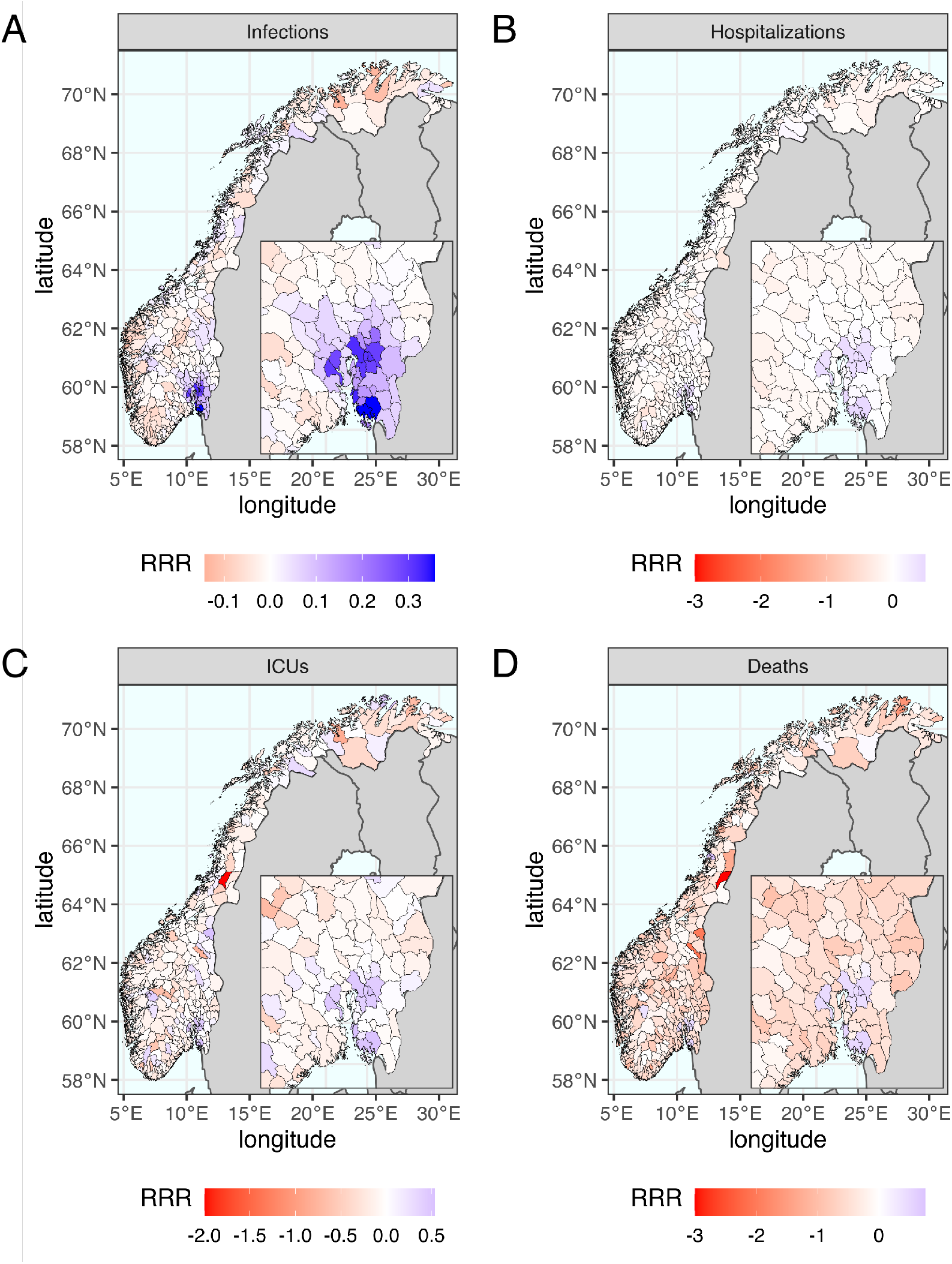
The geographic distribution of mean relative risk reduction (RRR) of the optimal scenario that minimizes infections. The optimal scenario is the one of reducing the priority by moving 25 municipalities to *Minus* and giving 300% extra doses (∆*n* = − 25, ∆*p* = 300%). The geographic trade-off between municipalities is illustrated by color from blue to red representing mean values of municipality-specific RRR from positive to negative. The benefits in Oslo and its surroundings are much greater than the drawback in other municipalities to minimize (A) infections, (B) hospitalizations and (C) ICU admissions but not (D) deaths.

### 5.3. Alternative starting time

The starting time of changing from the national to geographic prioritization strategy is also a key factor in the strategy for prioritizing vaccines. We selected a subset of the combinations of ∆*p* and ∆*n*, and explored alternative implementation dates ∆*t* ranging from January to July 2021. Our results indicate that earlier implementation yields better control by reducing adverse health outcomes across most prioritization strategies.

Fig 5 shows that the RRR of infections, hospitalizations, and ICU admissions decreases by the starting time across all alternative prioritization strategies. However, the RRR of deaths decreases only when prioritizing in mild and moderate levels (approximately ∆*p <* 200%). For higher priority levels (approximately ∆*p >* 200%), initiating the geographic prioritization in February or March results in a higher RRR of deaths compared to initiating in January, as shown Fig S17 and Fig S18 in S1 Appendix. Nevertheless, the moderate level (∆*p* = 70% − 100%) with a starting time in January as the previous section shown, the RRR of deaths is the highest among all strategies. Additionally, we found that all the RRR approach zero if the starting time is after May 2021.

**Fig 5.**
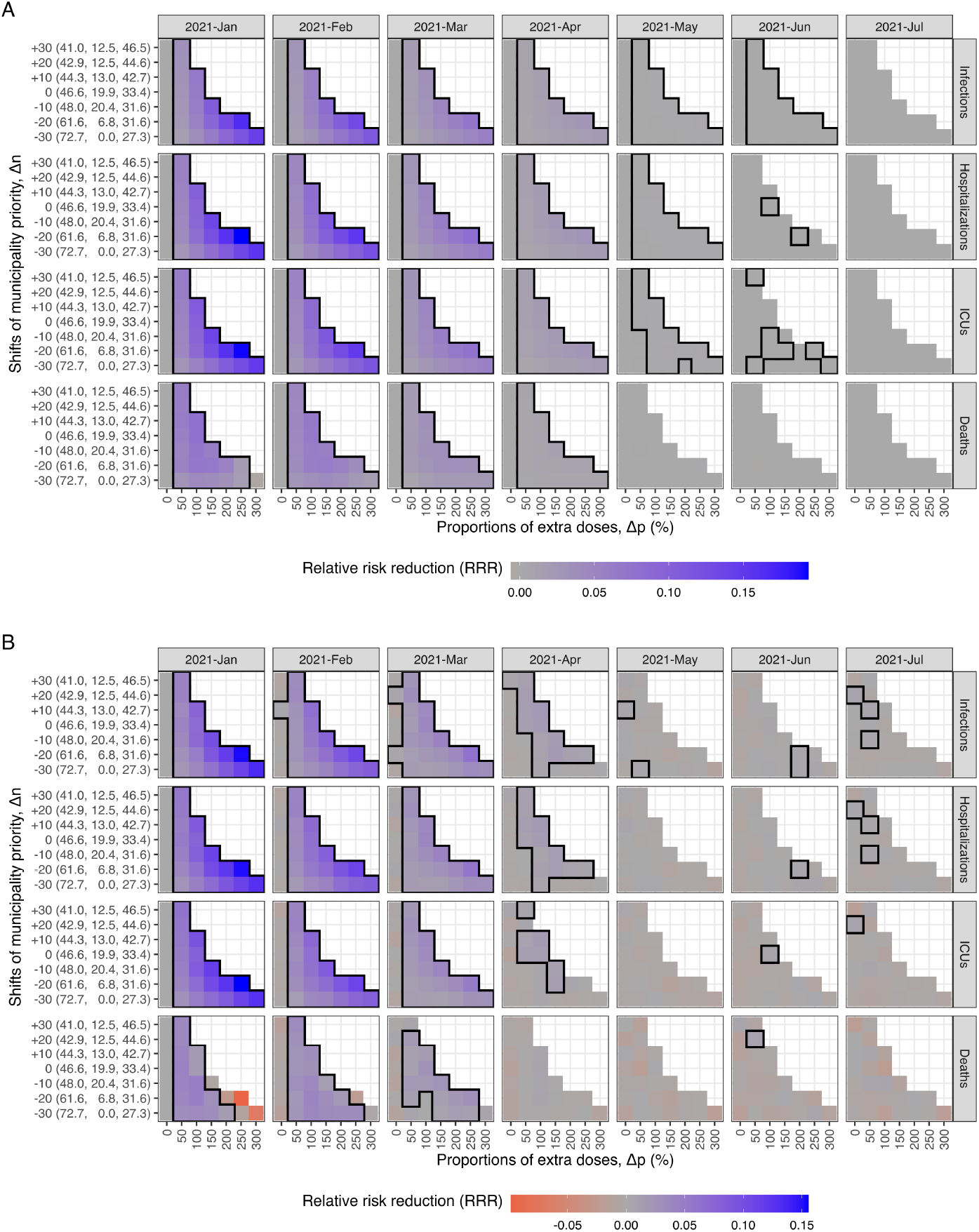
The mean relative risk reduction (RRR) of health outcomes for the alternative starting time modeled with (A) IBM and (B) MPM. The mean RRR of infections, hospitalizations, ICU admissions, and deaths is represented by color, ranging from red (negative RRR) to blue (positive RRR). The panels of columns correspond to alternative start time, with the first column reflecting the results shown in Fig 3. The RRR of infections, hospitalizations, and ICU admissions decrease by starting time, considering all alternative strategies. However, the RRR of deaths decrease only when mild and moderate priority levels (approximately ∆*p <* 200%) are implemented. The black square areas indicate positive mean values and their 95% confidence intervals. The benefits are limited if the geographic prioritization started after May 2021.

### 5.4. Sensitivity analysis on the hospitalization probabilities

Doubling the probabilities of hospitalization does not significantly change the optimal strategies for minimizing infections, hospitalizations, ICU admissions, and deaths. For minimizing infections, hospitalizations, and ICU admissions, the strategies remain similar to those with the highest prioritization level. The moderate level of prioritization is still preferred for minimizing deaths. Similarly, implementing geographic prioritization after January leads to a decrease in RRR, which reaches nearly zero with no benefits if it starts after May 2021. For a more detailed comparison of the two assumptions on hospitalization probabilities, please refer to S1 Appendix.

### 5.5. Comparing the two models

Fig 6 presents a comparison between two models regarding the optimal strategy for minimizing infections. Apart from the initial growth being more rapid in the IBM, the primary discrepancy between the two models is the number of infections on the 28th of January and 11th of March 2021, when the transmission rates *β* undergo a change. Specifically, the infection curve from the MPM decreases instantly and more rapidly than that from the IBM. Moreover, the MPM correspondingly exhibits lower trends of hospitalizations and ICU admissions compared to the IBM. However, the MPM shows a higher trend of deaths than the IBM, attributed to the diverse age composition between the two models. More specifically, the IBM shows a greater number of infections among the younger population, whereas the MPM records more deaths in the oldest age group (aged 80 years or older). Furthermore, the complexity of the IBM, which includes heterogeneous contact structures in four transmission routes, contributes to additional variations. For a detailed comparison of the actual vaccination strategy for calibration and the counterfactual scenario without vaccination, please refer to Fig S20 and Section S5 in S1 Appendix, respectively.

**Fig 6.**
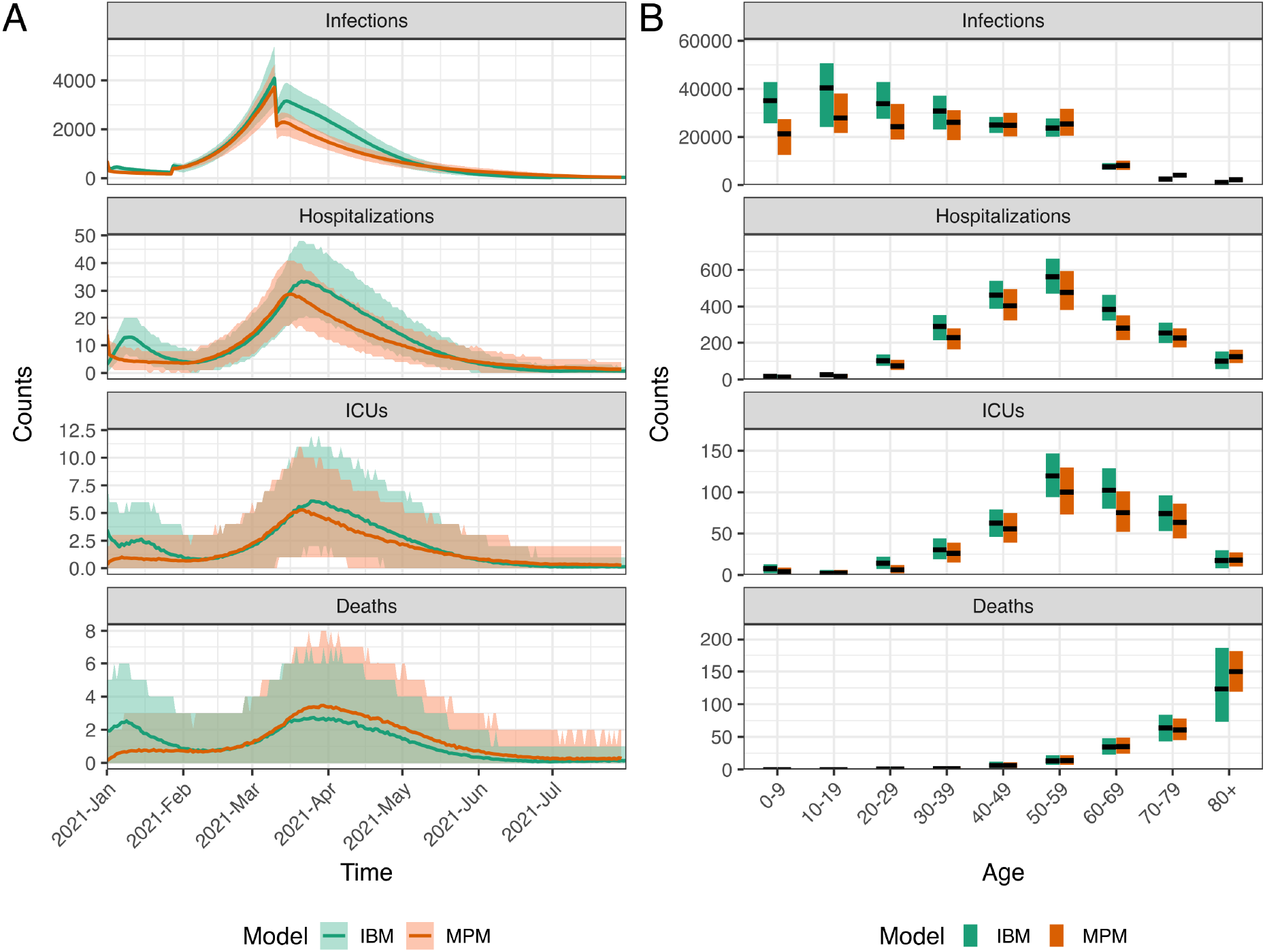
The comparison of two models for the optimal strategy for minimizing infections. A: The time series data of all ages. The lines show the model fits with their 95% prediction intervals represented by colored areas. B: The age distribution of total counts. The black lines show the median values, and the colored areas show the 95% prediction intervals.

## 6 Conclusion and Discussion

Early decision-making concerning the allocation of COVID-19 vaccines was critical, given the limited supply of vaccines. Mathematical models provide a powerful tool for investigating and quantifying the impact of different vaccination strategies. However, during the beginning of 2021, inherent uncertainties related to lack of data, the complex dynamics of the COVID-19 pandemic, its uncertain future trajectory, and the lack of knowledge regarding vaccine effectiveness and vaccine supply combined with the need for speedy results made such model-informed assessments challenging [57]. Specifically, one difficulty encountered during the pandemic was the accurate estimation of vaccine effectiveness against transmissibility. The vaccines demonstrated significant efficacy in inhibiting the transmission of the wild-type Wuhan variant. However, this effectiveness diminished with the emergence of each new variant, such as the Alpha variant.

In this study, we conducted a retrospective analysis to evaluate the geographic vaccination strategies for COVID-19 based on the outcome number of infections, hospitalizations, ICU admissions and deaths in Norway. By comparing alternative vaccination scenarios to the implemented strategy and using historical epidemiological data, we propose optimized vaccination strategies that provide valuable insights for guiding policy decisions and enhancing pandemic preparedness in the future.

Our findings demonstrated that early geographic vaccine allocation to areas with high infection levels could reduce COVID-19-related health outcomes. However, the optimal geographic deployment of vaccines, considering the geographic scope and the proportion of redistributed vaccines, depends critically on the specific objective, such as minimizing infections, hospitalizations, ICU admissions, or deaths.

For minimizing infections, hospitalizations, or ICU admissions, we found that prioritizing 12-17 municipalities, comprising approximately 25% of the Norwegian population, with the highest infection rates at the beginning of the vaccination campaign, and increasing their vaccine doses by a factor of 3-4, is crucial. This approach ensures earlier vaccination of younger people in those regions while delaying vaccination of older people in the non-prioritized regions, thereby offering direct protection to those at higher risk of infection and indirect protection in high transmission areas, consequently reducing the spread in other parts of the country through internal mobility. This finding is consistent with results by Monod et al. [58], which identified the age group of 20-49 years as the main group sustaining the pandemic, making vaccinating this group in high transmission areas earlier an effective way to control the pandemic. However, it is important to note that targeting a small number of municipalities or a minor proportion of the population can negatively impact mortality and lead to more deaths compared to the baseline strategy without geographic prioritization. This effect is caused by the significantly higher infection-fatality-ratio among older people and the slower vaccine uptake in that age group within low-priority regions. Notably, this pattern was not observed for hospitalizations because most admissions in Norway occurred in the 50-70 year age group, while the average age of those dying from COVID-19 was above 80 years old [59].

To minimize deaths, an early and geographically extended prioritization encompassing 21-34 municipalities comprising approximately 30% of the Norwegian population (alongside approximately 10 *Neutral* municipalities representing approximately 20% of the Norwegian population) and a more moderate level of priority, approximately doubling their vaccine doses, yielded the most favorable outcomes. The optimal strategies differed somewhat between the two models, likely due to slightly different population compositions. The results highlight the importance of determining the primary purpose of the vaccination program. In Norway, the government appointed an ethics committee that concluded a vaccination program aimed at minimizing the number of deaths should be chosen [60]. As we demonstrate, this leads to a significantly different optimal strategy than if the objective were to reduce hospitalizations, for instance. Moreover, we anticipate that minimizing disability-adjusted or quality-adjusted life year lost would also necessitate more geographic prioritization, rather than solely focusing on deaths [61].

Our findings are consistent with previous studies on geographic prioritization [28–31], which also recommended prioritizing high incidence areas. This is due to the higher local transmission rate from a geographic point of view, which is controlled by the regional reproduction numbers in our models. Regions with higher reproduction numbers are the major hubs spreading the disease both internally (within municipalities) and externally (between municipalities). Internally, growth rates of infections are exponential-like expanding given *R*_*t*_ *>* 1 even though vaccines are available. Externally, infectious individuals traveling between regions facilitate the spread of the virus, acting as sources of transmission. We assume that people moving between geographic cells follow the distance function, so short-range movement is more likely to happen. For example, Oslo and its neighboring municipalities form a cluster with high infection rates, and suppressing the growth of infections in the cluster before it becomes unstoppable is notably essential.

Furthermore, the geographic prioritization across populations is associated with age subsequently because of the individual-level prioritization within populations. The trade-off between infections and deaths is mainly due to the age-specific contact and risk as shown in several studies on age prioritization [16–27]. Many of the studies agree that prioritizing the elderly is the optimal strategy to prevent deaths directly, and so as our assumptions in the models to prioritize in a descending age order, starting with those older than middle-aged with higher risk. The selection of strategies between the direct and indirect effect is eventually to protect the older population.

In Norway, the actual strategy implemented was a moderate level of geographic prioritization, which aimed to maintain a good balance between high-risk and low-risk areas. However, our study found that the actual strategy was similar to the strategy without geographic prioritization, mainly because the second phase of the geographic prioritization as a core part was implemented in May 2021, which was too late to control the pandemic. In fact, this is consistent with our results on starting time.

Our study also highlights the importance of starting geographic prioritization as early as possible in reducing infections, hospitalizations, and ICU admissions. The time switching from the baseline strategy to geographic redistribution could play a significant role during the first three to four months of the implementation. Implementing geographic prioritization after May 2021 was less effective for two primary reasons.

First, the main wave of infections had already occurred in March, and the transmission had decreased to *R*_*t*_ *<* 1 as a result of extensive societal lockdown measures. The number of infections in May only reached one-fourth of the peak, making the vaccination strategy at that point less critical. This emphasizes the necessity of geographic prioritization during the first few months to reduce infections and subsequently severe illness. Second, most of the elderly, especially those above 80 years old, had already been vaccinated before May.

Postponing the implementation of geographic prioritization of vaccines reduce the RRR of outcomes on infections, hospitalizations, and ICU admissions. However, when considering a high level of geographic prioritization (such as more than tripling the number of doses to prioritized municipalities), postponing the implementation by one or two months could accelerate vaccination of the oldest age group and directly prevent deaths. Conversely, for mild or moderate levels of geographic prioritization, earlier implementation would be consistently a more effective strategy. This trade-off between priority level and start time is dependent on the decision to minimize either infections, hospitalizations, and ICU admissions, or deaths. Therefore, while earlier implementation of geographic prioritization could prevent more cases, it might not necessarily result in a substantial reduction in deaths. Conversely, postponing the implementation tended to converge towards the national distribution strategy. To make a good balance, there were two potential solutions: (i) implementing an earlier geographic strategy with a moderate level, or (ii) postponing a high priority level of geographic strategy would allow vaccine doses to be used more efficiently in reducing all infections, hospitalizations, ICU admissions, and deaths across the population. However, this presents a difficult ethical consideration during the pandemic [62].

We employed two models in parallel to evaluate the vaccination strategies, and the agreement between the two modeling approaches was essential to validate the results as shown in other studies comparing large-scale computational approaches [63, 64]. The same practice was done in the real-time analysis during the COVID-19 pandemic to determine recommendations of the vaccination strategies in Norway. The national recommendation reports by NIPH are available online [32–35, 65–76]. As an example, the geographic prioritization strategies were evaluated by our two models in the middle of distributing vaccines in February and March 2021 [32–35]. The use of our two models not only assisted in error detection during development but also provided a backup plan for implementation issues, particularly when working under tight deadlines. This retrospective study serves as a valuable tool for preparedness planning, providing valuable insights on how to allocate resources (i.e., vaccines) and how to prepare for future pandemics. In situations with new respiratory viruses where we can assume a high degree of geographic variation in transmission, it is important to consider geographic prioritization of a limited vaccine supply from the first dose given. This study provides an example of the importance of such prioritization and the value of large-scale computational modeling in public health decision-making.

In this study, it is crucial to consider several key assumptions and limitations when interpreting the results. First, we incorporated various simplifications concerning the properties of the virus and its transmission rates throughout the simulation period. We did not account for the growth of the Delta variant from July 2021, resulting in an underestimation of infections during the final phase. However, it is important to acknowledge that the impact of vaccination and its prioritization was beneficial during the Delta variant wave and in the longer term. The effectiveness of the prioritization strategy could have been even greater if potential cases that might have gone undetected were taken into consideration. Furthermore, we acknowledge that Norway, along with several other European countries, implemented a strict societal lockdown, resulting in a significantly lowered transmission rate during the spring. This shutdown of society created an exceptionally unnatural situation. In a scenario without such strong interventions, the effects of vaccine prioritization would likely have been different. Additionally, as our focus was on the short simulation period of 7 months, we did not account for waning vaccine immunity, despite the fact that immunity against infection may decline before the completion of 7 months. Future studies should explore the impact of waning immunity and booster doses given in a longer time frame.

Second, we assumed that all healthcare workers were vaccinated after the elderly (aged 65 or above) in our alternative strategies. This resulted in the elderly aged 65 years or older being vaccinated earlier than in the actual vaccination strategy, leading to an underestimation of the number of deaths in alternative scenarios. This differs from the actual vaccination strategy, which prioritizes front-line essential workers who have a higher risk of preventing outbreaks in hospitals and supporting the healthcare system. However, we assumed that these healthcare workers had the same contact rates as the general population in our models.

Third, while comparing different strategies, we kept the same transmission rates *β* as calibrated using the actual vaccination strategy. We assumed that the transmission rates vary with a seasonal effect, which is higher in winter and lower in summer [77]. The only difference was the plan of the vaccination program. We assumed that measures such as NPIs would remain exactly the same in all alternatives, although some regions could have large outbreaks. National interventions were captured by the transmission rates at three time intervals, while no regional interventions were considered in this study. We also assumed that the relative reproduction numbers of each municipality were stationary (i.e., constant in time). This made vaccination the only intervention varying in time between regions.

Fourth, our estimates of the regional scale factors for each municipality were based on several simplifying assumptions, potentially impacting the obtained results. Further research is needed to better describe the geographic variation in transmission rates among the municipalities.

Fifth, a key limitation of this study is that the contact matrix used was obtained during the pre-pandemic period. It is likely that the pandemic has changed contact patterns, with older populations likely reducing their contacts more in the first half of 2021 [78]. Regarding our estimates on the susceptibilities to infection, which decrease by age (except aged 50-59 years), this differs from the findings in other modeling studies [79–81] and cohort studies [82, 83]. Nonetheless, our estimates effectively capture the dynamics of transmission and address the distinction in their contact behavior. The decreasing susceptibilities could be associated with their contact patterns, the age-specific risk ratio of the Alpha variant estimated in Norway [56], or the assumed age-independent vaccine effectiveness. However, all models fitted well to the data and projected more infections than tested cases to reveal under-detection, especially of asymptomatic people or those with mild symptoms.

Finally, we assumed unrealistically that the capacity and logistics of vaccine distribution could always accommodate our scenarios. While the overall logistics in Norway were generally well developed, establishing vaccine centers for preparedness may require additional resources. Two main resources were needed when giving vaccines to the population: the number of doses in stock and human resources needed to vaccinate individuals in the front-line. We fixed the number of (first) doses per day nationally in all scenarios in accordance with the former, but there might be limits in the number of doses for each region, which might realistically cause problems due to the latter. Additionally, there might not be enough staff capacity or time to transport and handle large amounts of doses, although we prioritized some high-risk regions by giving 300% extra doses. Similar limitations exist regarding the assumption of the timing of receiving the second dose to be 12 weeks for everyone. Future studies should address the issue of distributing vaccines with different time delays between two doses as reported by the NIPH [35] and some studies [84–86].

To conduct a more comprehensive evaluation of geographic vaccination strategies, it is crucial to consider additional factors [61]. These factors include the impact of lockdown measures, the assessment of both short-term and long-term health outcomes, disease burden estimates that encompass the extended effects of ICU stays and post-infection complications or long COVID, as well as the implications of healthcare prioritization in regions characterized by high infection rates.

During a pandemic, social acceptability plays a crucial role in vaccine distribution. Prioritizing high-incidence areas with a larger allocation of doses can result in smaller municipalities facing inadequate supplies, necessitating a temporary halt to their vaccination programs. The evolving public sentiment surrounding the vaccination policy underscores the potential consequences on public trust and support. Therefore, it is essential to strike a balance between ensuring equitable distribution and maintaining public confidence in vaccination efforts, highlighting the ethical considerations that arise in this context [62].

In conclusion, our study provides important insights into the effectiveness of different COVID-19 vaccine distribution strategies in reducing mortality and morbidity in Norway. Our results suggest that geographic prioritization of vaccines can improve health outcomes during the initial phase of the vaccination program. However, the optimal level of geographic prioritization depends on the specific health outcomes targeted, and a moderate level of prioritization may be optimal for minimizing deaths. It is important to note that our analysis is based on modeling and retrospective data analysis. Employing these methodologies is particularly significant, as they enable the integration of supplementary information that was not available during the initial phase. While the policy of geographic prioritization in Norway has helped reduce the number of deaths, it may not have been optimized to minimize infections or avoid local outbreaks. Nonetheless, our findings can provide valuable guidance for policymakers in other countries or for future outbreaks, helping them make informed decisions about vaccine distribution strategies.

## Data Availability

The aggregated case data and computational code for two models are accessible to the public and shared on GitHub, available at https://github.com/folkehelseinstituttet/COVID19_vaccination-IBM or https://github.com/imlouischan/corona-no, and https://github.com/folkehelseinstituttet/COVID19_vaccination-MPM or https://github.com/Gulfa/regional_vaccination.
However, due to privacy considerations, vaccination data is not accessible to the public. Instead, demonstration of pseudo data with added noise on the original data is used.
The mobility data was collected and provided by Telenor Norway. Requests for access to mobility data should be directed to Telenor Research via.

https://github.com/folkehelseinstituttet/COVID19_vaccination-IBM

https://github.com/imlouischan/corona-no

https://github.com/folkehelseinstituttet/COVID19_vaccination-MPM

https://github.com/Gulfa/regional_vaccination

## 7 Supporting information

**S1 Appendix. The supplementary document**. This includes more detailed descriptions of the models and additional results.

## 8 Acknowledgments

The project was funded by the Research Council of Norway “COVID-19 in Norway: A real-time analytical pipeline for preparedness, planning and response during the COVID-19 pandemic in Norway” grant number 312721. The mobility data was collected and provided by Telenor Norway.

## S1 Appendix The supplementary document

Modeling geographic vaccination strategies for COVID-19 in Norway

### S1 Model descriptions

Here we give more detailed descriptions of the two models: the individual-based model (IBM) is a highly detailed model while the meta-population model (MPM) uses a population-based structure to describe the spread.

#### S1.1 Individual-based model (IBM)

In this study, we adapt a stochastic IBM previously used for studying Methicillin-resistant *Staphylococcus aureus* (MRSA) transmission [45] to model the spread of COVID-19. Our model incorporates individual-level factors such as age, location, occupation, risk status, epidemiological status, hospitalization status, and vaccination status. We characterize our synthetic population using Norwegian census data, and assign individuals to households based on population density and age distribution in 13,521 contiguous areas known as Grunnkrets in Norwegian or Basic circuits in English [87].

Occupation locations, including schools, universities, workplaces, and those who are unemployed or retired, are assigned based on age. The simulated age-specific contact matrices in different routes are depicted in Fig S1. Our model aligns with other similar models used to study different interventions against COVID-19 [18, 20, 27, 88–90].

##### S1.1.1 Households

To generate a distribution of household sizes for individuals, we utilize a two-dimensional size-age distribution, as illustrated in Fig S2A, from Statistics Norway [92]. Specifically, individuals are assigned to households ranging in size from 1 (i.e., single living) to a maximum of 7 individuals, with households determined based on the size-age distribution. Individuals aged 15 years and younger are assigned to households with at least two individuals. Individuals aged 88 years and older are aggregated into a single category, as data availability indicates that they are primarily living alone. The methodology underlying household construction follows the algorithm used in [45, 91].

##### S1.1.2 Occupations

Individuals are assigned in schools, universities or workplaces according to their ages and specific places according to size distributions of occupations. Specifically, children between 1 and 5 years old attend kindergarten, those between 6 and 15 years old attend primary school, those between 16 and 18 years old attend secondary school, and those over 19 years old attend university. Individuals who are 17 years old or older and not attending university or unemployed are assigned to workplaces [45].

In the context of the COVID-19 pandemic, it is assumed that 20% of employees work from home as a result of Norwegian interventions implemented in 2021. In addition, some individuals are specifically assigned to work as healthcare workers based on an age distribution, and they are given priority for vaccination within the population.

##### S1.1.3 Community

In the individual-based model, contact between individuals can occur through community activities as long as they are not completely isolated from society. Community contacts refer to interactions that take place outside of the home, school, or workplace, and may include activities such as taking public transportation, grocery shopping, dining out, socializing with friends, and visiting non-cohabiting family members. Age-specific contact data were collected in Norway in 2017 and show that younger individuals have higher rates of community contact than older individuals.

**Fig S1.**
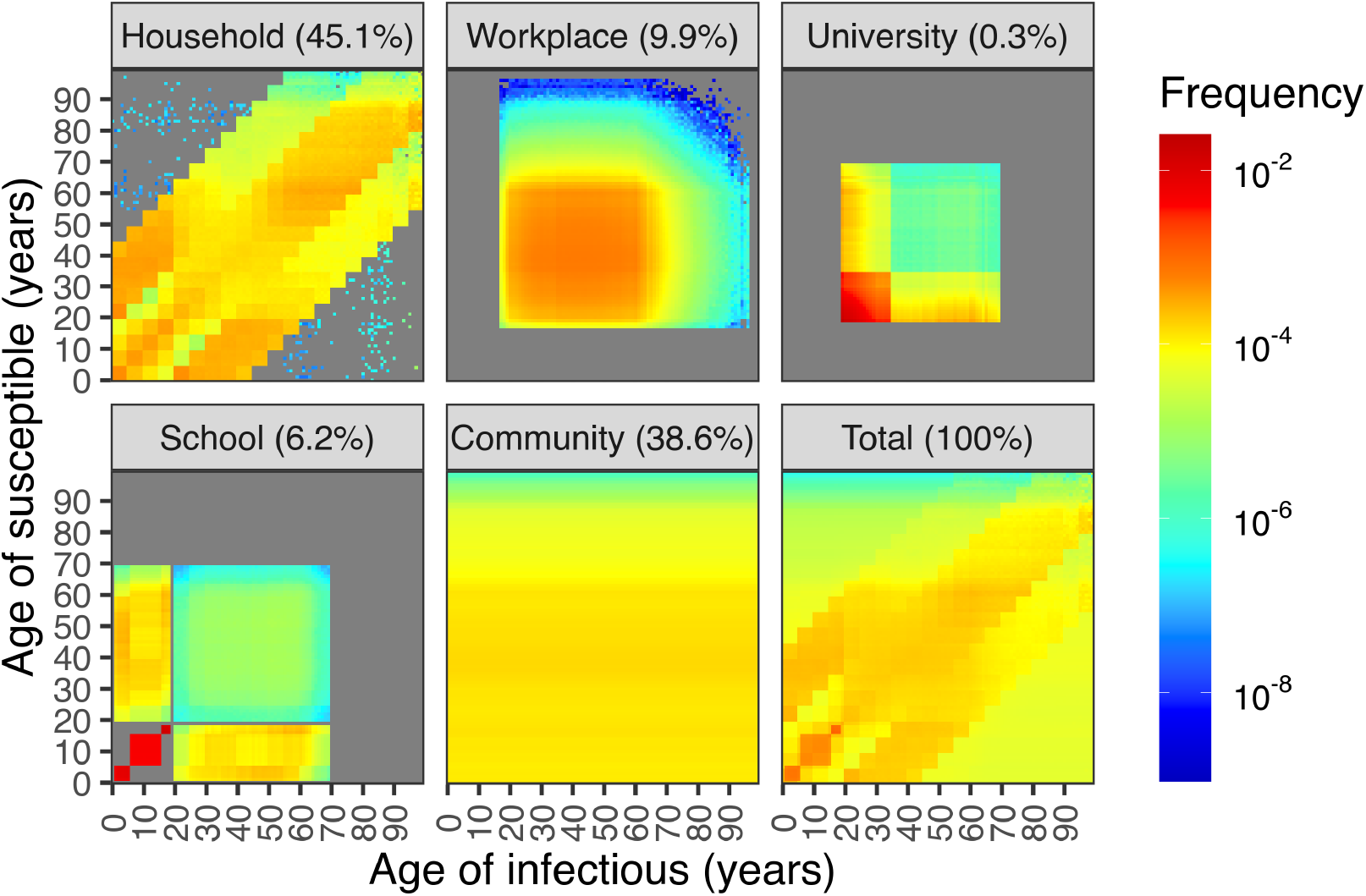
The age-specific contact pattern in the IBM. The age-specific contact frequencies of each transmission setting and total sum across all settings from low to high are represented by a color gradient from blue to red. The total frequency is obtained by the linear combination of each transmission setting, with the corresponding contributing proportion shown in brackets. These contact patterns are similar to those in the modeling study in the UK [91].

Additionally, we account for individual mobility, where contact patterns can vary by distance. We obtained mobility data for each municipality in Norway from Telenor in 2021 [44]. These data reveal that long-range movements, such as travel from Oslo to Bergen, occur less frequently than short-range movements, such as travel between different districts in Oslo. Fig S2B shows the fitted distribution of mobility distances based on the proportion of radius of gyration for the top five most populated municipalities in Norway (Oslo, Bergen, Trondheim, Stavanger, Bærum).

##### S1.1.4 Epidemiological status

Each individual in the model is assigned an epidemiological status according to a SEIR-like epidemiological model, which is illustrated in gray in Fig S3. A susceptible individual can become infected by coming into contact with asymptomatic, pre-symptomatic, or symptomatic infectious individuals. Once infected, the individual becomes exposed, but is not yet infectious, and may develop either asymptomatic or symptomatic illness in the next few days. The probability of remaining asymptomatic is 40%, while the probability of developing pre-symptomatic illness before becoming symptomatic is 60%. Symptomatic individuals may either recover or die, as shown in red and blue in Fig S3, respectively. Asymptomatic individuals who become infected are assumed to recover without developing symptoms.

The time spent of each status following exposure is represented by a gamma distribution and is age-independent. Table S1 shows the parameters for each status. The median generation time, defined as the time between the infection in a primary case and a secondary case, is approximately 7 days. An example of the generation time distribution from one simulation is shown in Fig S2C.

**Fig S2.**
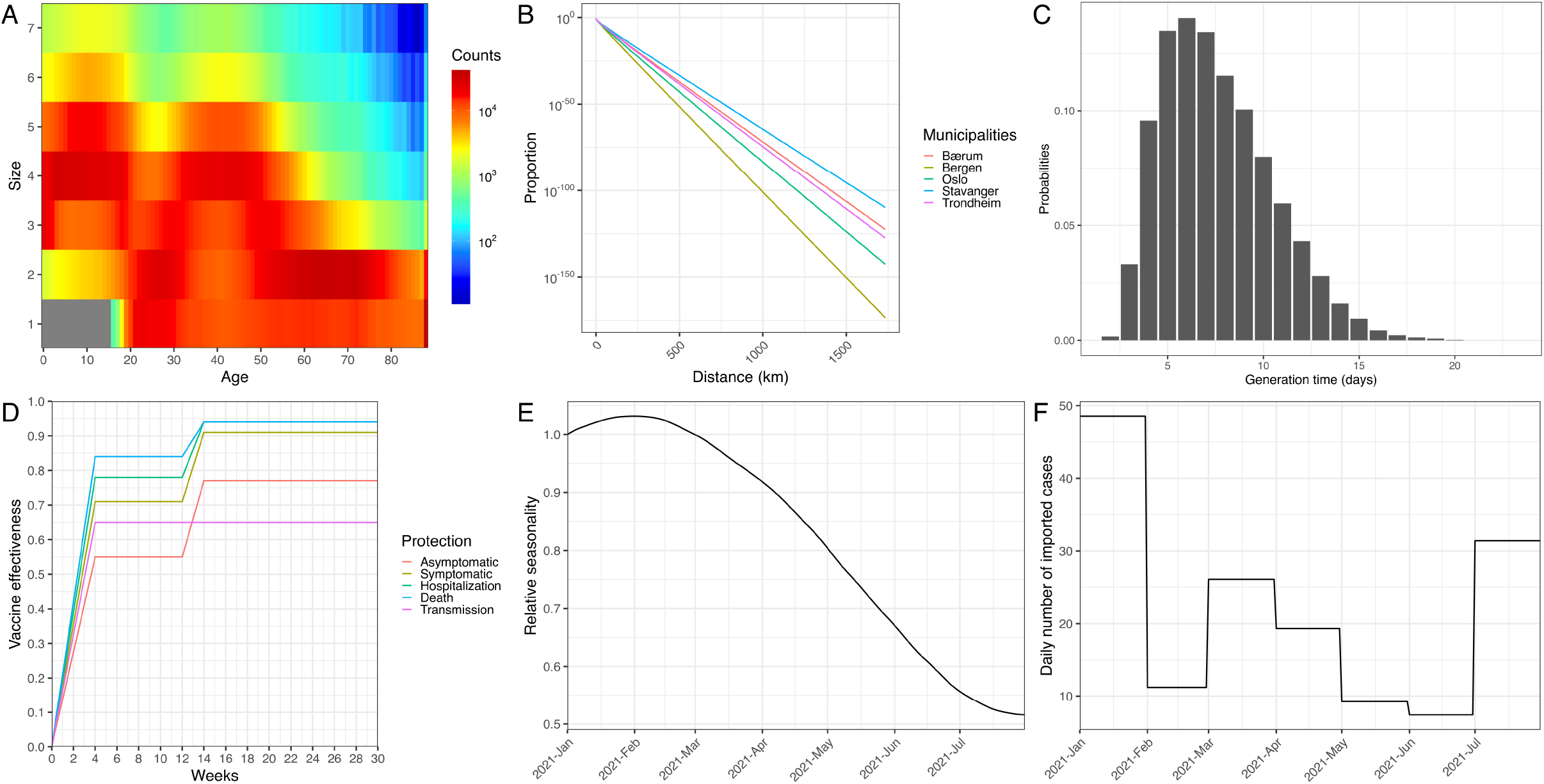
The model assumptions. A: Age-specific distribution of household sizes in Norway. The gray color represents that those 15 years old and younger are not single living. The rightmost column shows the group of individuals 88 years old or older, who are primarily living alone. Household counts are shown on a log-scale. B: The municipality-specific mobility pattern in Norway. The proportions of radius of gyration of the top 5 most populated municipalities (Oslo, Bergen, Trondheim, Stavanger, Bærum) are shown in a log-scale. C: The generation time distribution. Probability mass function (PMF) of the generation time distribution of the individual-based model is illustrated using results from one simulation. The median generation time is 7 days. D: The growth of vaccine effectiveness. The effectiveness of the first dose increases linearly from zero to the first full effect in 28 days (4 weeks) and then remains constant for 56 days (8 weeks) after reaching the first full effect. The time interval between first and second doses is assumed to be 84 days (12 weeks). The effectiveness of the second dose increases linearly to the second full effect in 14 days (2 weeks) after vaccination. E: The seasonality of transmission rate. The relative seasonality of transmissibility starts from 1.0 on the 1st of January and varies according yearly temperature data in Norway. The 50% seasonality refers to a 50% decrease in the lowest transmissibility during summer compared to the highest in winter. F: The importation of cases. The daily imported cases are average values based on monthly national data from MSIS. The observed monthly cases from January to July 2021 were 1505, 313, 809, 579, 285, 221, 973. These cases were then distributed among different counties and age groups according to empirical distributions.

The probability of death given a symptomatic infection is age-specific and depends on whether an individual belongs to a risk group, i.e. risk status. These values are calibrated using Norwegian registry data, which is explained in the following section.

**Fig S3.**
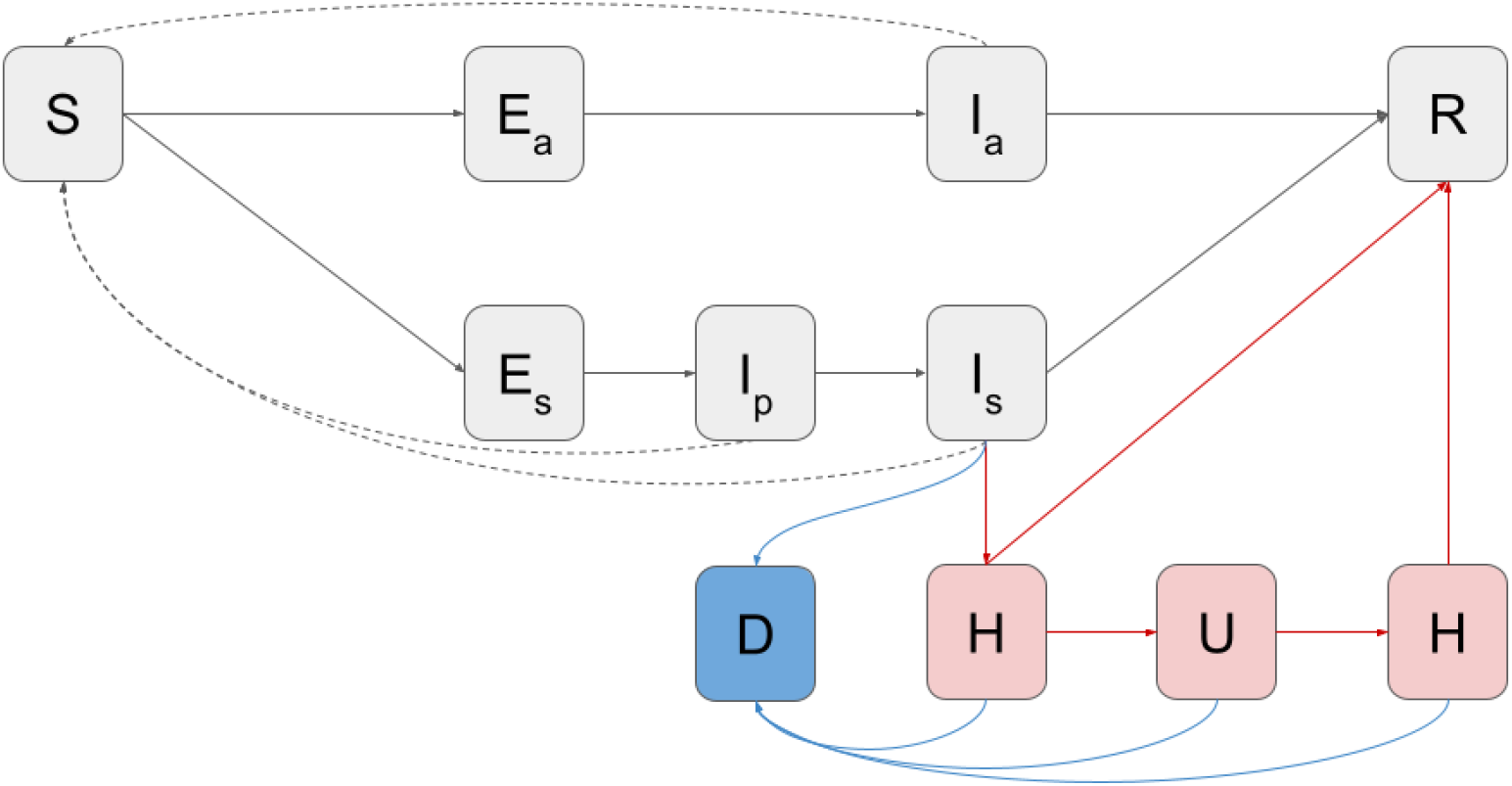
The epidemiological and hospitalization model. Susceptible individuals (*S*) who have not been exposed to the virus or the vaccine, upon exposure, enter a latent state (*E*_*a*_ or *E*_*s*_) before becoming infectious. Symptomatic individuals enter a pre-symptomatic state (*I*_*p*_) before developing symptoms (*I*_*s*_). Asymptomatic infections (*I*_*a*_) are assumed to recover (*R*) without developing symptoms. Those with symptomatic infections (*I*_*s*_) may recover (*R*) or require hospitalization (*H*) due to severe illness. Hospitalization can lead to recovery (*R*), or admission to the ICU (*U*) followed by a second stay in the hospital ward (*H*), and eventual recovery (*R*). Deaths (*D*) occur from the symptomatic state or during hospitalization. The epidemiological statuses are shown in gray. *S*: susceptible. *E*_*a*_: asymptomatic exposed. *I*_*a*_: asymptomatic infectious. *E*_*s*_: symptomatic exposed. *I*_*p*_: pre-symptomatic infectious. *I*_*s*_: symptomatic infectious. *R*: recovered. The solid directed lines are transitions between statuses and the dashed directed lines represent infectious transmissions. The symptomatic infectious people may be hospitalized shown in red statuses and/or die shown in blue as extra layers. *H*: hospitalized. *U* : ICU admission. *D*: died.

**Table S1.** The time spent and infectiousness of compartments in the transmission model.

| Compartments | Mean (days) | Shape | Scale | Infectiousness |
| --- | --- | --- | --- | --- |
| Presymptomatic latent ($E_s$) | 3.0 | 7.5 | 0.4 | 0 |
| Asymptomatic latent ($E_a$) | 3.0 | 7.5 | 0.4 | 0 |
| Presymptomatic ($I_p$) | 2.0 | 5.0 | 0.4 | 1.25 |
| Symptomatic ($I_s$) | 4.8 | 6.0 | 0.8 | 1.00 |
| Asymptomatic ($I_a$) | 4.8 | 6.0 | 0.8 | 0.10 |

Individuals who are exposed to the virus enter a latent period during which they are not infectious. After this period, 40% of them may become asymptomatic, which means they are less infectious than those who develop symptoms. The table provides information about the gamma distribution of the time spent in each status, including the shape and scale parameters, as well as the corresponding mean values.

##### S1.1.5 Hospitalization status

We account for the possibility that individuals with symptomatic infections may develop severe illness and require hospitalization or ICU admission. While some hospitalized individuals may not require ICU admission and can recover and be discharged after a few days, others may need to stay in the hospital longer, especially those who require ICU admission. Age is the only factor that influences the time spent in hospital or ICU.

The time spent in each status, including the time from symptom onset to hospitalization, follows negative binomial distributions with their probabilities and size parameters. We first model the time from symptom onset to hospitalization and then split it into two ways to model individuals requiring ICU admission and those who do not. For those who do not require ICU admission, the length of stay in the hospital is captured by one distribution. For those who require ICU admission, we model the time from hospitalization to ICU admission, length of stay in ICU, and length of stay in the hospital after ICU admission in three separate steps.

The probabilities of hospitalization given symptomatic infection are age-specific and depend on whether individuals belong to a risk group. These probabilities are calibrated using Norwegian registry data, which we present in the next section.

##### S1.1.6 Vaccination status

Each individual can be vaccinated on a specific date. However, after vaccination, it takes some time for immunity to develop, and protection increases gradually over the following weeks. In this study, we assume a leaky type of vaccine effectiveness instead of an all-or-nothing effect [16]. This means that while all vaccinated people are not fully protected, their risk of infection is reduced by a certain fraction in each contact. Additionally, we assume no waning immunity during the simulation period due to its relatively short duration.

Our assumptions for vaccine effectiveness are as follows. The protection probability increases linearly from zero to the first full effect during the first 28 days (4 weeks) after vaccination. The effectiveness then remains constant for the following 56 days (8 weeks), as the time interval between the first and second doses is assumed to be 84 days (12 weeks). We assume that everyone who takes the first dose will take the second, and that the effectiveness of the second dose increases linearly to the second full effect over the following 14 days (2 weeks).

The vaccine provides several protections against (i) symptomatic infection, (ii) asymptomatic infection, (iii) hospitalization, (iv) death and (v) transmissibility. The full protection are (i) 71/91%, (ii) 55/77%, (iii) 78/94%, (iv) 84/94% and (v) 65/65% after the first/second dose, respectively. The values are based on the recommendation reports by the Norwegian Institute of Public Health (NIPH) [32–35] and are similar to estimates from another study [53]. Table S2 and Fig S2D show the vaccine effectiveness and its growth over time.

The conditional effectiveness against severe illness (i.e. hospitalization or death) given symptomatic infection can be calculated by Eq (S1).

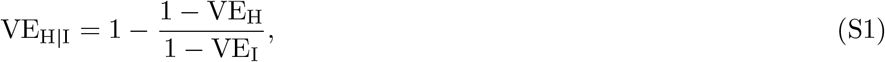

where VE_H|I_, VE_H_, VE_I_ are the vaccine effectiveness against severe illness given symptomatic infection, severe illness and symptomatic infection, respectively. For example assuming vaccine reaches the maximum effectiveness (i.e. more than 2 weeks after the second dose), we have VE_I_ = 0.91, VE_H_ = 0.94 and VE_H|I_ = 0.33, which means that the risk of severe illness given symptomatic infection is reduced by 33%.

##### S1.1.7 Seasonality

Apart from control interventions, the transmission rate is known to vary seasonally, with higher rates occurring in the winter and lower rates in the summer. We assume a 50% relative difference between the highest and lowest seasonal rates, which is defined based on the temperature over a year. Fig S2E illustrates the seasonal variation in the transmission rate from January to July.

**Table S2.** The vaccine effectiveness against different health statuses.

| Vaccine effectiveness against | 1st dose (%) | 2nd dose (%) |
| --- | --- | --- |
| Symptomatic | 71 | 91 |
| Asymptomatic | 55 | 77 |
| Hospitalization | 78 | 94 |
| Death | 84 | 94 |
| Transmissibility | 65 | 65 |

##### S1.1.8 Importation

Importation of infectious cases is simulated by converting susceptible individuals to either symptomatic or asymptomatic infected individuals. We select individuals based on age- and location-specific empirical distributions derived from observed data. At each time step (each day) in the simulation, a daily importation number is drawn from a Poisson distribution with a time-varying mean, which is based on the data presented in Fig S2F.

##### S1.1.9 Calibration

We calibrated the ratios between the four setting-specific betas (household, school, workplace, and community) using COVID-19 test positive data. The main contributions to the positive cases were infections within households and in the community, which includes all routes other than households, schools, or workplaces.

To account for time-specific changes, we divided the seven-month simulation period into three intervals, separated by two change points on the 28th of January and 11th of March 2021. We selected the change points based on the local minimum and maximum rolling averages of daily hospital admissions and shifted them back 12 days to account for the average delay between infection and hospitalization.

To calibrate the IBM, we used 12 free parameters, including beta (i.e. transmission rates) during three time periods and susceptibilities of nine age groups to capture time-specific changes and age-specific differences, respectively. We sampled 100,000 parameters using the Latin hypercube sampling (LHS) approach and selected the best 10 sets based on the least squares method. The least squares error was calculated using a data set consisting of two marginal distributions: *N*_*t*_ = 212 time points of daily hospital admissions and the seven-month cumulative admissions of *N*_*a*_ = 9 age groups. The least squares error is

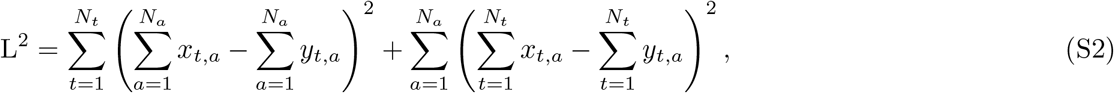

where *x*_*t,a*_ and *y*_*t,a*_ are the daily hospital admission of model output and observed data at time *t* and of age group *a*, respectively. The marginals 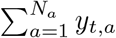 and 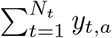 are the daily hospital admissions of all ages at time *t* and the 7-month total hospital admissions of each age group *a*, respectively.

To narrow down the range of parameters ranging from 0 to 1, we performed several rounds of Latin hypercube sampling (LHS). Fig S4 shows the final round parameter distributions, with higher beta values in February corresponding to the growth of cases in March.

Our estimated susceptibilities to infection showed a decrease with age, except for the 50-59 age group. This is in contrast to findings in other modeling studies [79–81] and cohort studies [82, 83]. Several factors could explain these differences. First, we applied age-specific risk ratios of the Alpha variant in Norway [56], which had large uncertainties and may have overestimated values for younger age groups or underestimated values for older age groups. Therefore, susceptibilities for older individuals may have been underestimated. Second, our age-specific contact patterns were taken from 2017 before the pandemic, while contact matrices in other modeling studies [80, 81] were separated into pre-pandemic and post-lockdown periods. It is possible that older individuals reduced their contacts more than younger individuals during the pandemic, leading to underestimated susceptibilities for older age groups. Third, vaccine effectiveness may have been lower for older individuals [93], while we assumed equal effectiveness across all age groups, which may have overvalued vaccine effectiveness for older individuals and led to underestimated susceptibilities for this group. The national recommendation reports by NIPH addressed this issue by splitting vaccine effectiveness into two categories: those aged 65 years and younger and those older than 65 years [94–97].

Fig S5 shows the fits of the model to the data. The estimated numbers of infections are higher than the reported cases due to under-detection, particularly in younger age groups. The calibration process uses only daily and age-specific hospital admissions to estimate the 12 free parameters. The good match between the observed and modeled ICU admissions and deaths in Fig S5A validates the calibration method. Fig S5C, which shows the geographic-specific differences not used in the estimation, also supports the model accuracy.

Using the observed number of deaths and the estimated mean number of infections, we calculate the age-specific probabilities of death given infection. The age-specific probabilities of ICU admission given hospitalization are calculated based on the observed numbers of ICU admissions and hospitalizations. The resulting age-specific probabilities of death and ICU admission are presented in Table S3. Note that these probabilities are calculated in the absence of vaccination and that the vaccine effectiveness on the probabilities depends on the timing of vaccination for each individual.

**Table S3.** The age-specific probabilities of death given infection and ICU admission given hospitalization in the IBM.

| Age groups | Probabilities of death (%) |  | Probabilities of ICU admission (%) |
| --- | --- | --- | --- |
|  | Without risk factors | With risk factors | All population |
| 0-9 | 0.00009 | 0.0002 | 33.3 |
| 10-19 | 0.0004 | 0.0007 | 6.9 |
| 20-29 | 0.007 | 0.01 | 12.2 |
| 30-39 | 0.008 | 0.01 | 9.9 |
| 40-49 | 0.04 | 0.07 | 13.1 |
| 50-59 | 0.08 | 0.1 | 20.8 |
| 60-69 | 0.6 | 1.06 | 25.5 |
| 70-79 | 2.9 | 5.4 | 28.5 |
| 80+ | 13.0 | 23.9 | 17.1 |

We assume that the probabilities of death depend on both age and risk status while the probabilities of ICU admission depend on age only. Based on the observed number of deaths and the estimated mean number of infections, we calculate the age-specific probabilities of death given infection. Similarly, based on the observed numbers of ICU admissions and hospitalizations, we calculate the age-specific probabilities of ICU admission given hospitalization.

To assess the sensitivity of the results to the choice of parameters, we also calibrate the model using the probabilities of hospitalization without the factor of 0.5. In this case, the estimated number of infections is roughly halved, and the resulting probabilities of death given infection are approximately doubled.

**Fig S4.**
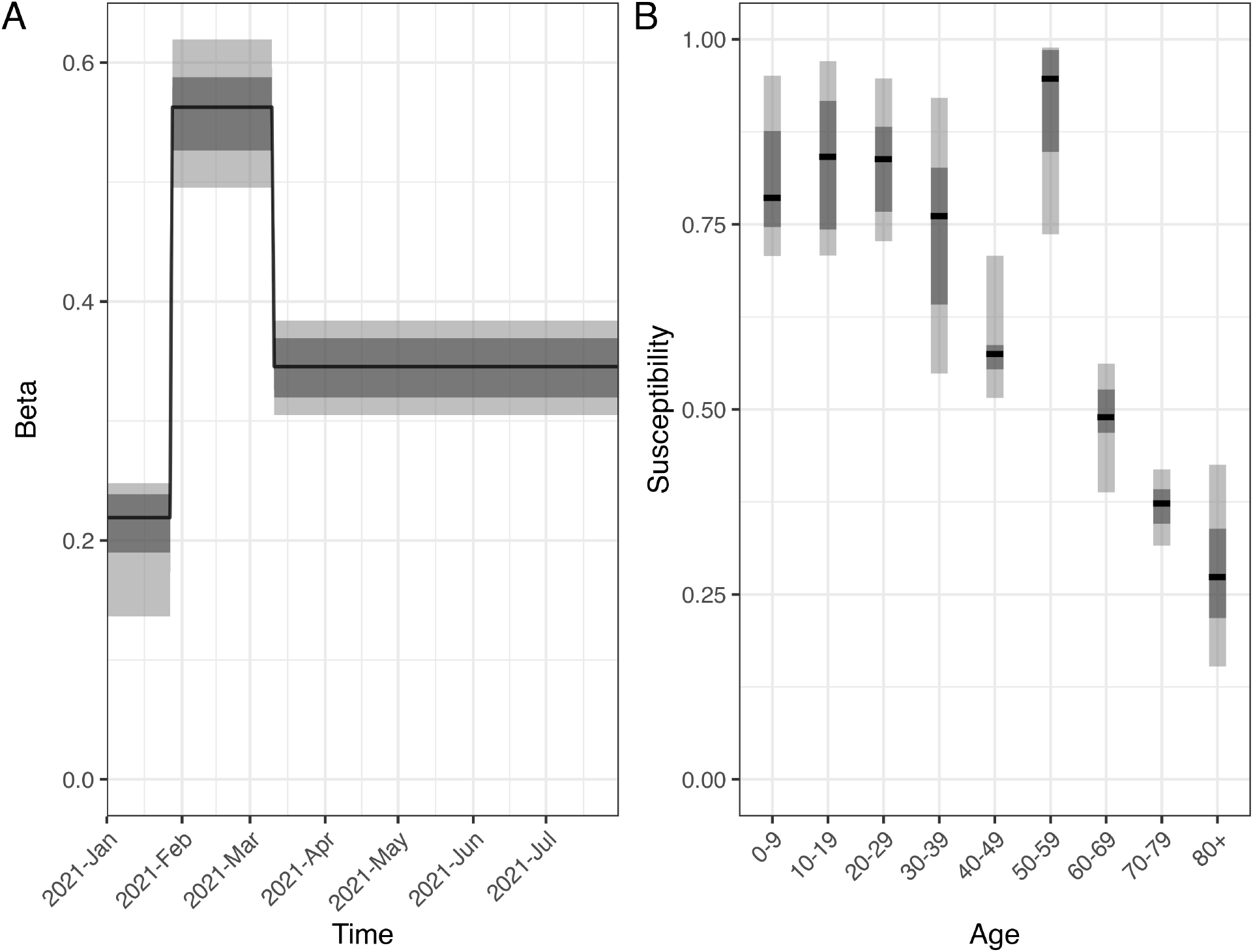
The marginal distributions of 12 estimated parameters in the IBM. (A) Beta in 3 time periods and (B) susceptibilities of 9 age groups are sampled from Latin hypercube sampling (LHS) approach. The black lines show the median values, and the darker (lighter) gray areas show the ranges of 25th and 75th percentiles, i.e. lower and upper quantiles (2.5th and 97.5th percentiles) of the estimated parameters.

#### S1.2 Meta-population model (MPM)

The meta-population model (MPM) is based on an epidemiological model identical to the one used in the individual-based model (IBM). The model incorporates meta-populations that are characterized by age, risk factor, and geography. The geographic regions are based on the five regions of Norway (North, West, Mid, East, and South) and different prioritization schemes. This results in a representation of every possible combination of prioritization (*Plus, Neutral*, or *Minus*) and region. In the baseline model, we have 10 geographic regions, 9 age groups, and 2 risk profiles, resulting in 180 separate meta-populations. Vaccination is incorporated as an additional layer of meta-populations, resulting in 540 meta-populations in the baseline when considering 3 vaccination states (No vaccine, first dose, and second dose). The vaccinated groups exhibit different values for susceptibility, severity, and transmissibility compared to the non-vaccinated groups.

**Fig S5.**
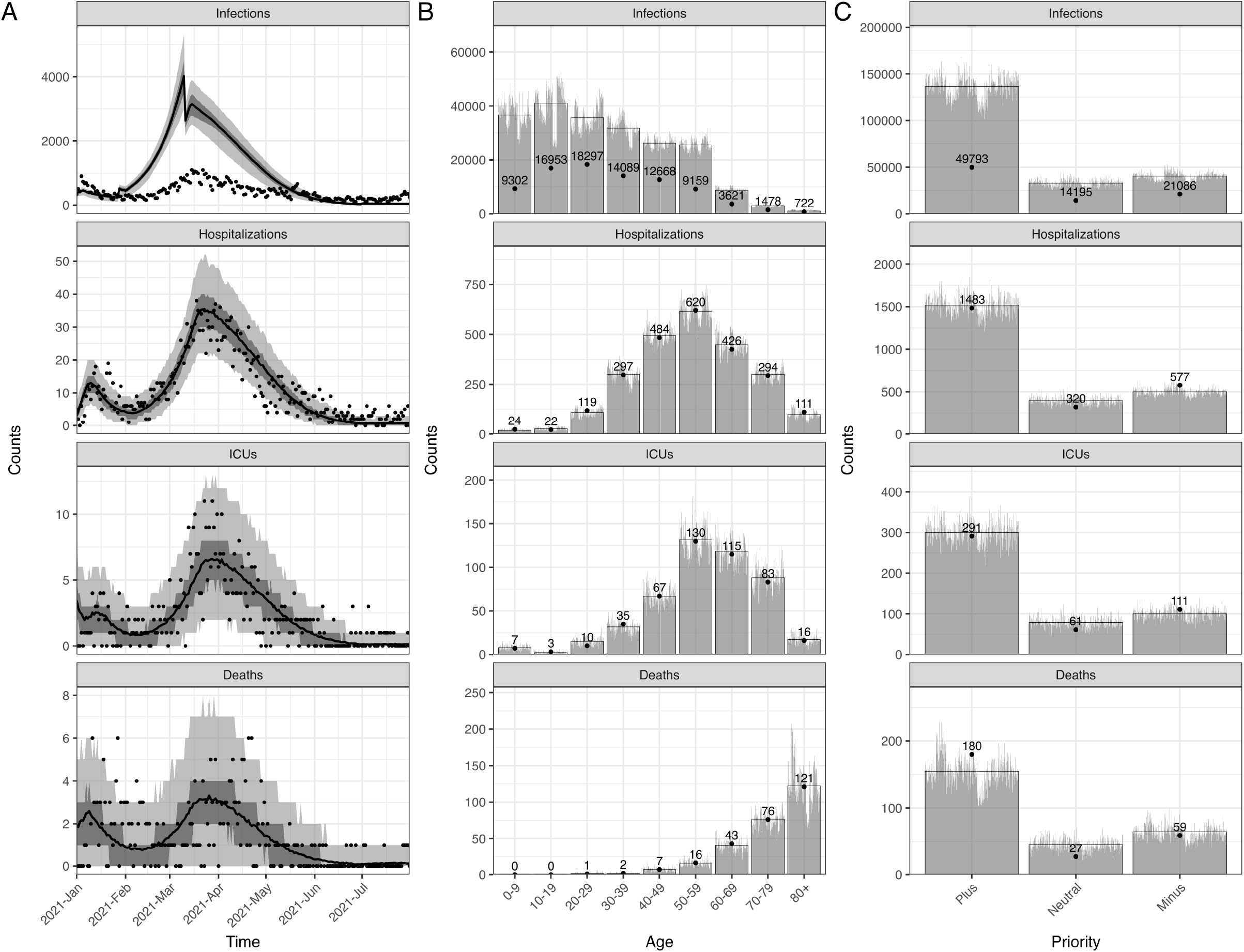
The calibrated IBM and data. A: The time series data of all ages. The lines show the model fits with their 50 and 95% prediction intervals represented by gray areas. The dots show the observed data. B: The age distribution of total counts. C: The geographic-specific priority distribution of total counts. The gray bars show the counts of each simulation and the full bars with black borders show the mean of all simulations. The data are shown in dots with their exact numbers.

The model is defined by the following equations (see Table S4 for description of variables):

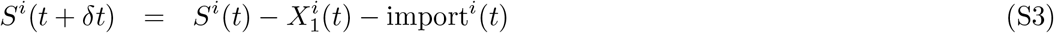

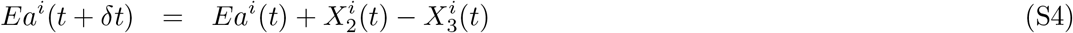

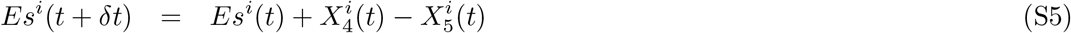

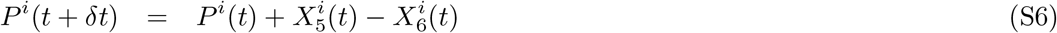

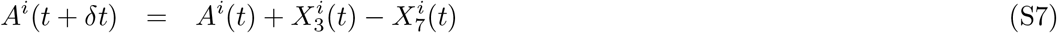

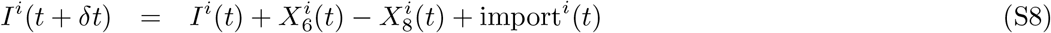

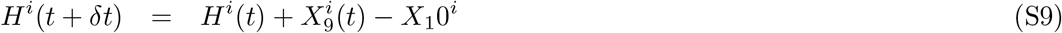

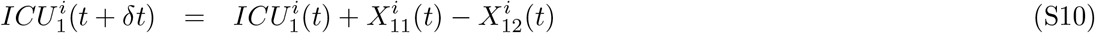

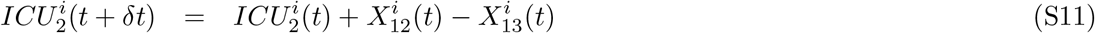

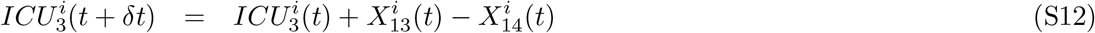

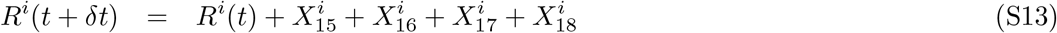

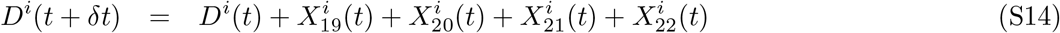

The infection process, given by 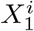 is simulated from a binomial distribution with a probability given by the force of infection, Λ_*i*_

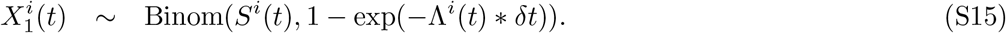

The force of infection is

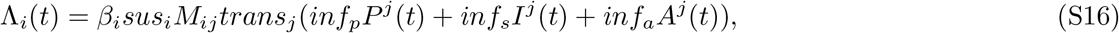

where 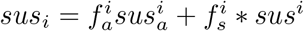. The contact matrix *M*_*ij*_ is the outer product of an age-based contact matrix based on a survey in Norway and regional contact matrix. We assume the people in risk groups have the same contact pattern as the people not in risk groups. Fig S6 shows the age-based contact matrix.

The other transition probabilities *X*_*i*_ are determined by the appropriate probabilities and lengths of stay in various compartments.

**Fig S6.**
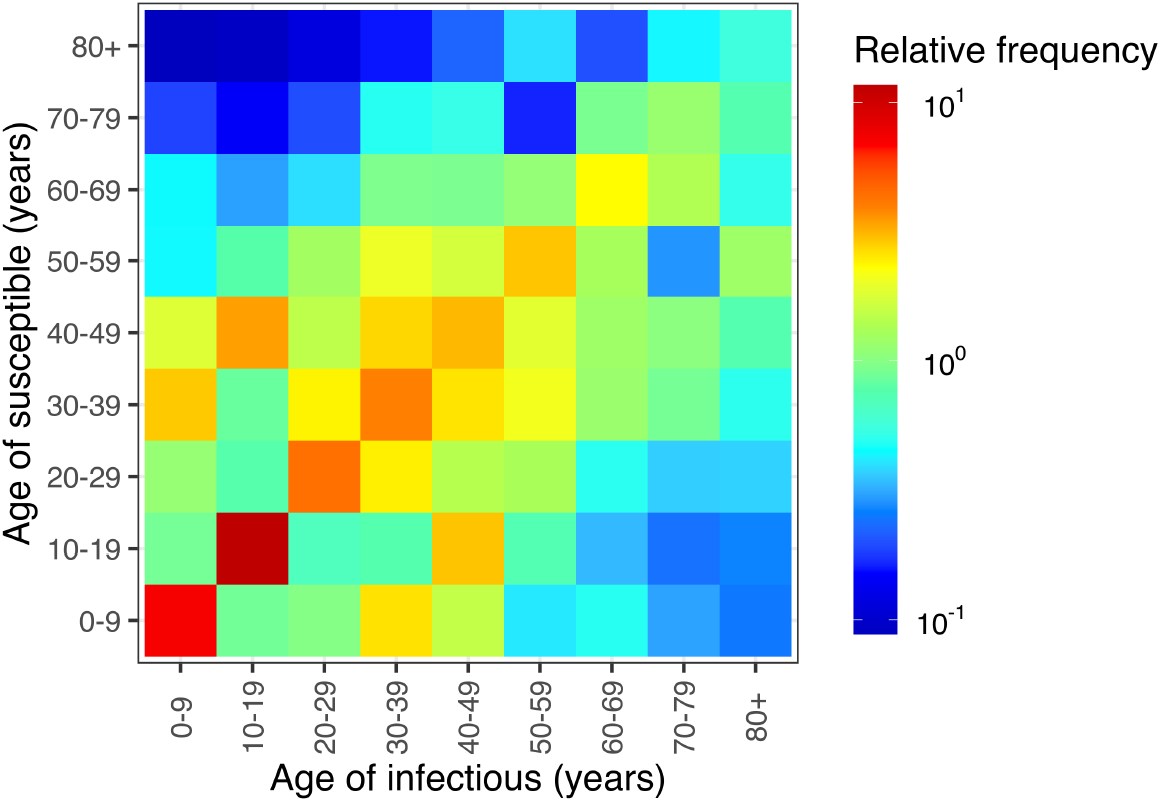
The age-specific contact pattern in the MPM. The age-specific relative contact frequencies based on a survey in Norway from low to high in log10-scale are represented by color from blue to red.

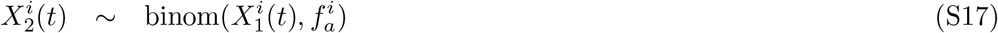

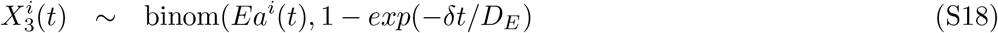

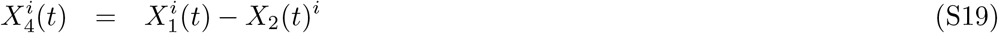

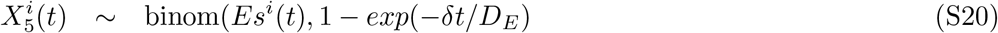

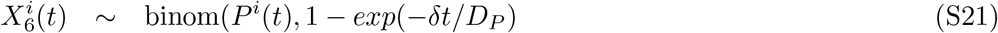

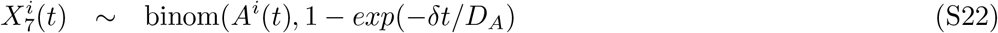

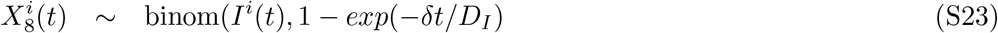

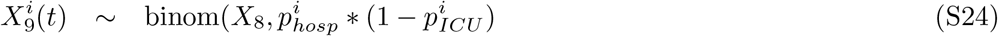

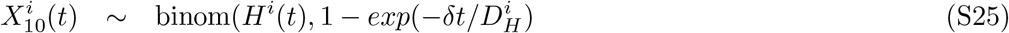

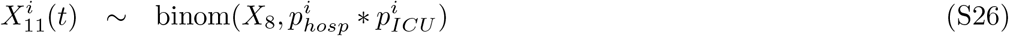

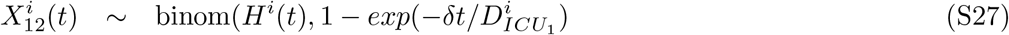

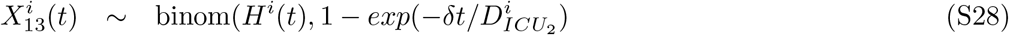

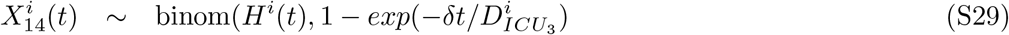

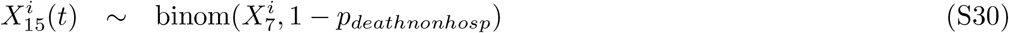

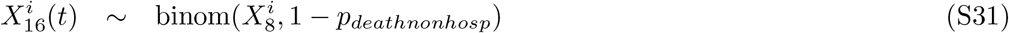

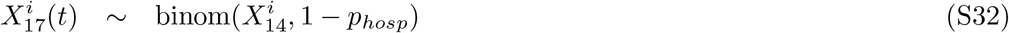

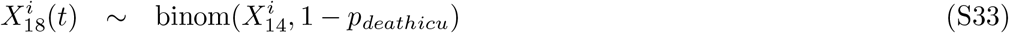

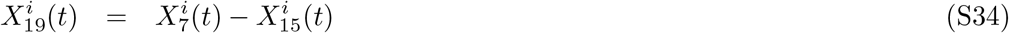

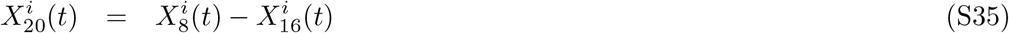

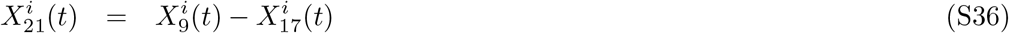

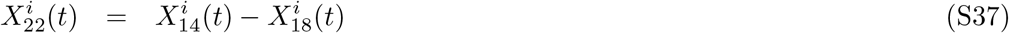

**Table S4.** Compartments in the meta-population model.

| Variable | Description |
| --- | --- |
| $S$ | Susceptible |
| $Ea$ | Exposed, will become asymptomatic |
| $Es$ | Exposed, will become symptomatic |
| $P$ | Presymptomatic infectious |
| $A$ | Asymptomatic infectious |
| $I$ | Symptomatic infectious |
| $H$ | Hospitalized |
| $ICU_1$ | In hospital will go to ICU |
| $ICU_2$ | In ICU |
| $ICU_3$ | Post-ICU care |
| $R$ | Recovered and immune |
| $D$ | Dead |

##### S1.2.1 Model parameters

The main model parameters are described in Tables S5 and S6. The probabilities of hospitalization, ICU admission and death are further adjusted by risk group and vaccination. The probabilities of hospitalization for the model can be found in Table S8. Risk groups are discussed in Section SS2.3.

**Table S5.** Parameters in the meta-population model.

| Variable | Description | Value | Source |
| --- | --- | --- | --- |
| $\beta^i(t)$ | Overall transmissibility | Fitted | |
| $inf_s$ | Symptomatic infectiousness | 1 | Baseline |
| $inf_a$ | Asymptomatic infectiousness | 0.1 | Feretti et al. [98] |
| $inf_p$ | Presymptomatic infectiousness | 1.25 | Modified Feretti et al. [98] |
| $D_E$ | Latent period | 3 days | |
| $D_P$ | Presymptomatic period | 2 days | |
| $D_A$ | Duration asymptomatic | 4.8 days | |
| $D_I$ | Duration symptomatic | 4.8 days | |

**Table S6.** Age varying parameters in the meta-population model.

| Variable | Description | 0-9 | 10-19 | 20-29 | 30-39 | 40-49 | 50-59 | 60-69 | 70-79 | 80+ |
| --- | --- | --- | --- | --- | --- | --- | --- | --- | --- | --- |
| $sus_i$ | Symptomatic Susceptibility | Fitted | | | | | | | | |
| $sus_a$ | Asymptomatic susceptibility | Fitted | | | | | | | | |
| $f_a$ | Fraction asymptomatic | 0.4 | 0.4 | 0.4 | 0.4 | 0.4 | 0.4 | 0.4 | 0.4 | 0.4 |
| $f_s$ | Fraction symptomatic | 0.6 | 0.6 | 0.6 | 0.6 | 0.6 | 0.6 | 0.6 | 0.6 | 0.6 |
| $D_H$ | Length of stay in Hospital | 1.8 | 3.5 | 3.6 | 3.4 | 4.7 | 5.5 | 6.4 | 6.2 | 6.4 |
| $D_{ICU_1}$ | Length of stay in Hospital pre-ICU | 1.4 | 1.4 | 1.4 | 2.8 | 3.7 | 3.1 | 3.9 | 4.5 | 4.5 |
| $D_{ICU_2}$ | Length of stay on ICU | 3.3 | 3.3 | 3.3 | 3.3 | 9.3 | 9.5 | 16 | 15 | 14 |
| $D_{ICU_3}$ | Length of stay in hospital post-ICU | 5 | 5 | 5 | 8.7 | 8.6 | 8.3 | 11 | 11 | 11 |
| $picu$ | Risk of ICU given hospitalization | Fitted | | | | | | | | |
| $p_{deathnonhosp}$ | Base Probability of death | Fitted | | | | | | | | |

##### S1.2.2 Regional contact matrix

The regional contact matrix is based on mobility data from Telenor [44] aggregated to the number of movements between the specified regions. We define the contact matrix such that amount of infectiousness that “leaks” out of a region is equal to the fraction of the population who traveled out of the region in one day. If the number of of travelers from region *i* to region *j* is *T*_*ij*_ and the population in region *i* is *P*_*i*_, then the regional contact matrix is given by:

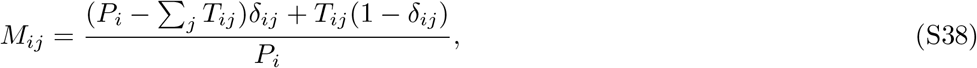

where *δ*_*ij*_ is the Kronecker delta symbol such that *δ*_*ij*_ = 1 for *i* = *j* and zero otherwise.

##### S1.2.3 Vaccination

As in the IBM, the vaccine effects for the different doses are given in Table S2. In the MPM, we implement the time from receiving the first dose to getting the full effect of this dose as half the ramp-up time used in the IBM which corresponds to 14 days. Then the effect of the second dose is taken into account after the time interval between the doses of 84 days. The vaccination is implemented separately from the equations above as a constant moving of people from one meta-population *i* to another *j* at the appropriate time as determined by the vaccination strategy.

We also assume that the doses are equally distributed among everyone who has not yet been vaccinated including the people in other compartments than *S*, who will not receive any benefits.

##### S1.2.4 Seasonality and importation

Seasonality is implemented in the same way in the MPM model as in the IBM. Importation is implemented as shown in the model specification above by flipping on member of the *S* compartment to the *I* compartment. The amount of importation is identical to the IBM.

##### S1.2.5 Calibration

The calibration of the meta-population model follows the same overall approach as the IBM. We aim to simultaneously calibrate *β*-values and susceptibility values to the hospital incidence over time and to the age distribution of cumulative hospitalizations, shown in Fig S7. The two targets are calibrated at the same time by assuming that the observed cumulative age distribution is constant over time and we define a log-likelihood for a given day *t*:

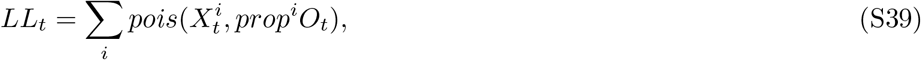

where *pois* is the Poisson distribution, 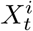 the modeled hospital incidence in age-group *i* on day *t, prop*^*i*^ is the proportion of the cumulative hospitalization that is in age group *i* and *O*_*t*_ is the observed hospital incidence on day *t*.

From this log-likelihood we use a particle filter to better estimate the likelihood for the stochastic model. As for the IBM, we then use a Latin Hypercube Sampling (LHS) strategy for finding the maximum log-likelihood. We fit the 12 parameters by running 15,000 parameter values and estimating the log-likelihood by the particle filter with 15 particles. We choose the 10 parameter values with the highest log-likelihood. Fig S8 shows the distributions of estimated parameters.

We calibrate *β* and susceptibility for the actual strategy. For the other scenarios with a different configuration of municipalities into the regions in the model, we need modify the *β* values somewhat to get exactly the same epidemic trajectory as in the actual strategy. For each of the geographic patterns, we therefore fit a second scale-factor that is used to scale *β* such that the overall number of hospitalizations is on average the same as in the actual strategy when we have a baseline vaccination strategy without geographic prioritization.

**Fig S7.**
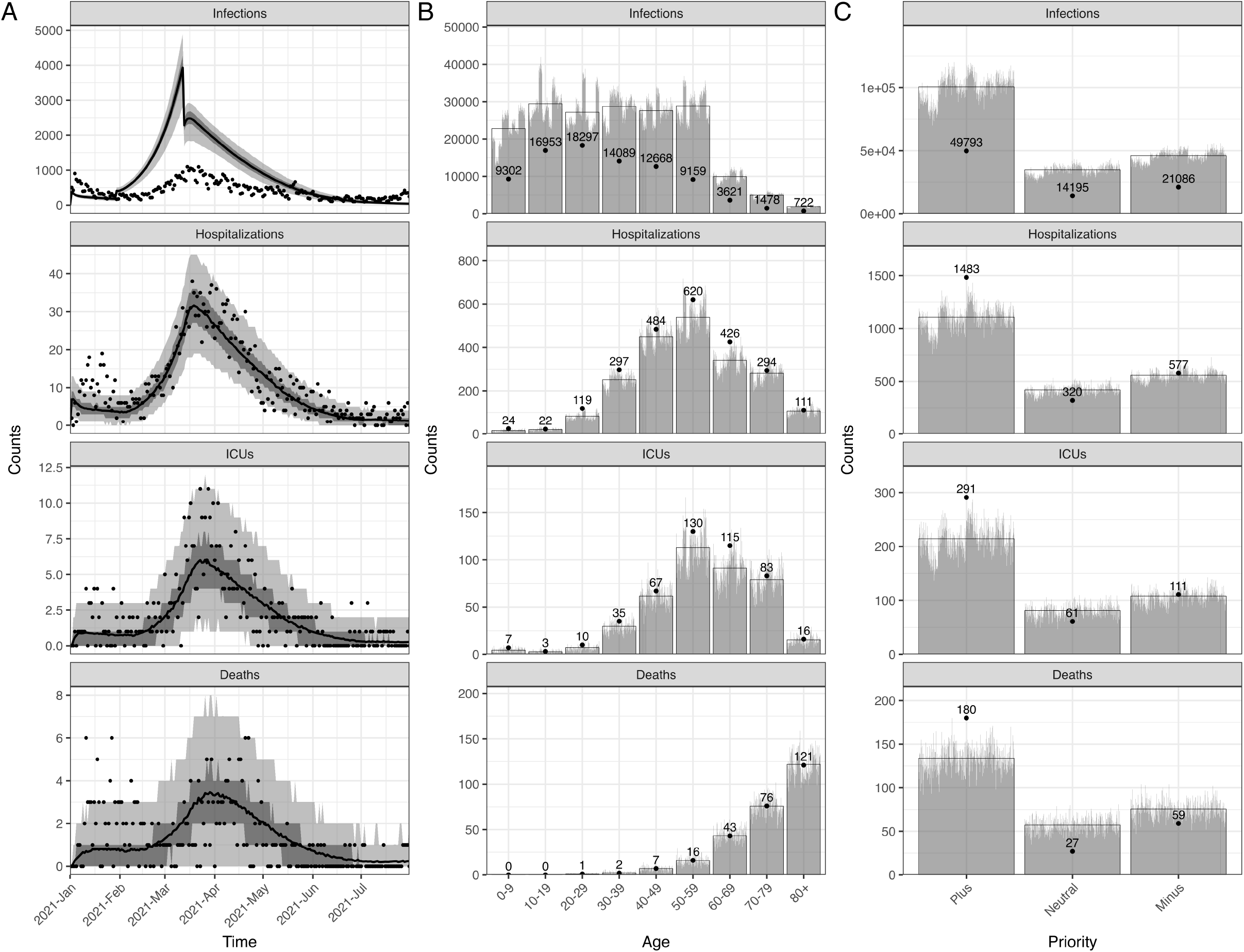
The calibrated MPM and data. A: The time series data of all ages. The lines show the model fits with their 50 and 95% prediction intervals represented by gray areas. The dots show the observed data. B: The age distribution of total counts. C: The geographic-specific priority distribution of total counts. The gray bars show the counts of each simulation and the full bars with black borders show the mean of all simulations. The data are shown in dots with their exact numbers.

**Fig S8.**
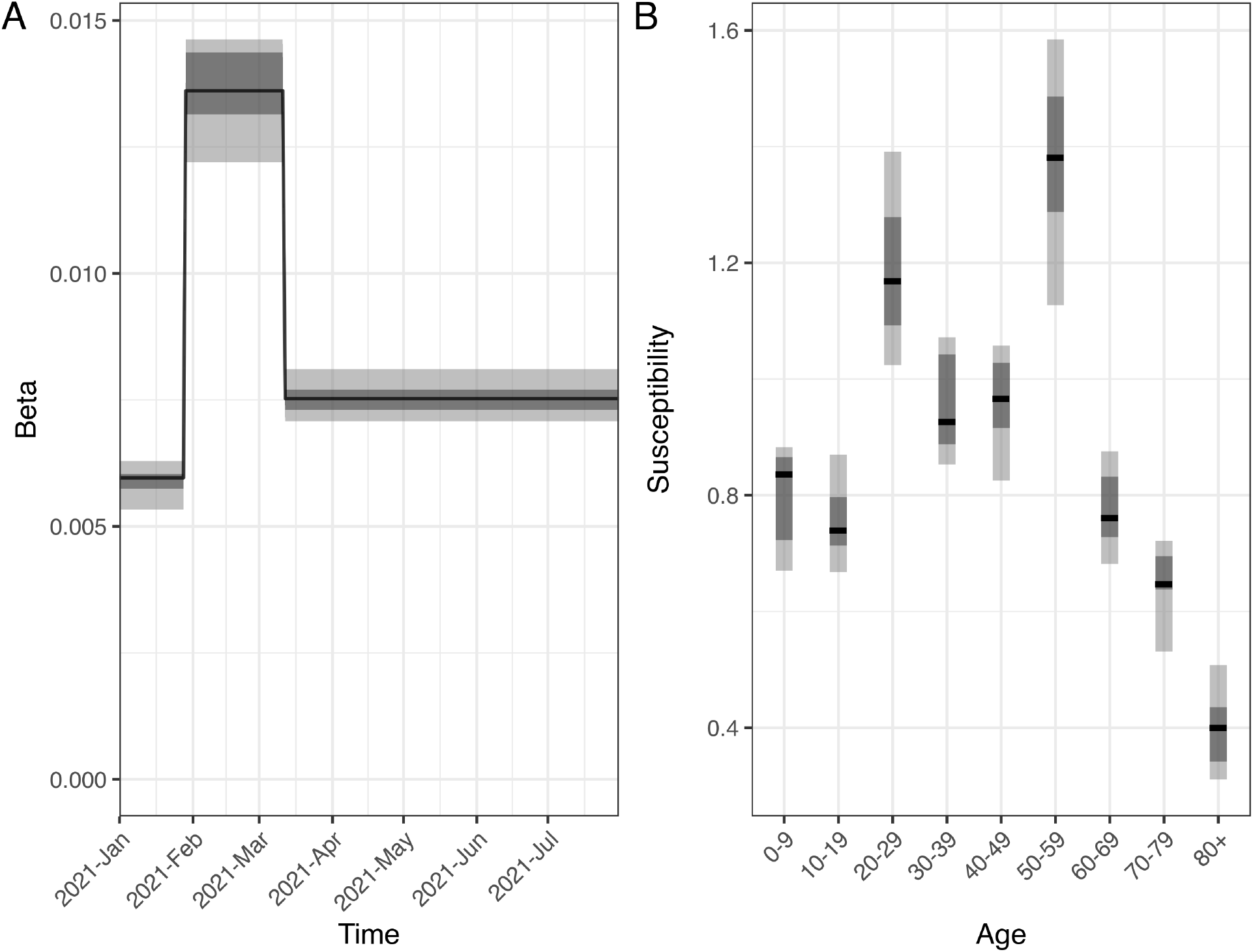
The marginal distributions of 12 estimated parameters in the MPM. (A) Beta in 3 time periods and (B) susceptibilities of 9 age groups are sampled from Latin hypercube sampling (LHS) approach. The black lines show the median values, and the darker (lighter) gray areas show the ranges of 25th and 75th percentiles, i.e. lower and upper quantiles (2.5th and 97.5th percentiles) of the estimated parameters.

The age-specific probabilities of death and ICU admission are calculated following the same approach as the IBM. Table S7 show the age-specific probabilities of death and ICU admission.

### S2 Model assumptions

There are some assumptions shared between 2 models.

#### S2.1 Initial condition

The initial conditions for the two models (IBM and MPM) were taken from the situational awareness model run by the Norwegian Institute of Public Health (NIPH) [43, 50, 51]. In this model, we estimated the numbers in each of the epidemiological compartments for each county in Norway based on calibration to hospital incidence. To get initial conditions on the municipality level, we distributed the county level data by population.

**Table S7.** The age-specific probabilities of death given infection and ICU admission given hospitalization in the MPM.

| Age groups | Probabilities of death (%) |  | Probabilities of ICU admission (%) |
| --- | --- | --- | --- |
|  | Without risk factors | With risk factors | All population |
| 0-9 | 0.00006 | 0.0001 | 29.2 |
| 10-19 | 0.00001 | 0.00002 | 13.6 |
| 20-29 | 0.0008 | 0.001 | 8.4 |
| 30-39 | 0.001 | 0.002 | 11.8 |
| 40-49 | 0.007 | 0.01 | 13.8 |
| 50-59 | 0.03 | 0.05 | 21.0 |
| 60-69 | 0.2 | 0.4 | 27.0 |
| 70-79 | 1.5 | 2.7 | 28.2 |
| 80+ | 9.4 | 17.3 | 14.4 |

We assume that the probabilities of death depend on both age and risk status while the probabilities of ICU admission depend on age only. The age-specific probabilities of death given infection are calculated based on the observed number of deaths and the estimated mean number of infections. The age-specific probabilities of ICU admission given hospitalization are calculated based on the observed numbers of ICU admissions and hospitalizations.

#### S2.2 Relative regional reproduction numbers

It is very difficult to accurately estimate the reproduction number in each municipality due to small number of cases. We therefore estimate and approximate relative reproduction numbers per municipality based on the total number of confirmed cases between February 2020 and May 2021. The aim of the method is to determine what factor we need to scale the estimated national reproduction numbers to get the observed fraction of infected people in each municipality as compared to the national fraction.

Since there are many small municipalities in Norway where a small outbreak can significantly affect the overall fraction of infected, we first do a simple partial-pooling analysis where we fit a simple Bayesian model of the number of observed cases in each municipality, *k*_*i*_ by estimating a fraction, *f*_*i*_ and combining it with the population in the municipality, *p*_*i*_. We use an hierarchical prior on the fractions, *f*_*i*_ to achieve the partial pooling effect.

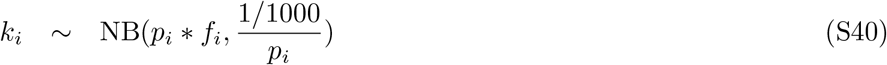

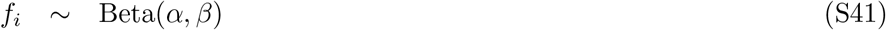

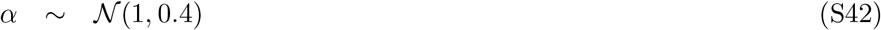

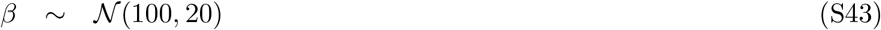

The model was implemented in the Stan language [99] using the rStan interface [100].

Once we have an estimated partially-pooled fraction *f*_*i*_ for each municipality we implemented a simple SEIR model with change points for the beta-parameter with values corresponding to the estimated reproduction number in the NIPH situational awareness model [50, 51]. The SEIR model was a simplified version of the national model described here [43]. The model had symptomatic, presymptomatic and asymptomatic transmission and was seeded by data on imported cases in Norway. We assumed a detection probability of 55%.

For each of the fractions, *f*_*i*_, we found the scale factor *s*_*i*_ which if applied to all the beta-values in the national model would have given a national fraction of infections equal to *f*_*i*_. The reproduction numbers are then normalized to be equal to or less than 1, shown in Fig S9C.

In the IBM, these 356 relative reproduction numbers are used directly, while in the MPM they have to be aggregated since the model uses fewer regions. We use a population-weighted average of the individual scale factor to estimate the relative reproduction number for these combined regions. As mentioned in the MPM section, this aggregation does not perfectly preserve the overall reproduction number, but this is fixed in the calibration step.

#### S2.3 Probabilities of hospitalization

We used the meta-analysis [49] for probabilities of needing hospitalization in each age group as the base values. We increased the probability for those 0-9 years old to the level for those 10-19 years old due to higher observed hospital admissions in Norway. We reduced the probability for those over 80 years old by 25% since this proportion of elderly lived in assisted care homes or nursing homes, and they were generally not admitted to hospital for COVID-19 but treated in the facilities in Norway.

As the main wave during the simulation period (January - July 2021) was contributed by the Alpha variant, we used the age-specific adjusted risk ratio of the Alpha variant compared to wild-type [56] on top of the above probabilities of hospitalization. We noted that the risk ratio could be overestimated compared to other studies in the UK [101, 102], Denmark [103] and some European countries [104], and we thus applied a factor of 0.5 for the probability of hospitalization. In the absence of seroprevalence data, it was difficult to identify the true infection-hospitalization ratio as other studies for example in the Netherlands and Portugal [80, 81]. Here, we included both scenarios of applying the 0.5 factor and without it. The latter was considered as a sensitivity analysis. The corresponding case detection fractions (i.e. the ratios of observed cases from the data and infections from the model) could be low and high, supported by two independent studies (one in January 2021 [105–107] and one in November/December 2020 [108]), respectively.

Furthermore, based on an analysis of Norwegian data from emergency registry Beredt C19 [36], we found that people in risk groups had a 2.7 times higher likelihood of hospitalization and a 1.8 higher likelihood of death [66]. The analysis was based on data until November 2020 and compared the probability of hospitalization and death among those who tested positive based by risk group adjusting for age. Based on the relative sizes of the risk groups, we estimated the risk of hospitalization. Table S8 shows the age-specific probabilities of hospitalization and all the scale factors.

**Table S8.**
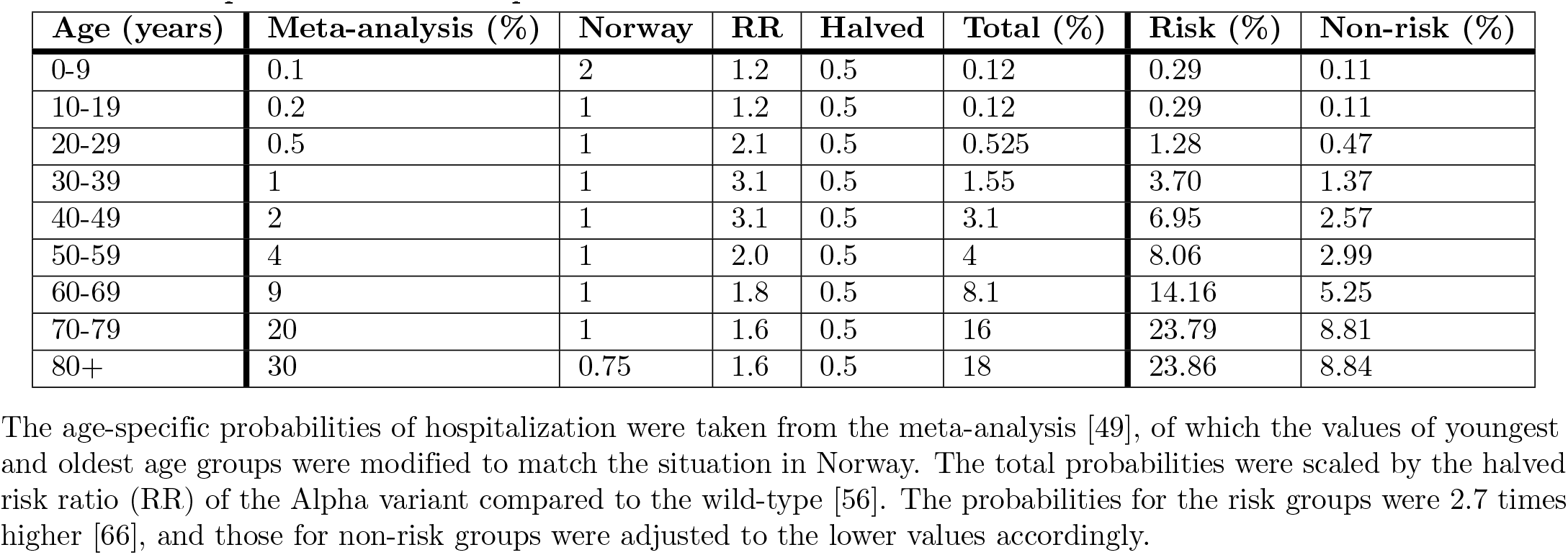
The probabilities of hospitalization.

**Fig S9.**
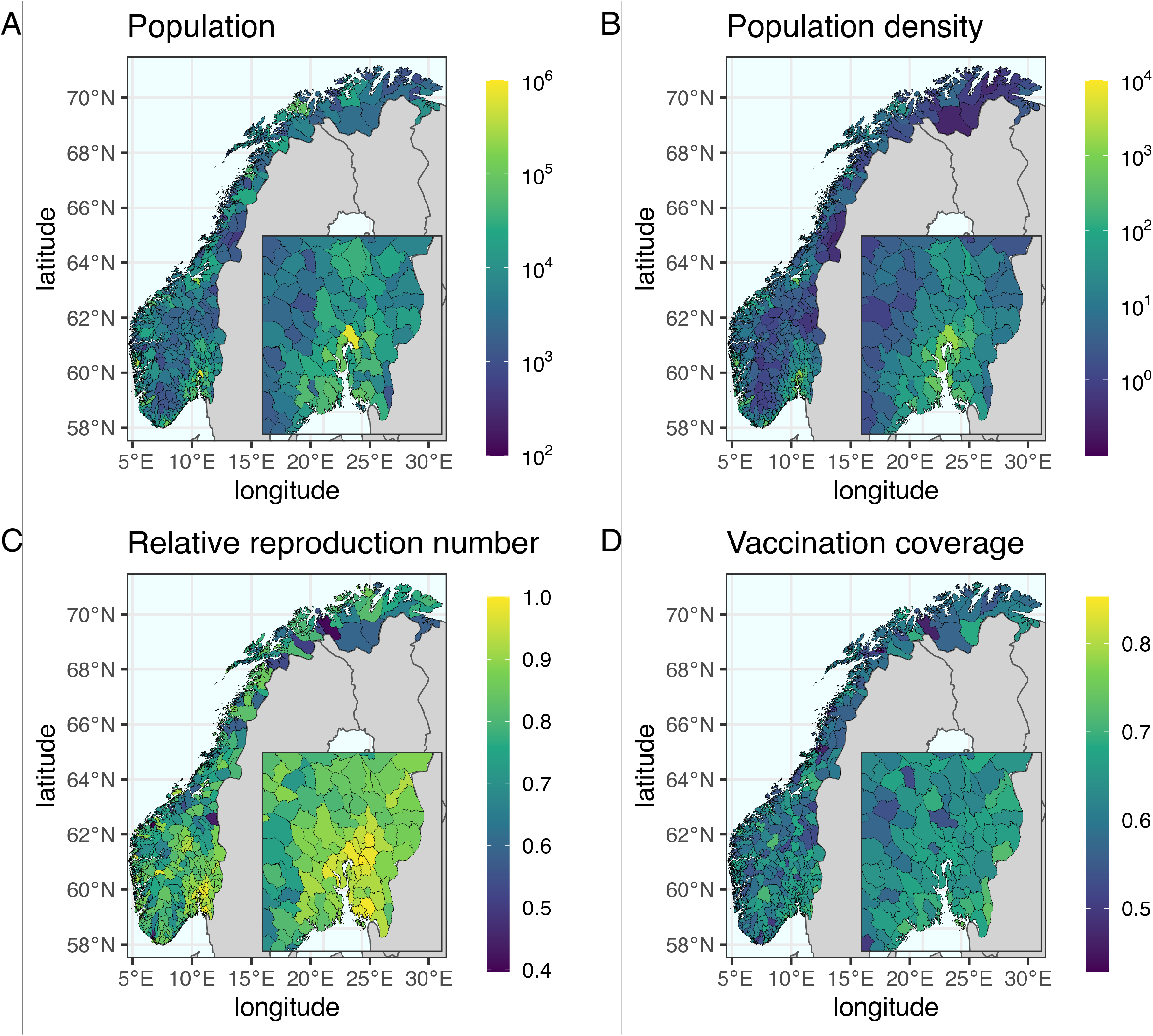
The population, density, relative reproduction number and vaccination coverage of 356 municipalities in Norway. A: The population size in log10-scale illustrates a highly heterogeneous geographic distribution. The population size in Oslo is about 700,000. B: The population density in log10-scale. The population density in Oslo is about 1500 per km2. C: The relative reproduction number is normalized with a maximum at 1. Oslo and its surrounding municipalities are the ones with the highest numbers. D: The vaccination coverage of the 1st dose. The overall coverage in Norway at the end of July 2021 was about 64%.

### S3 Data

There are mainly three sets of Norwegian data used in this study taken from Emergency preparedness register for COVID-19 (Beredt C19) [36].

#### S3.1 Registry data

There were in total 86289 confirmed cases, 2397 hospital admissions, 466 ICU admissions and 266 deaths registered and collected by Norwegian Surveillance System for Communicable Diseases (MSIS) [37] and the Norwegian Intensive Care and Pandemic Registry (NIPaR) [38, 39] from January to July 2021. Fig S10 shows the daily numbers with rolling averages, cumulative numbers, geographic distribution and age distributions of the 4 outcomes. Cases are defined as people who tested positive with a PCR test in Norway by date of testing. Hospitalizations include all admissions where COVID-19 was the main cause for the admission as determined by the doctor and where the date of admission and testing positive was after the 1st of January 2021. The number of ICU admissions are patients who tested positive and where admitted to an ICU after the 1st of January 2021. Deaths are COVID-19 associated deaths, defined as deaths reported by doctors as due to COVID-19 or as deaths with COVID-19 as the cause of death on the death certificate. We included COVID-19 associated deaths where the date of test and date of death was after the 1st of January 2021.

**Fig S10.**
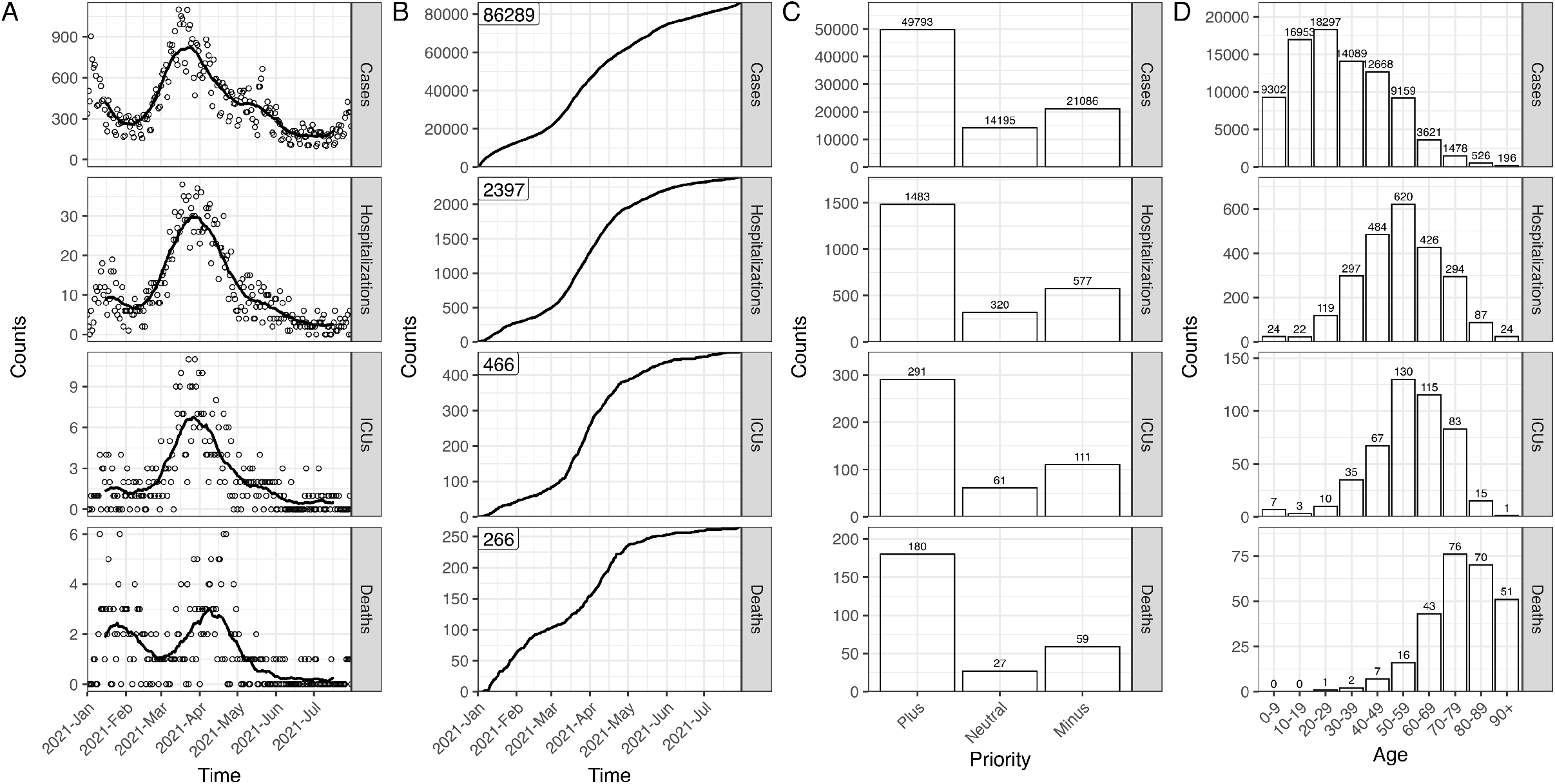
The observed data from January to July 2021 in Norway. A: The daily numbers (circles) and rolling averages (lines). B: The cumulative numbers. C: The priority distributions of geographic regions. D: The age distributions of confirmed cases, hospitalizations, ICU admissions and deaths are shown in each panel.

#### S3.2 Test data

The test positive data were reported by medical doctors and collected by Norwegian Surveillance System for Communicable Diseases (MSIS) [37]. The data from January to July 2021 shows that 45.1%, 38.6%, 9.9% and 6.2% of the cases were infected within households, in community, workplaces and schools, respectively. Community transmission refers to all routes (e.g. public event, private arrangement, prison, health institution, organized leisure activity, private event in public place, travel, collection in private homes, restaurant, bar, nightclub and public transport) other than households, schools or workplaces as the categorization in the individual-based model (IBM). Additionally, we note that the data of importation used in both models are shown Fig S2F.

#### S3.3 Vaccination data

SYSVAK is a national, electronic immunization registry that records an individual’s vaccination status and vaccination coverage in Norway [40]. The data were aggregated by date, municipality, age and risk status. Fig S11A shows the total number of first doses distributed from January to July in 2021 in Norway is about 3.5M. Fig S11B shows the age-specific coverage from January to July in 2021, and Fig S9D shows the geographic distribution of coverage. The models used the normalized data according to the synthetic population to match with the vaccination coverage.

**Fig S11.**
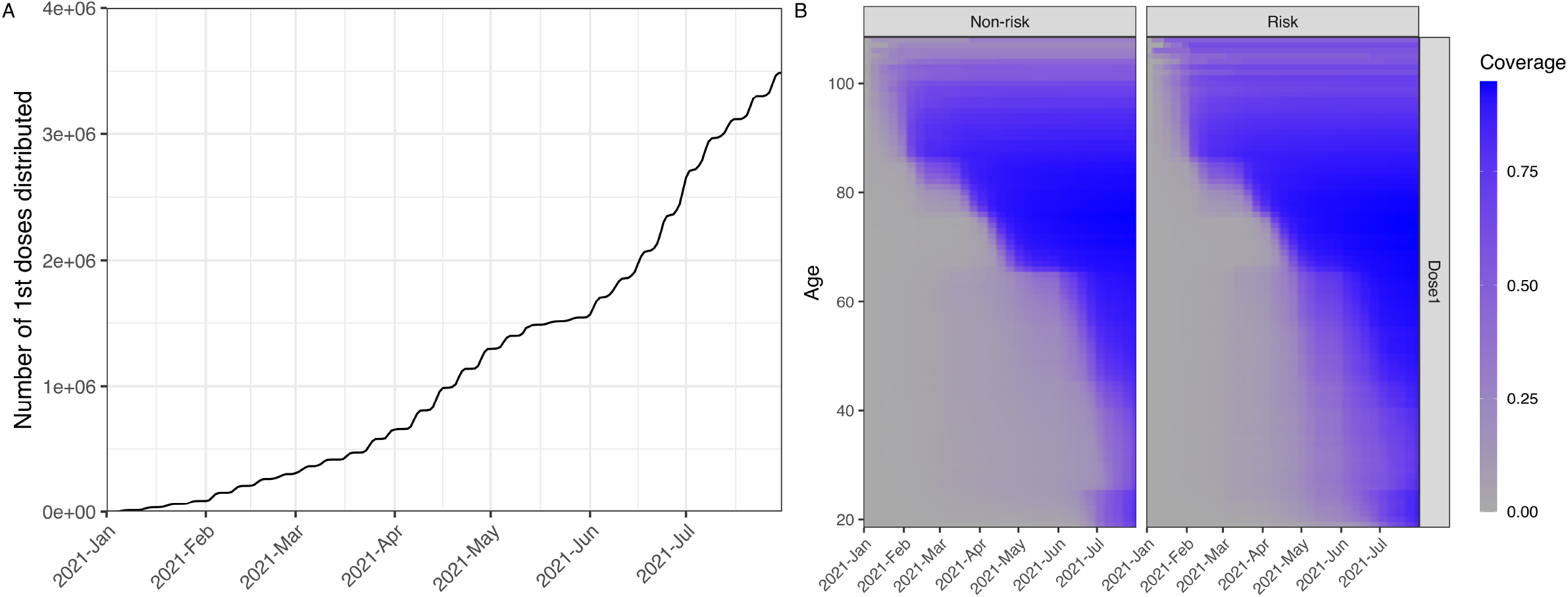
The vaccination data from SYSVAK. A: The cumulative number of first doses distributed nationally. The number of first doses per day taken from historical data is shared in all scenarios. B: The age- and risk-specific vaccination coverage. The coverage of each group from low to high is shown by color from gray to blue. The age prioritization is shown in Table S10.

### S4 Vaccination strategies

The distribution of all vaccine doses follows a prioritization process. Firstly, doses are allocated to different regions based on geographic prioritization, and secondly, within each region, doses are prioritized based on age (and risk) of individuals. Fig S12 illustrates the distribution of relative reproduction numbers for municipalities grouped by priority level (*Plus, Minus*, and *Neutral*) for each of the 15 selection strategies. The original selection strategy (∆*n* = 0) is based on the real implemented step-2 geographic prioritization plan in Norway, from the 19th of May 2021 [14]. The *Plus* group comprises municipalities with the highest regional reproduction numbers, while the *Neutral* group may not necessarily be located between the *Plus* and *Minus* groups. Table S9 presents the number of municipalities and their corresponding adult population fractions in each priority group.

**Fig S12.**
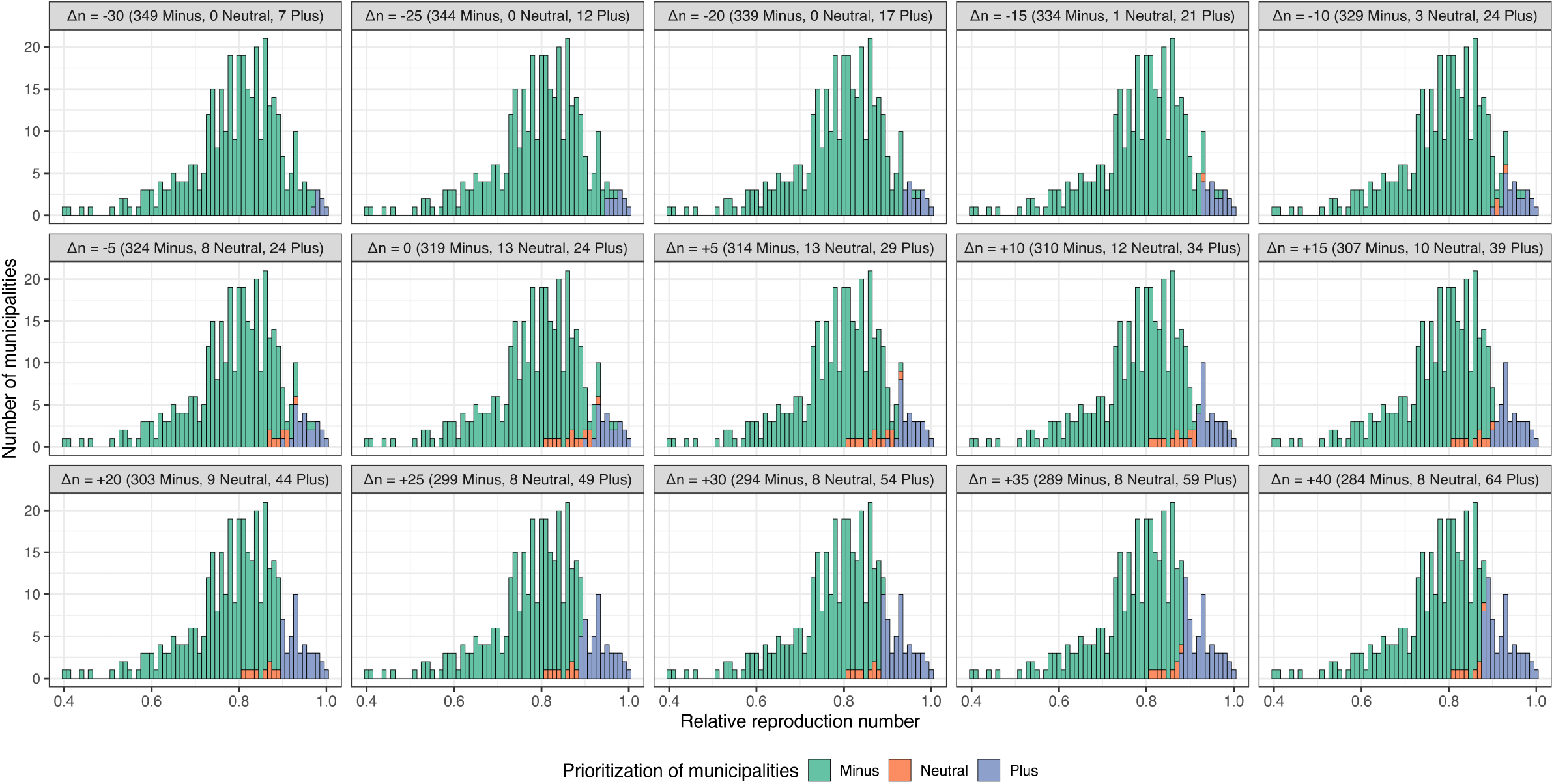
The relative reproduction number distribution of municipalities in alternative strategies. 356 municipalities with their own relative reproduction numbers are classified into 3 groups: *Minus, Neutral* and *Plus* colored in green, orange and blue. Each strategy has different numbers of municipalities in the three groups. The original prioritization (∆*n* = 0) is the selection in the real implementation. The index of moves of municipality priority ∆*n* is shown in brackets, and the number of municipalities in each group are shown accordingly on each panel title.

Fig S13 shows the total number of prioritized vaccine doses shifted from the *Minus* to the *Plus* group for all alternative strategies. The most extreme strategy (∆*p* = 300% and ∆*n* = *−*25) prioritizes 3 million doses to 12 municipalities in the *Plus* group. However, for each ∆*n*, there exists a maximum threshold of ∆*p*, as shown in Table S9. The smaller population size in the *Minus* group limits the total number of doses that can be prioritized to the *Plus* group. The percentage of extra prioritized doses can only reach a maximum of ∆*p* = 70% by moving 40 municipalities from the *Minus* group (∆*n* = +40) in one extreme direction. Conversely, prioritized doses can reach up to ∆*p* = 300% by moving 30 municipalities from the *Plus* group (∆*n* = *−*30) in another extreme direction.

Table S10 shows the age prioritization categories for different risk groups and their corresponding population sizes. Within each region, vaccines are distributed according to these priority groups. However, in reality, a portion of vaccine doses are allocated to healthcare workers, who face higher risk due to their front-line roles, while simultaneously prioritizing the elderly. The NIPH modeling report [70] determined the age prioritization for the general population.

Notably, those aged 25-39 years old are the lowest priority group, after individuals aged 18-24 years old.

Fig S14 illustrates the age-based trade-offs resulting from different geographic prioritization strategies. The percentage of extra doses (∆*p*) is varied while municipality priority moves are fixed (∆*n* = − 25). The geographic prioritization of vaccine distribution can ultimately influence age-based prioritization. For instance, without geographic prioritization (∆*p* = 0), all individuals aged 80 years and above can receive their first dose by March. Conversely, under the most extreme strategy (∆*p* = 300%), they can receive it by April. There are switches between strategies for other age groups, such as in mid-February and early April for those aged 70-79 and 60-69 years old, respectively. It is important to note that the results from the IBM refer to the timing of vaccination, while those from the MPM indicate the timing of vaccine effectiveness, resulting in a time delay between the two models.

**Table S9.** The characteristics of the shifts of municipality priority ∆*n*.

| $\Delta n$ | Number of municipalities | | | Adult population fraction (%) | | | Population ratio (%) |
| --- | --- | --- | --- | --- | --- | --- | --- |
|  | *Minus* | *Neutral* | *Plus* | *Minus* | *Neutral* | *Plus* | *Minus / Plus* |
| -30 | 349 | 0 | 7 | 80.5 | 0.0 | 19.5 | 412 |
| -25 | 344 | 0 | 12 | 76.3 | 0.0 | 23.7 | 321 |
| -20 | 339 | 0 | 17 | 72.7 | 0.0 | 27.3 | 265 |
| -15 | 334 | 1 | 21 | 69.5 | 0.4 | 30.1 | 230 |
| -10 | 329 | 3 | 24 | 61.6 | 6.8 | 31.6 | 194 |
| -5 | 324 | 8 | 24 | 56.9 | 11.5 | 31.6 | 180 |
| **0** | **319** | **13** | **24** | **48.0** | **20.4** | **31.6** | **151** |
| +5 | 314 | 13 | 29 | 47.7 | 20.4 | 31.9 | 149 |
| +10 | 310 | 12 | 34 | 46.6 | 19.9 | 33.4 | 139 |
| +15 | 307 | 10 | 39 | 44.8 | 13.5 | 41.7 | 107 |
| +20 | 303 | 9 | 44 | 44.3 | 13.0 | 42.7 | 103 |
| +25 | 299 | 8 | 49 | 43.6 | 12.5 | 43.9 | 99 |
| +30 | 294 | 8 | 54 | 42.9 | 12.5 | 44.6 | 96 |
| +35 | 289 | 8 | 59 | 42.5 | 12.5 | 45.0 | 94 |
| +40 | 284 | 8 | 64 | 41.0 | 12.5 | 46.5 | 88 |

For each $\Delta n$ , the table shows the number of municipalities, their corresponding adult population fraction in *Minus*, *Neutral* and *Plus* groups and the population ratio of *Minus* and *Plus*.

**Table S10.** The age prioritization in different risk categories.

| Priority order of risk categories | Population |
| --- | --- |
| All aged 85+ | 116995 |
| All aged 75-84 | 290097 |
| All aged 65-74 | 534724 |
| All healthcare workers | 322994 |
| People with underlying diseases, aged 55-64 | 164691 |
| People with underlying diseases, aged 45-54 | 119524 |
| People with underlying diseases, aged 18-44 | 154318 |
| Other people, aged 55-64 | 422891 |
| Other people, aged 45-54 | 563932 |
| Other people, aged 18-24 and 40-44 | 662403 |
| Other people, aged 25-39 | 896401 |

The priority of each risk category is placed in order. The oldest age group is given the highest priority among all while those aged 25-39 years without underlying diseases are the least. Their approximate population are shown accordingly.

**Fig S13.**
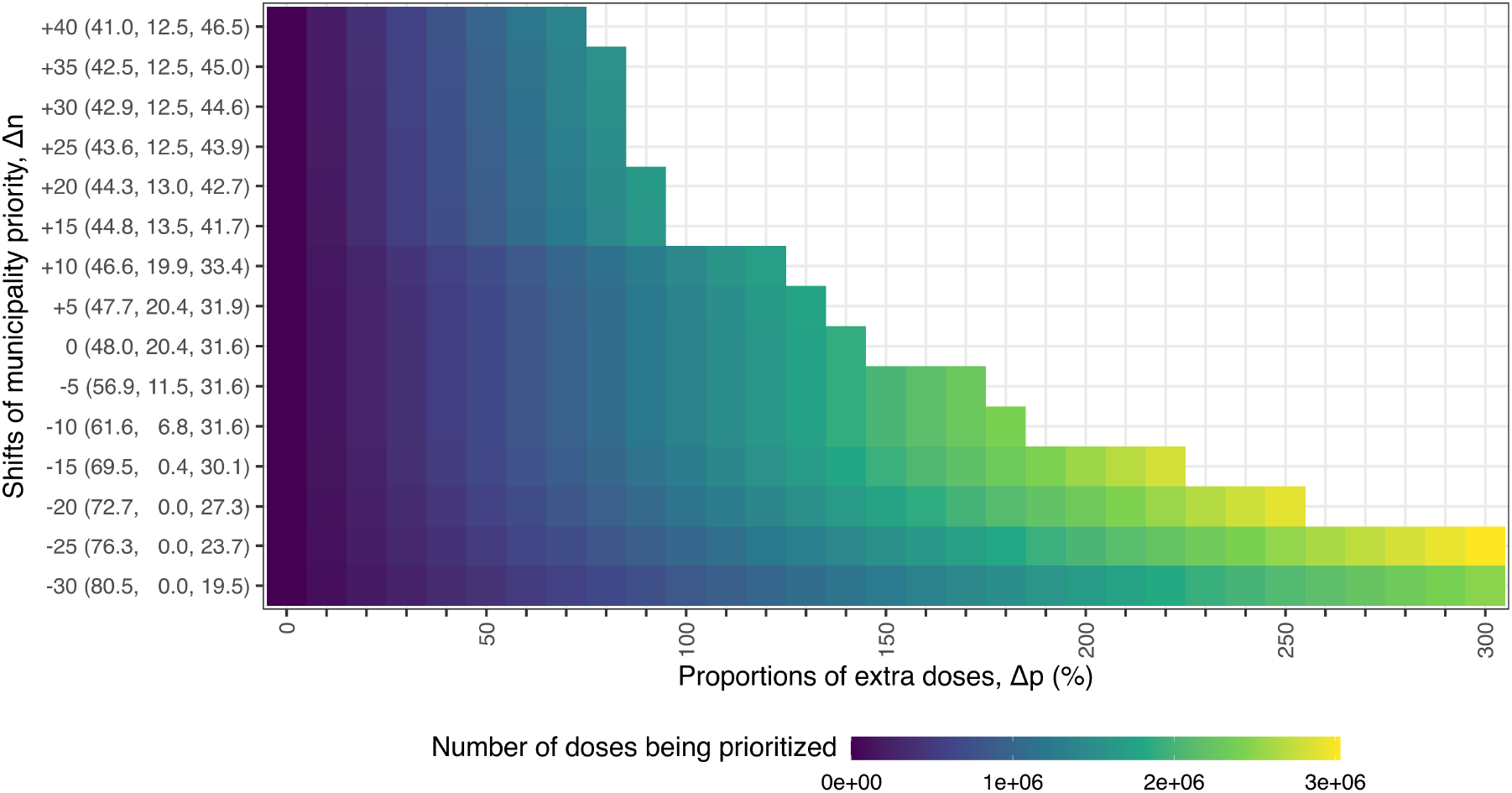
The number of prioritized doses distributed from the *Minus* group to the *Plus* group in the alternative strategies. The color shows the number of doses from low to high by color blue to yellow. The baseline strategy without geographic prioritization on the first column (∆*n* = 0%) refers to no doses being prioritized. The y-axis shows the population fractions (%) of three groups (*Minus, Neutral*, and *Plus*) for each shift in municipality priority (∆*n*). The geographic distribution of municipality priority (∆*n*) can be found in Fig 2. The maximum thresholds of ∆*p* can be found in Table S9.

### S5 The scenario without vaccination

We considered a scenario without vaccination to show the effect of the mass vaccination program. In the absence of vaccination, as shown in Table S11, all infections, hospitalizations, ICU admissions, and deaths were higher than in scenarios where vaccination was implemented. All RRR were negative. In the IBM, the mean RRR (and their 95% CIs) were -5.8 (−6.3 to -5.4)%, -17.2 (−17.7 to -16.7)%, -20.0 (−20.7 to -19.4)%, and -80.7 (−82.1 to -79.3)%, corresponding to 13,734 (12,723 to 14,746) more infections, 455 (443 to 469) more hospitalizations, 103 (100 to 106) more ICU admissions, and 190 (187 to 194) more deaths than the baseline strategy, respectively. In the MPM, the mean RRR (and their 95% CIs) were -13.5 (−13.9 to -13.1)%, -26.4 (−26.9 to -25.9)%, -28.7 (−29.4 to -27.9)%, and -106.0 (−107.0 to -104.0)%, corresponding to 26,246 (25,476 to 27,017) more infections, 574 (563 to 586) more hospitalizations, 118 (115 to 121) more ICU admissions, and 252 (249 to 255) more deaths than the baseline strategy, respectively. The effective reproduction number from the IBM is shown in Fig S19.

**Fig S14.**
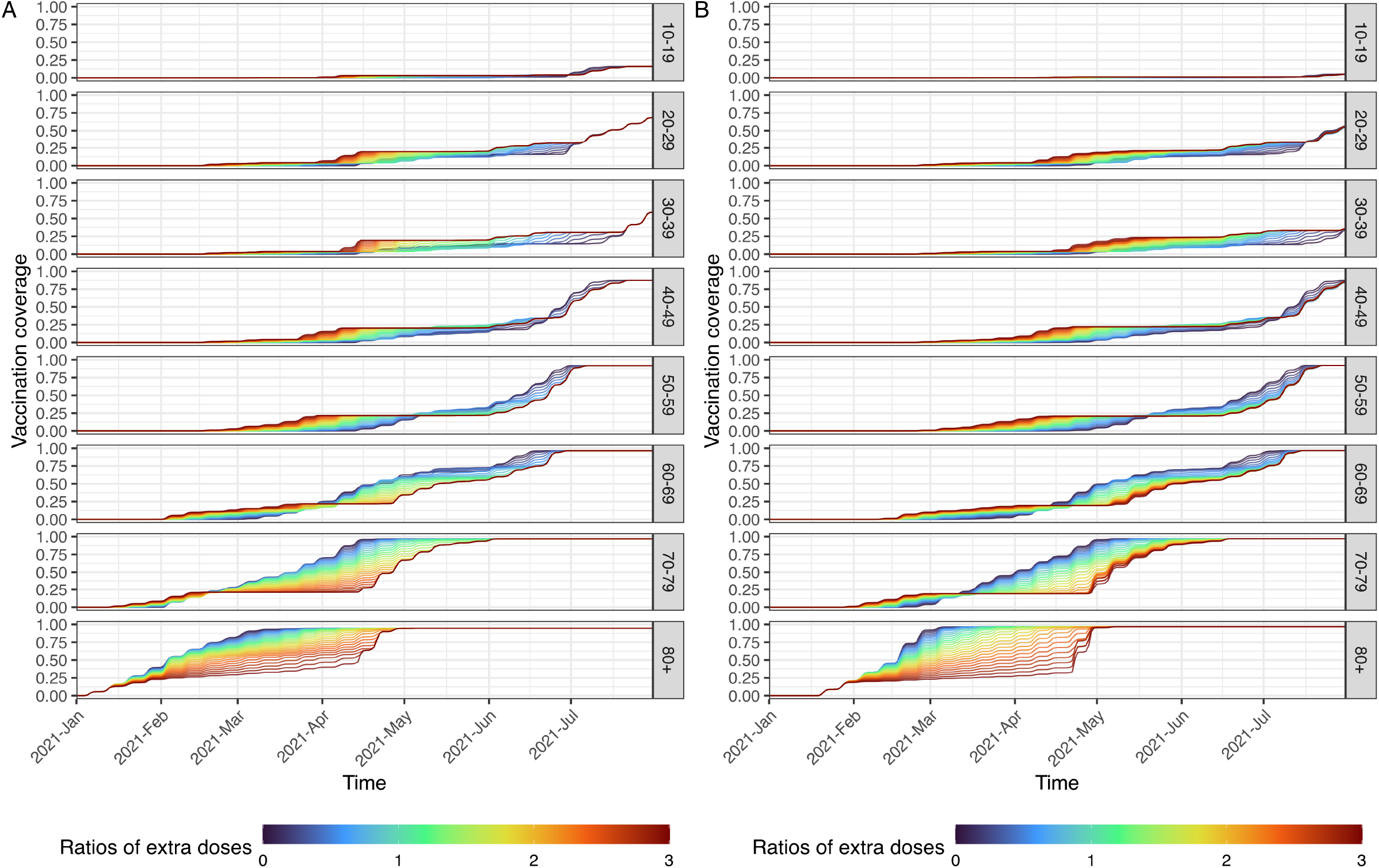
The age-specific vaccination coverage in the alternative strategies. Each line in different colors shows a strategy of alternative ratio of extra doses (∆*p*) given fixed moves of municipality priority (∆*n* = *−*25%). The blue and red color present lower and higher levels of geographic prioritization, respectively. Given a higher level of geographic prioritization, the priority of the older people is lower, for example fewer doses are distributed to those above 80 years old during the first few months. A: The timing of vaccination in the IBM. B: The timing of vaccine protection activated in the MPM. Note that there is a 2-week time delay from vaccination to providing protection in the MPM.

### S6 Additional results in the alternative strategies

Fig S15 and Fig S16 show the geographic- and age-specific RRR from both IBM and MPM in the alternative strategies, respectively. Fig S17 and Fig S18 show the RRR from both IBM and MPM in the alternative starting time, respectively. Fig S19 shows the effective reproduction numbers from the IBM. We note that the reproduction numbers are the case reproduction numbers instead of instantaneous reproduction numbers [109–111]. Four scenarios (i) the actual strategy, i.e. the reality; (ii) the scenario without vaccination; (iii) the baseline strategy without geographic prioritization; and (iv) the optimal strategy (∆*n* = −25, ∆*p* = 300%) for minimizing infections as shown in the main text are included.

**Table S11.** The relative risk reduction (RRR) of different strategies compared to the baseline strategy without geographic prioritization.

| Models | Outcomes | No vaccination | Best strategy |
| --- | --- | --- | --- |
| | | RRR, % (95% CI) | RRR, % (95% CI) [$\Delta p$, $\Delta n$] |
| IBM | Infections | -5.8 (-6.3, -5.4) | 16.4 (16.1, 16.7) [300%, -25] |
| IBM | Hospitalizations | -17.2 (-17.7, -16.7) | 19.4 (19.0, 19.7) [250%, -20] |
| IBM | ICUs | -20.0 (-20.7, -19.4) | 19.2 (18.8, 19.6) [250%, -20] |
| IBM | Deaths | -80.7 (-82.1, -79.3) | 8.0 (7.5, 8.6) [100%, +10] |
| MPM | Infections | -13.5 (-13.9, -13.1) | 16.0 (15.7, 16.3) [300%, -25] |
| MPM | Hospitalizations | -26.4 (-26.9, -25.9) | 15.4 (15.0, 15.7) [300%, -25] |
| MPM | ICUs | -28.7 (-29.4, -27.9) | 15.6 (15.1, 16.1) [300%, -25] |
| MPM | Deaths | -106.0 (-107.0, -104.0) | 5.5 (4.9, 6.2) [70%, -15] |

The mean RRR (and their 95% confidence intervals) of the scenario without vaccination and the best strategies are shown in the two columns. The best strategies, selected separately for each health outcome, are shown in square brackets.

**Fig S15.**
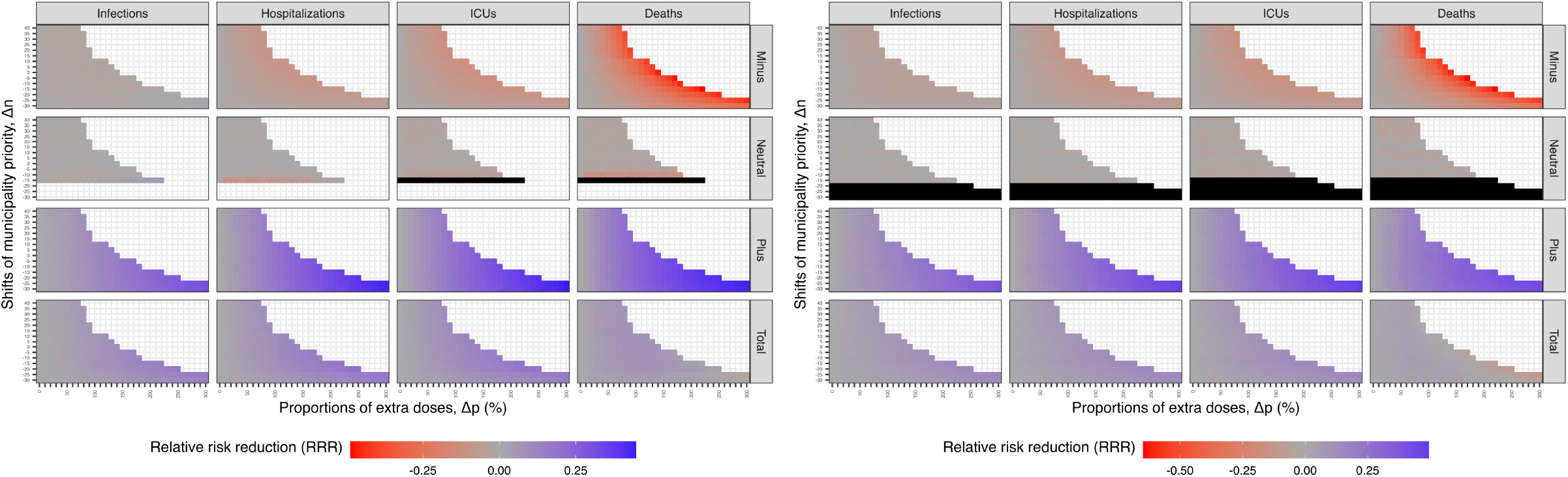
The geographic-specific mean RRR from both (Left) IBM and (Right) MPM in the alternative strategies. The RRR is calculated for each 3 geographic-prioritization groups and 4 health outcomes on infections, hospitalizations, ICU admissions, and deaths in alternative selection of municipality priority and level of geographic prioritization. The black color represents unidentifiable RRR values due to low influence or the absence of municipalities in *Neutral* group. The total RRR on the bottom row shows the total of all geographic groups.

Fig S20 and Fig S21 show the comparison of two models in the actual strategy and optimal strategy for minimizing infections, respectively. Fig S21 shows the comparison in the geographic difference on top of the trends and age distributions in Fig 6 in the main text. In general, there are more counts, especially infections, hospitalizations and ICU admissions, in the *Plus* group in the IBM than MPM.

**Fig S16.**
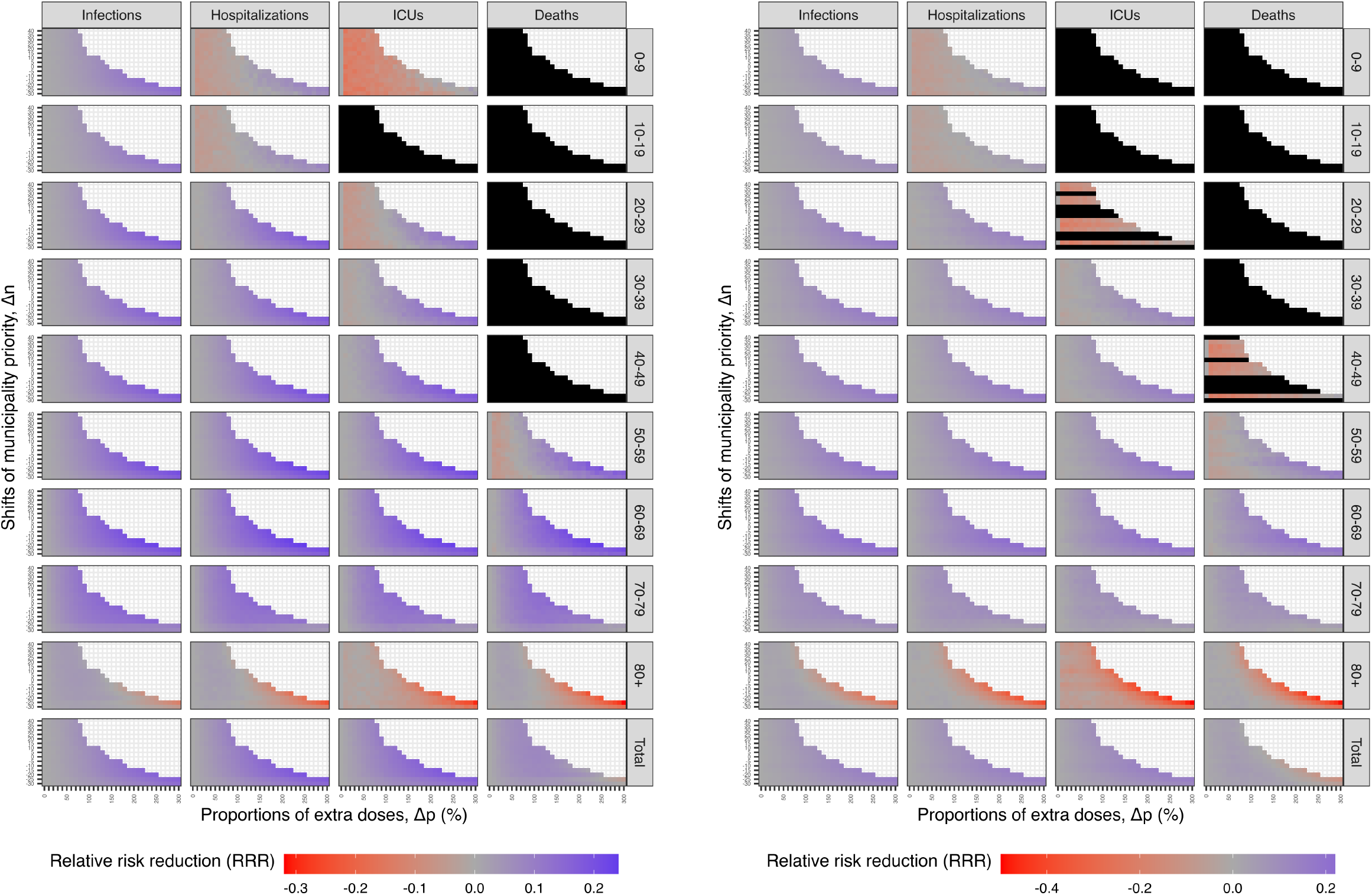
The age-specific mean RRR from both (Left) IBM and (Right) MPM in the alternative strategies. The RRR is calculated for each 9 age groups and 4 health outcomes on infections, hospitalizations, ICU admissions, and deaths in alternative selection of municipality priority and level of geographic prioritization. The black color represents unidentifiable RRR values due to low influence of severe outcomes among the younger population. The total RRR on the bottom row shows the total of all ages.

### S7 Sensitivity analysis

In this sensitivity analysis, the entire analysis was performed without the factor of 0.5 on the probabilities of hospitalization. We found that our results are not sensitive to the factor of 0.5 on the probabilities of hospitalization by obtaining similar results in both IBM and MPM.

Both models are re-calibrated that 12 free parameters were estimated by fitting to the daily and age-specific hospital admissions using Latin hypercube sampling (LHS) and least squares method. Tables S12 and S13 show the age-specific probabilities of death given infection from two models, respectively. The probabilities calculated based on the ratios of observed number of deaths to the estimated mean number of infections. Fig S22A and Fig S23A show the distributions of calibrated parameters and the fits to observed data from the IBM. Fig S22B and Fig S23B show the ones from the MPM.

**Fig S17.**
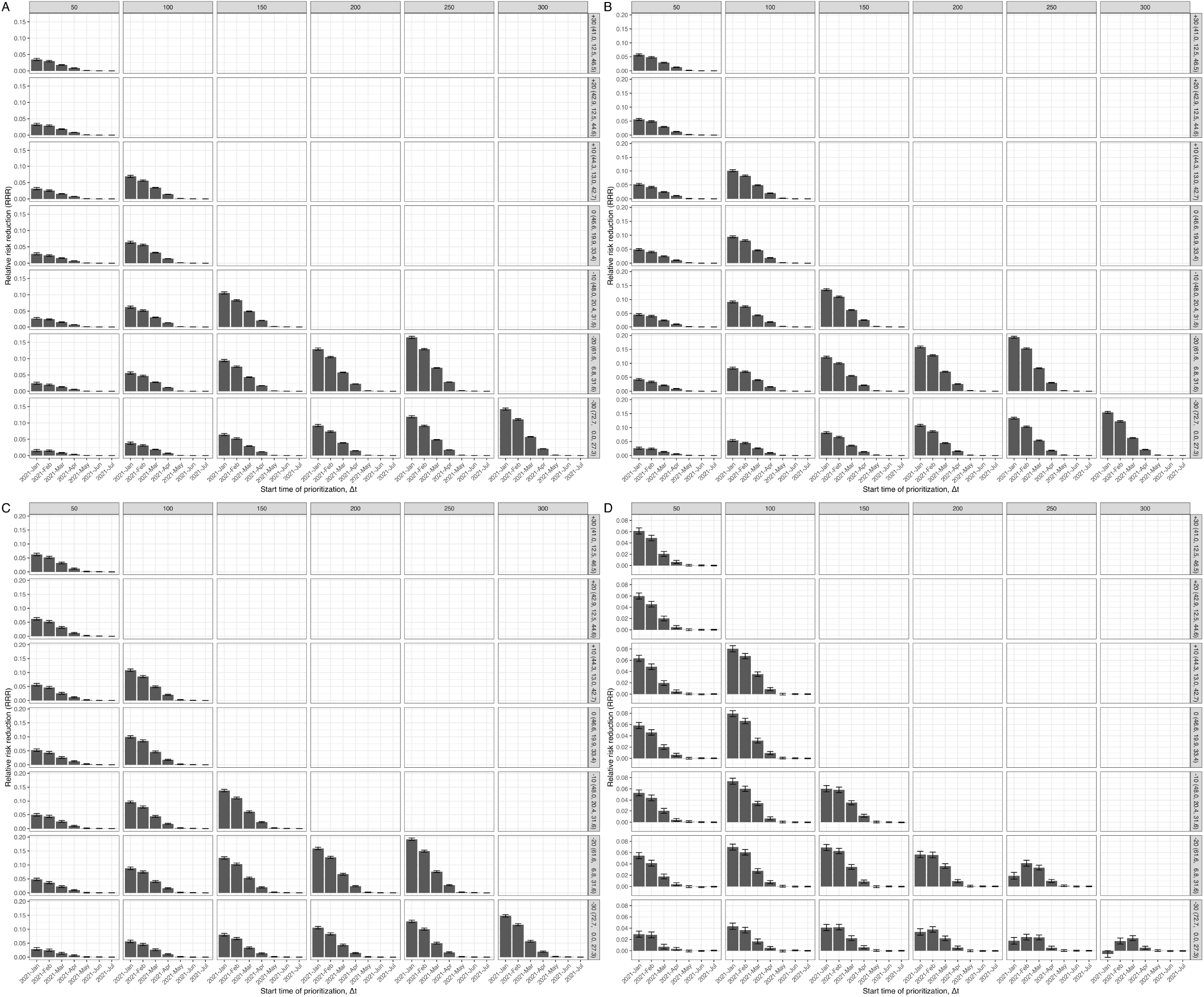
The RRR in the alternative starting time for the IBM. Each panel shows the RRR for (A) infections, (B) hospitalizations, (C) ICU admissions, and (D) deaths. While the RRR for infections, hospitalizations, and ICU admissions decrease by starting time across all strategies, the RRR of deaths decrease by starting time only when ∆*p <* 200%. The bars show the mean values and their 95% confidence intervals using 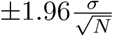, where *σ* is standard deviation of RRR and *N* = 1000 is the number of simulations. The benefits are limited if the geographic prioritization started after May 2021.

**Fig S18.**
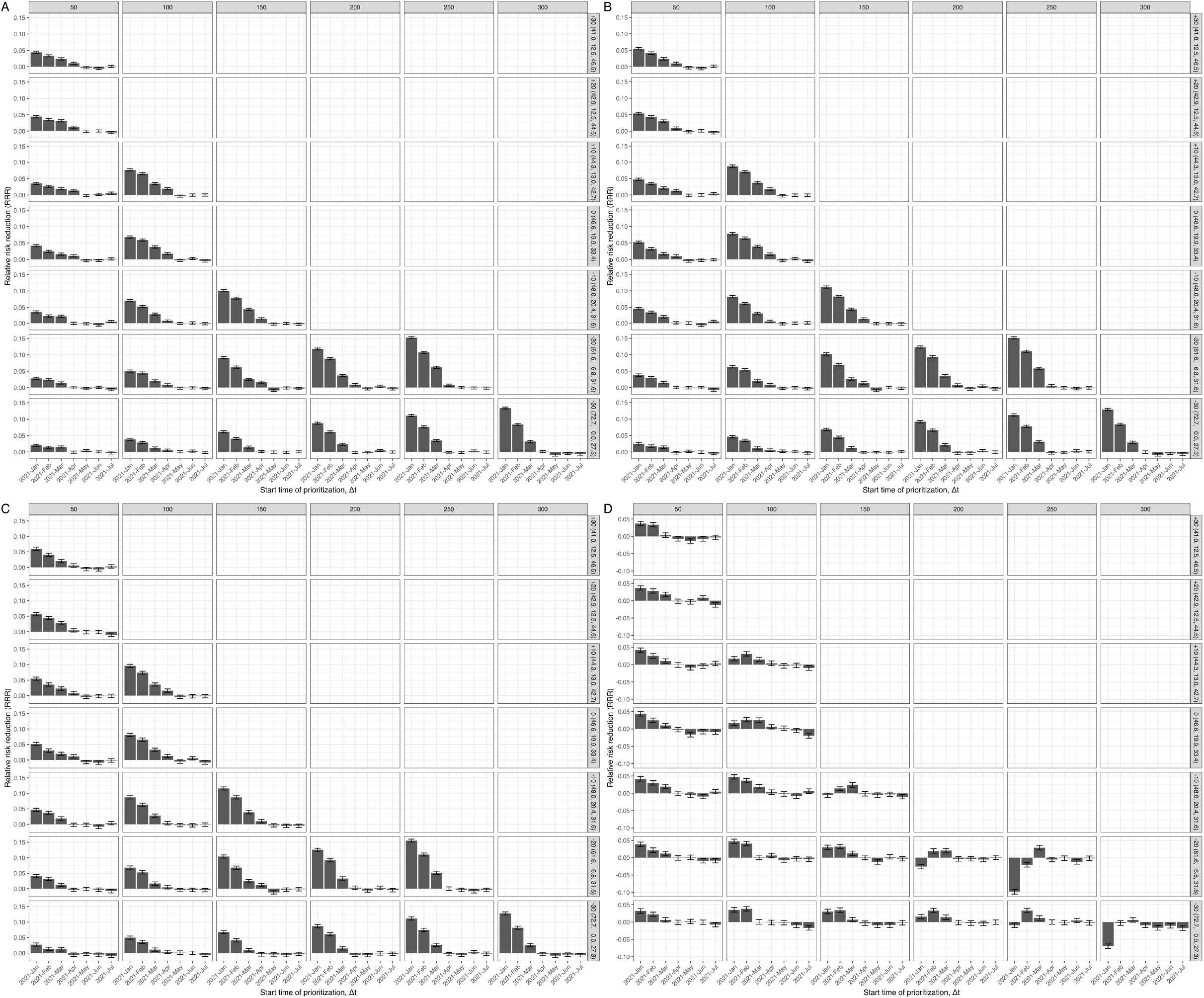
The RRR in the alternative starting time for the MPM. Each panel shows the RRR for (A) infections, (B) hospitalizations, (C) ICU admissions, and (D) deaths. While the RRR for infections, hospitalizations, and ICU admissions decrease by starting time across all strategies, the RRR of deaths decrease by starting time only when ∆*p <* 200%. The bars show the mean values and their 95% confidence intervals using 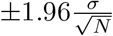, where *σ* is standard deviation of RRR and *N* = 1000 is the number of simulations. The benefits are limited if the geographic prioritization started after May 2021.

**Fig S19.**
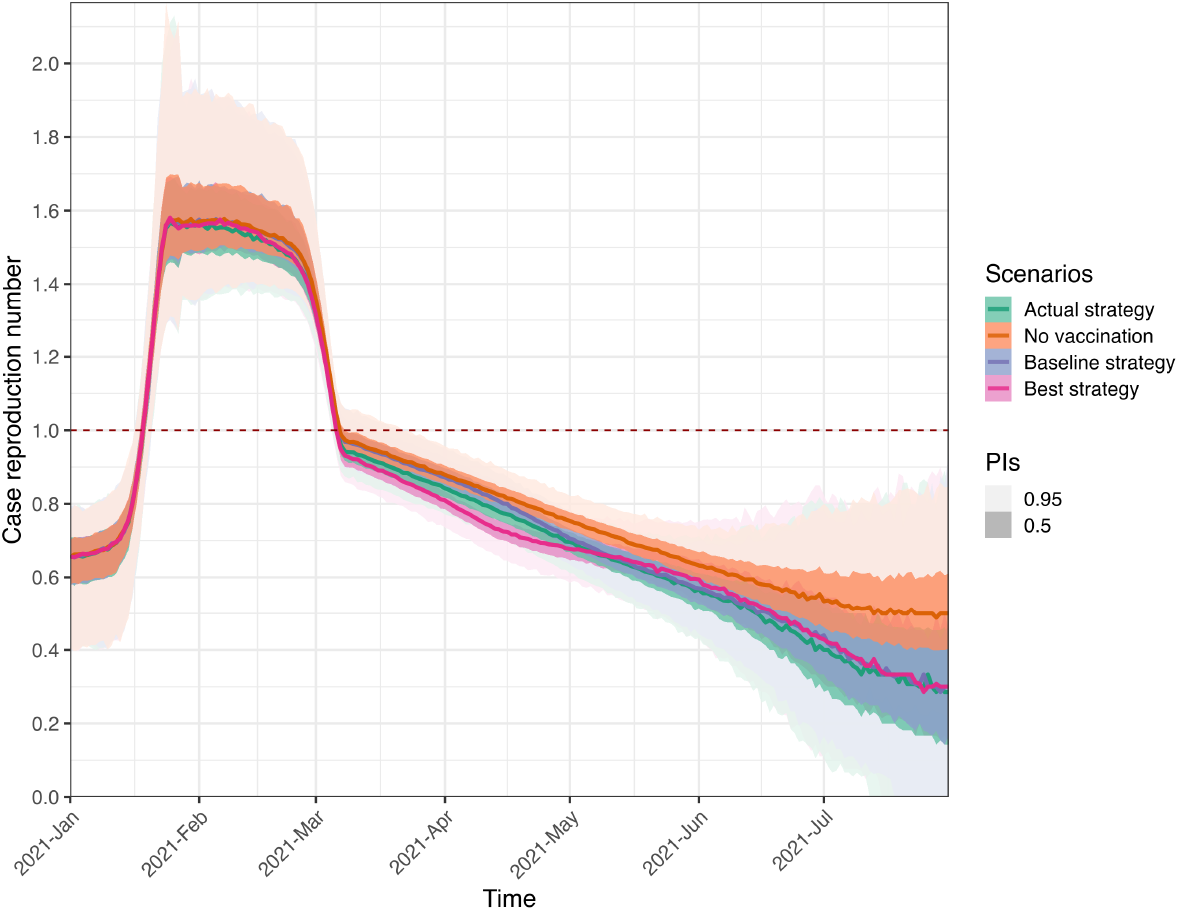
The effective reproduction numbers (Rt) in the IBM. The case reproduction numbers from the IBM are shown given (i) the actual strategy, i.e. the reality; (ii) the scenario without vaccination; (iii) the baseline strategy without geographic prioritization; and (iv) the optimal strategy (∆*n* = *−*25, ∆*p* = 300%) for minimizing infections as shown in the main text. The case reproduction numbers are calculated by using simulated transmission chains of all infectious individuals [109–111]. The lines show the mean and the colored areas represent the 50 and 95% prediction intervals.

**Table S12.** The age-specific probabilities of death given infection and ICU admission given hospitalization in the IBM, in the sensitivity analysis without the factor of 0.5 on the probabilities of hospitalization.

| Age groups | Probabilities of death (%) |  | Probabilities of ICU admission (%) |
| --- | --- | --- | --- |
|  | Without risk factors | With risk factors | All population |
| 0-9 | 0.00009 | 0.0002 | 31.6 |
| 10-19 | 0.0004 | 0.0007 | 6.6 |
| 20-29 | 0.005 | 0.009 | 9.8 |
| 30-39 | 0.02 | 0.03 | 10.4 |
| 40-49 | 0.08 | 0.1 | 13.2 |
| 50-59 | 0.2 | 0.3 | 20.8 |
| 60-69 | 1.3 | 2.4 | 26.5 |
| 70-79 | 5.8 | 10.6 | 28.2 |
| 80+ | 19.4 | 35.8 | 13.3 |

We assume that the probabilities of death depend on both age and risk status while the probabilities of ICU admission depend on age only. The age-specific probabilities of death given infection are calculated based on the observed number of deaths and the estimated mean number of infections. The age-specific probabilities of ICU admission given hospitalization are calculated based on the observed numbers of ICU admissions and hospitalizations.

**Fig S20.**
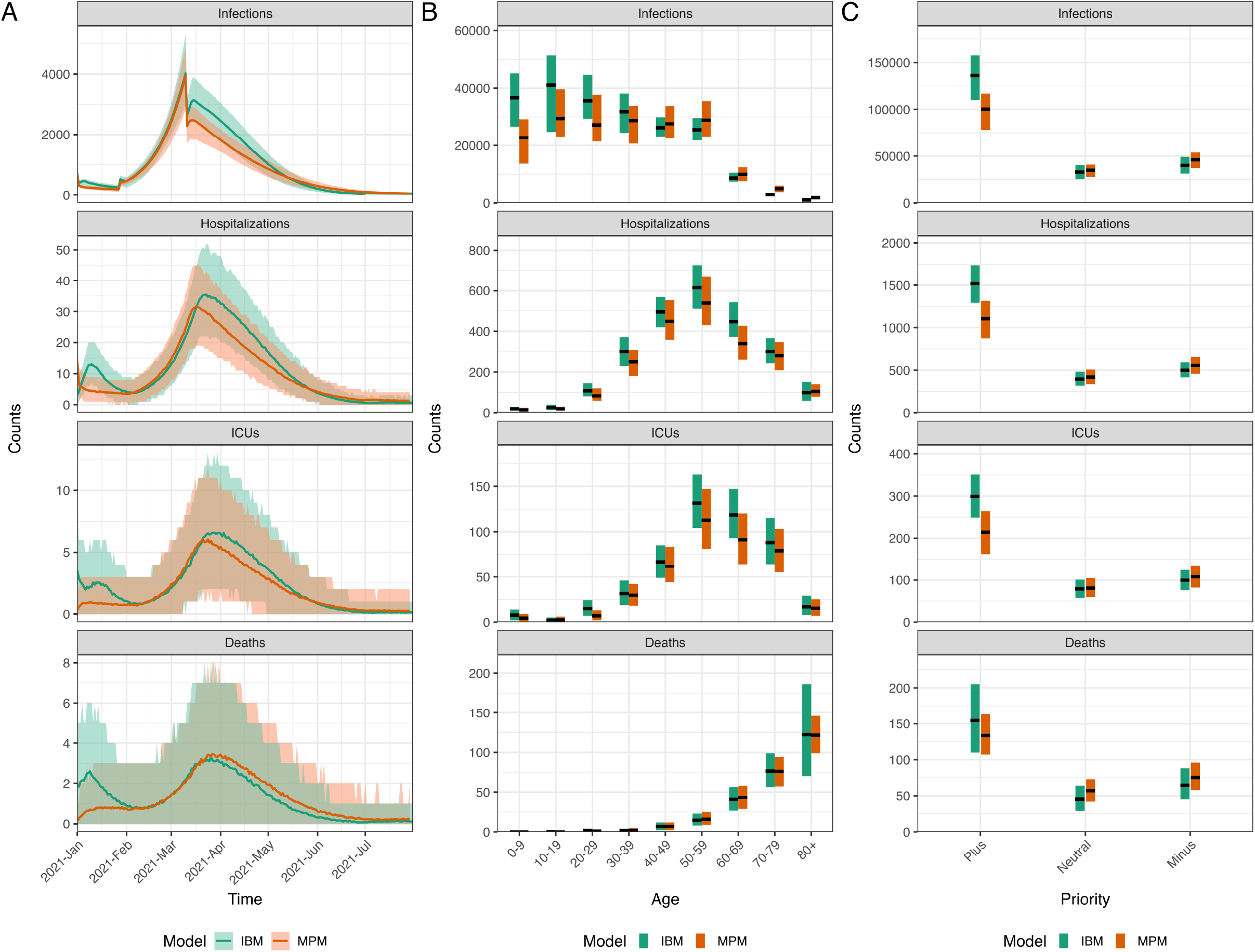
The comparison of two models in the actual strategy. A: The time series data of all ages. The lines show the model fits with their 95% prediction intervals represented by colored areas. B: The age distribution of total counts. C: The geographic-specific priority distribution of total counts. The black lines show the median values, and the colored areas show the 95% prediction intervals.

**Fig S21.**
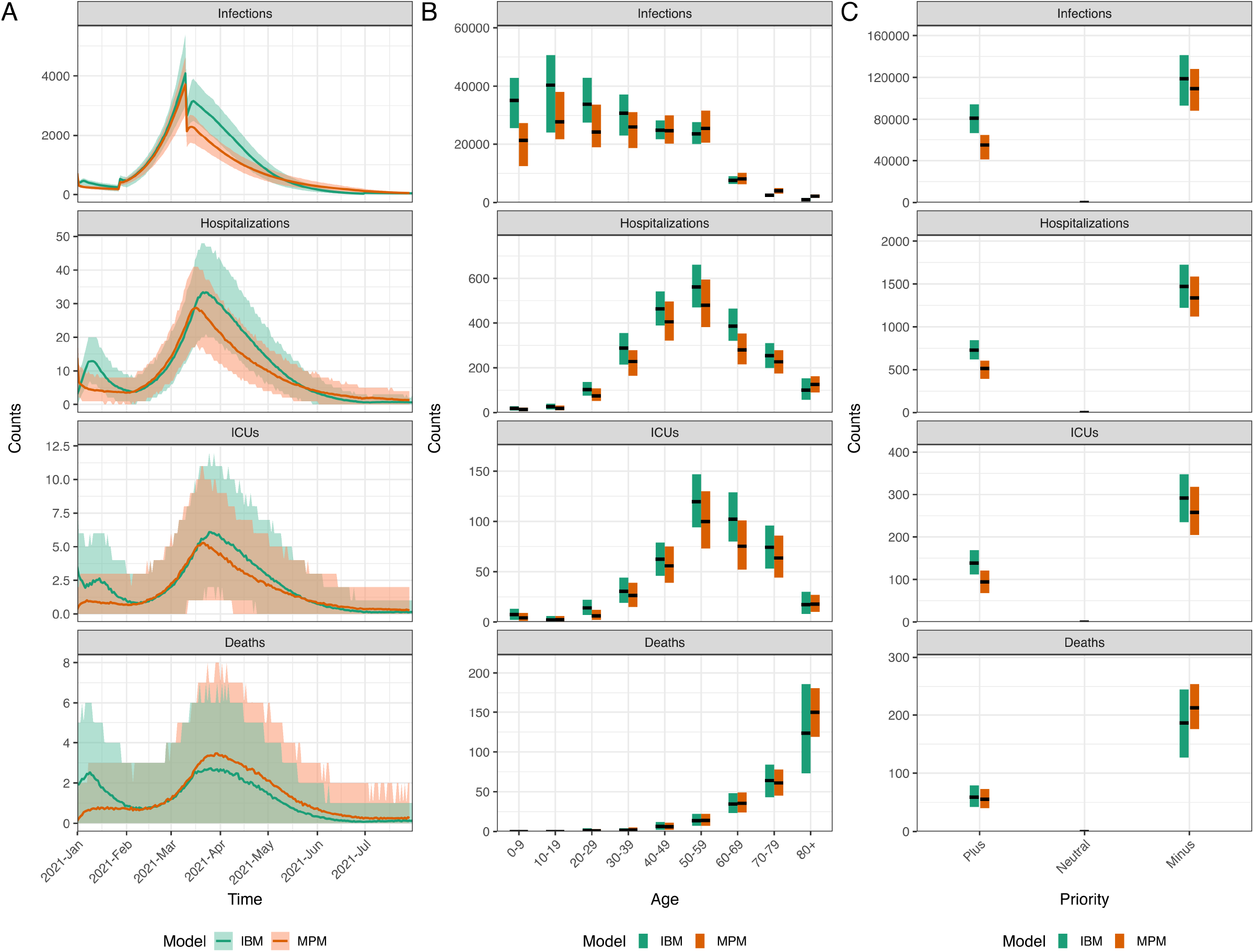
The comparison of two models in the optimal strategy for minimizing infections. A: The time series data of all ages. The lines show the model fits with their 95% prediction intervals represented by colored areas. B: The age distribution of total counts. C: The geographic-specific priority distribution of total counts. The black lines show the median values, and the colored areas show the 95% prediction intervals.

**Table S13.** The age-specific probabilities of death given infection and ICU admission given hospitalization in the MPM, in the sensitivity analysis without the factor of 0.5 on the probabilities of hospitalization.

| Age groups | Probabilities of death (%) |  | Probabilities of ICU admission (%) |
| --- | --- | --- | --- |
|  | Without risk factors | With risk factors | All population |
| 0-9 | 0.00007 | 0.0001 | 29.2 |
| 10-19 | 0.0001 | 0.0002 | 13.6 |
| 20-29 | 0.001 | 0.003 | 8.4 |
| 30-39 | 0.002 | 0.004 | 11.8 |
| 40-49 | 0.01 | 0.02 | 13.8 |
| 50-59 | 0.05 | 0.1 | 21.0 |
| 60-69 | 0.4 | 0.7 | 27.0 |
| 70-79 | 2.8 | 5.2 | 28.2 |
| 80+ | 15.4 | 28.5 | 14.4 |

We assume that the probabilities of death depend on both age and risk status while the probabilities of ICU admission depend on age only. The age-specific probabilities of death given infection are calculated based on the observed number of deaths and the estimated mean number of infections. The age-specific probabilities of ICU admission given hospitalization are calculated based on the observed numbers of ICU admissions and hospitalizations.

**Fig S22.**
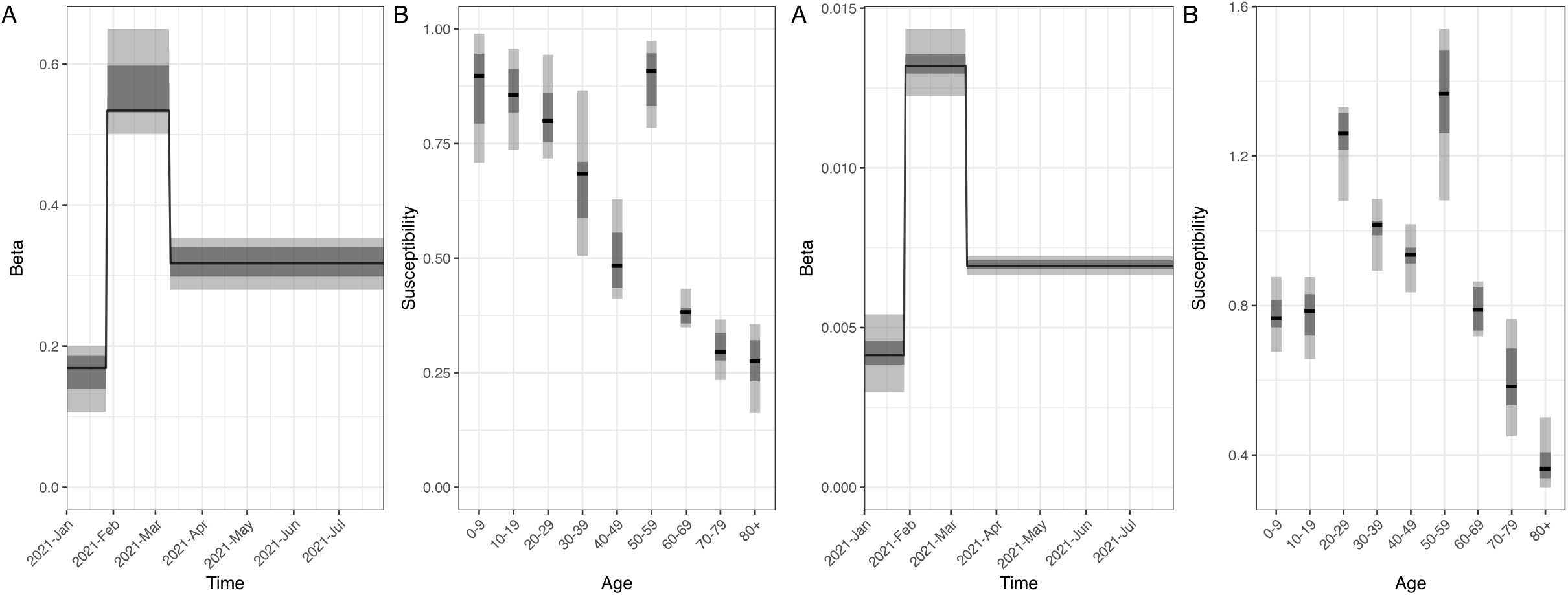
The marginal distributions of 12 estimated parameters in both (Left) IBM and (Right) MPM given doubled hospitalization probabilities. (A) Beta in 3 time periods and (B) susceptibilities of 9 age groups are sampled from Latin hypercube sampling (LHS) approach. The black lines show the median values, and the darker (lighter) gray areas show the ranges of 25th and 75th percentiles, i.e. lower and upper quantiles (2.5th and 97.5th percentiles) of the estimated parameters.

**Fig S23.**
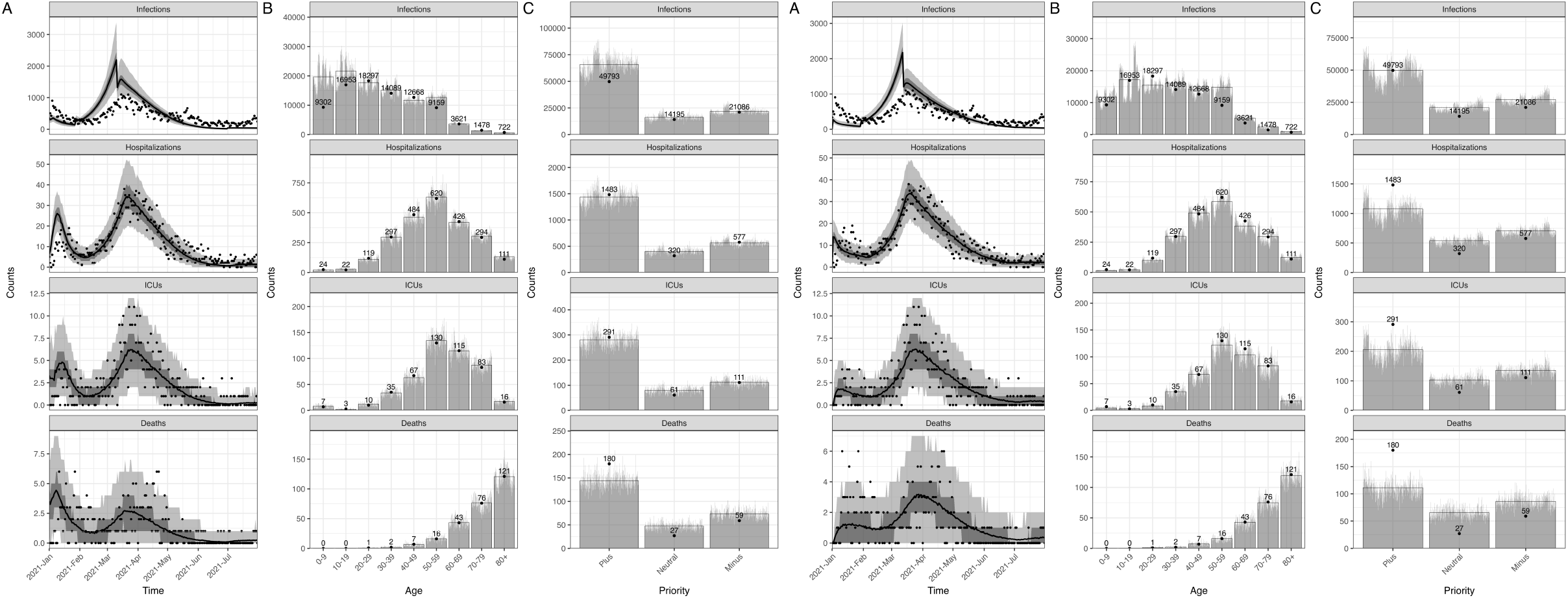
The calibrated (Left) IBM and (Right) MPM with observed data given doubled hospitalization probabilities. A: The time series data of all ages. The lines show the model fits with their 50 and 95% prediction intervals represented by gray areas. The dots show the observed data. B: The age distribution of total counts. C: The geographic-specific priority distribution of total counts. The gray bars show the counts of each simulation and the full bars with black borders show the mean of all simulations. The data are shown in dots with their exact numbers.

**Fig S24.**
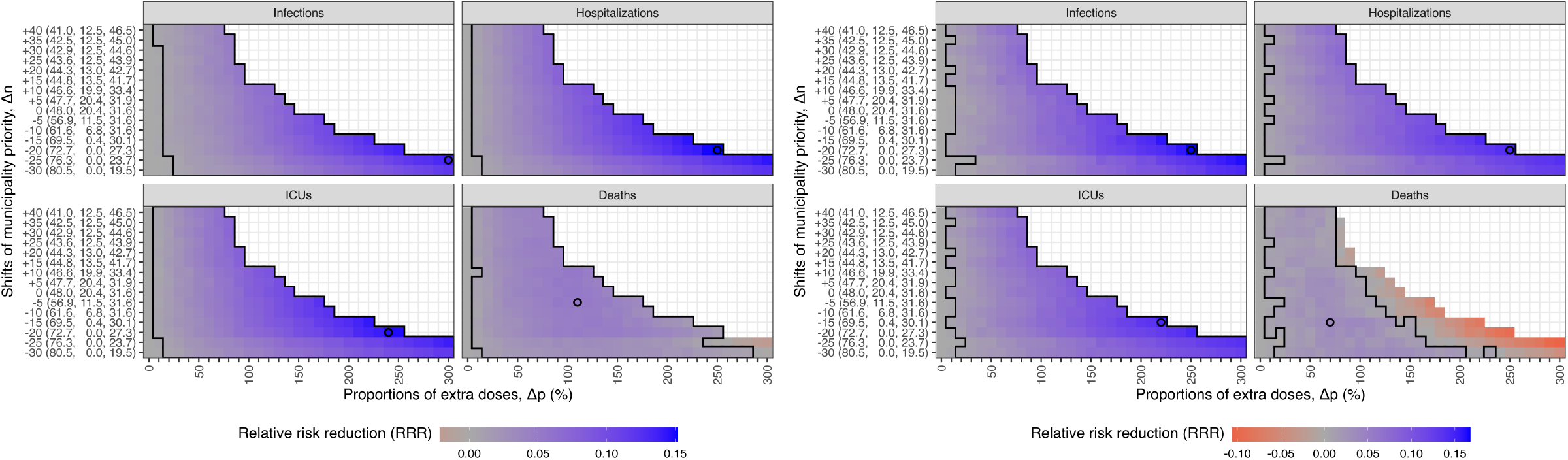
The mean relative risk reduction (RRR) of health outcomes from both (Left) IBM and (Right) MPM in the alternative strategies given doubled hospitalization probabilities. The mean RRR of infections, hospitalizations, ICU admissions and deaths from negative to positive is represented by color from red to blue. The empty circles represent the best strategies for the highest RRR of each target. For minimizing infections, hospitalizations or ICU admissions, the higher prioritization is associated with the higher RRR. For minimizing deaths, the highest level of prioritization leads to a negative impact and medium levels of prioritization correspond to the highest RRR. The y-axis shows the population fractions (%) of three groups (*Minus, Neutral*, and *Plus*) for each shift in municipality priority (∆*n*). The geographic distribution of municipality priority (∆*n*) can be found in Fig 2.

**Table S14.**
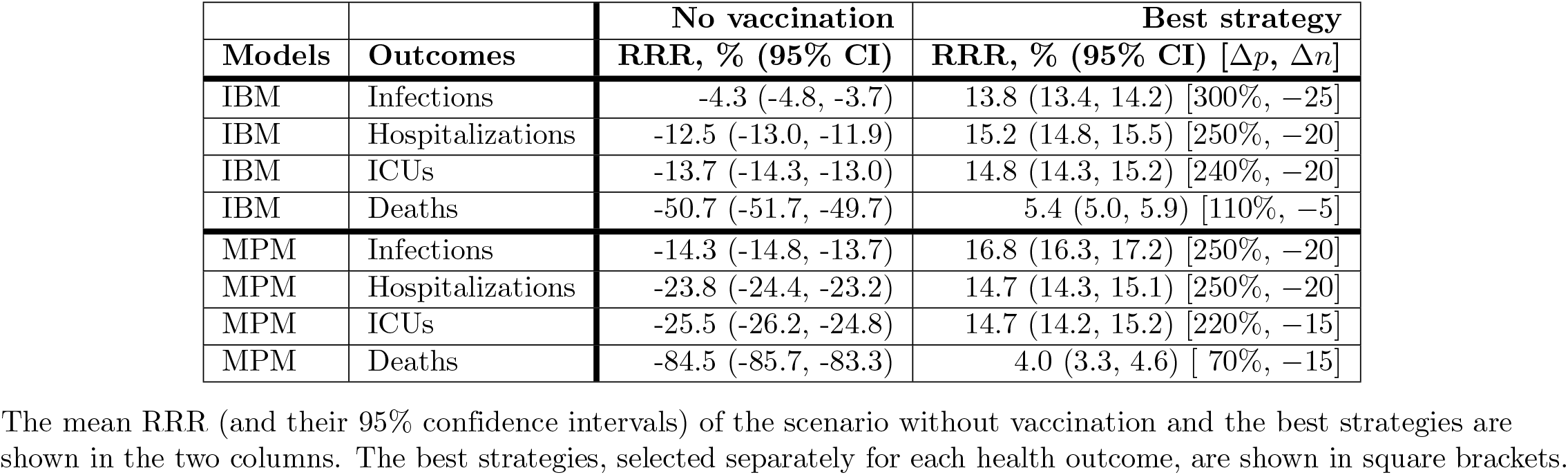
The relative risk reduction (RRR) of different strategies compared to the baseline strategy without geographic prioritization, given doubled hospitalization probabilities.

|  |  | No vaccination | Best strategy |
| --- | --- | --- | --- |
| Models | Outcomes | RRR, % (95% CI) | RRR, % (95% CI) [$\Delta p$, $\Delta n$] |
| IBM | Infections | -4.3 (-4.8, -3.7) | 13.8 (13.4, 14.2) [300%, -25] |
| IBM | Hospitalizations | -12.5 (-13.0, -11.9) | 15.2 (14.8, 15.5) [250%, -20] |
| IBM | ICUs | -13.7 (-14.3, -13.0) | 14.8 (14.3, 15.2) [240%, -20] |
| IBM | Deaths | -50.7 (-51.7, -49.7) | 5.4 (5.0, 5.9) [110%, -5] |
| MPM | Infections | -14.3 (-14.8, -13.7) | 16.8 (16.3, 17.2) [250%, -20] |
| MPM | Hospitalizations | -23.8 (-24.4, -23.2) | 14.7 (14.3, 15.1) [250%, -20] |
| MPM | ICUs | -25.5 (-26.2, -24.8) | 14.7 (14.2, 15.2) [220%, -15] |
| MPM | Deaths | -84.5 (-85.7, -83.3) | 4.0 (3.3, 4.6) [70%, -15] |

The mean RRR (and their 95% confidence intervals) of the scenario without vaccination and the best strategies are shown in the two columns. The best strategies, selected separately for each health outcome, are shown in square brackets.

The relative risk reduction (RRR) of alternative strategies were also calculated accordingly. Fig S24 shows the results from the IBM and MPM in alternative strategies of geographic prioritization. Table S14 shows all the RRR of the scenario without vaccination and the best strategies from two models. All the RRR are similar to the original analysis.

## References

1. Li Q, Guan X, Wu P, Wang X, Zhou L, Tong Y, et al. Early Transmission Dynamics in Wuhan, China, of Novel Coronavirus–Infected Pneumonia. New England Journal of Medicine. 2020;382(13):1199–1207. doi:10.1056/NEJMoa2001316.

2. Chan LYH, Yuan B, Convertino M. COVID-19 non-pharmaceutical intervention portfolio effectiveness and risk communication predominance. Scientific reports. 2021;11(1):1–17.

3. One person has tested positive for coronavirus, Norwegian Institute of Public Health, Norway;. Available from: https://www.fhi.no/en/archive/covid-19-archive/COVID-19-archived-news-from-2020/february/one-person-has-tested-positive-for-coronavirus/ [cited 19th October 2021].

4. Norwegian Directorate of Health implements the following today - from 6 p.m. on Thursday 12 March until Thursday 26 March 2020, Norwegian Institute of Public Health, Norway;. Available from: https://www.fhi.no/en/archive/covid-19-archive/COVID-19-archived-news-from-2020/march/norwegian-directorate-of-health-implements-the-following-today---from-6-p.m/ [cited 19th October 2021].

5. Social distance and physical contact, Norwegian Institute of Public Health, Norway;. Available from: https://www.fhi.no/en/archive/covid-19-archive/covid-19-guidance-archived-articles/social-distance-and-fewer-contacts/ [cited 19th October 2021].

6. Information about the use of face masks in other languages, Norwegian Institute of Public Health, Norway;. Available from: https://www.fhi.no/en/archive/covid-19-archive/covid-19-guidance-archived-articles/informasjon-om-bruk-av-munnbind-pa-flere-sprak/ [cited 31st October 2022].

7. 6 January: The City of Oslo extends the social lockdown of Oslo until 21st January, City of Oslo;. Available from: https://www.oslo.kommune.no/english/coronavirus/status-reports-on-coronavirus-measures/6-january-the-city-of-oslo-extends-the-social-lockdown-of-oslo-until-21st-january [cited 31st October 2022].

8. Saunes IS, Vrangbæk K, Byrkjeflot H, Jervelund SS, Birk HO, Tynkkynen LK, et al. Nordic responses to Covid-19: Governance and policy measures in the early phases of the pandemic. Health Policy. 2022;126(5):418–426. doi:https://doi.org/10.1016/j.healthpol.2021.08.011.

9. Coronavirus vaccination: Recommendation to prioritise vaccine groups based on infection situation, Norwegian Institute of Public Health, Norway;. Available from: https://www.fhi.no/en/news/2020/anbefaler-a-prioritere-vaksinegruppene-ut-fra-smittesituasjonen/ [cited 19th October 2021].

10. Who will get the coronavirus vaccine, Norwegian Institute of Public Health, Norway;. Available from: https://www.fhi.no/en/id/vaccines/coronavirus-immunisation-programme/who-will-get-coronavirus-vaccine-first/ [cited 19th October 2021].

11. Advice on priority groups for coronavirus vaccination in Norway, Norwegian Institute of Public Health, Norway;. Available from: https://www.fhi.no/contentassets/9d23593d6ebe443ba12556d3f7284eb8/norwegian-ethics-advisory-report-for-corona-vaccination.pdf [cited 19th October 2021].

12. 18-year-olds should be vaccinated with 44-year-olds, recommends NIPH, Norwegian Institute of Public Health, Norway;. Available from: https://www.fhi.no/en/news/2021/18-year-olds-should-be-vaccinated-with-44--year-olds-recommend-niph/ [cited 19th October 2021].

13. Changes in the vaccine strategy, Norwegian Institute of Public Health, Norway;. Available from: https://www.fhi.no/en/archive/covid-19-archive/covid-19---archived-news-2021/march/changes-in-the-vaccine-strategy/ [cited 19th October 2021].

14. Regjeringen har besluttet en geografisk omfordeling av vaksiner, Helse-og omsorgsdepartementet, Norway;. Available from: https://www.regjeringen.no/no/aktuelt/regjeringen-har-besluttet-en-geografisk-omfordeling-av-vaksiner/id2850061/ [cited 19th October 2021].

15. Coronavirus immunisation programme, Norwegian Institute of Public Health, Norway;. Available from: https://www.fhi.no/en/id/vaccines/coronavirus-immunisation-programme/ [cited 19th October 2021].

16. Bubar KM, Reinholt K, Kissler SM, Lipsitch M, Cobey S, Grad YH, et al. Model-informed COVID-19 vaccine prioritization strategies by age and serostatus. Science. 2021;371(6532):916–921. doi:10.1126/science.abe6959.

17. Buckner JH, Chowell G, Springborn MR. Dynamic prioritization of COVID-19 vaccines when social distancing is limited for essential workers. Proceedings of the National Academy of Sciences. 2021;118(16):e2025786118. doi:10.1073/pnas.2025786118.

18. Chen J, Hoops S, Marathe A, Mortveit H, Lewis B, Venkatramanan S, et al. Prioritizing allocation of COVID-19 vaccines based on social contacts increases vaccination effectiveness. medRxiv. 2021;doi:10.1101/2021.02.04.21251012.

19. Choi Y, Kim JS, Kim JE, Choi H, Lee CH. Vaccination Prioritization Strategies for COVID-19 in Korea: A Mathematical Modeling Approach. International Journal of Environmental Research and Public Health. 2021;18(8). doi:10.3390/ijerph18084240.

20. Goldenbogen B, Adler SO, Bodeit O, Wodke JAH, Escalera-Fanjul X, Korman A, et al. Control of COVID-19 Outbreaks under Stochastic Community Dynamics, Bimodality, or Limited Vaccination. Advanced Science. 2022;9(23):2200088. doi:https://doi.org/10.1002/advs.202200088.

21. Hjorleifsson KE, Rognvaldsson S, Jonsson H, Agustsdottir AB, Andresdottir M, Birgisdottir K, et al. Reconstruction of a large-scale outbreak of SARS-CoV-2 infection in Iceland informs vaccination strategies. Clinical Microbiology and Infection. 2022;28(6):852–858. doi:https://doi.org/10.1016/j.cmi.2022.02.012.

22. Li R, Bjørnstad ON, Stenseth NC. Prioritizing vaccination by age and social activity to advance societal health benefits in Norway: a modelling study. The Lancet Regional Health - Europe. 2021;10:100200. doi:https://doi.org/10.1016/j.lanepe.2021.100200.

23. Li R, Bjørnstad ON, Stenseth NC. Switching vaccination among target groups to achieve improved long-lasting benefits. Royal Society Open Science. 2021;8(6):210292. doi:10.1098/rsos.210292.

24. Matrajt L, Eaton J, Leung T, Brown ER. Vaccine optimization for COVID-19: Who to vaccinate first? Science Advances. 2021;7(6):eabf1374. doi:10.1126/sciadv.abf1374.

25. Moore S, Hill EM, Dyson L, Tildesley MJ, Keeling MJ. Modelling optimal vaccination strategy for SARS-CoV-2 in the UK. PLOS Computational Biology. 2021;17(5):1–20. doi:10.1371/journal.pcbi.1008849.

26. Sjödin H, Rocklöv J, Britton T. Evaluating and optimizing COVID-19 vaccination policies: a case study of Sweden. medRxiv. 2021;doi:10.1101/2021.04.07.21255026.

27. Voigt A, Omholt S, Almaas E. Comparing the impact of vaccination strategies on the spread of COVID-19, including a novel household-targeted vaccination strategy. PLOS ONE. 2022;17(2):1–16. doi:10.1371/journal.pone.0263155.

28. Bertsimas D, Digalakis Jr V, Jacquillat A, Li ML, Previero A. Where to locate COVID-19 mass vaccination facilities? Naval Research Logistics (NRL). 2022;69(2):179–200. doi:https://doi.org/10.1002/nav.22007.

29. Lemaitre JC, Pasetto D, Zanon M, Bertuzzo E, Mari L, Miccoli S, et al. Optimal control of the spatial allocation of COVID-19 vaccines: Italy as a case study. PLOS Computational Biology. 2022;18(7):1–20. doi:10.1371/journal.pcbi.1010237.

30. Molla J, Ponce de Léon Chávez A, Hiraoka T, Ala-Nissila T, Kivelä M, Leskelä L. Adaptive and optimized COVID-19 vaccination strategies across geographical regions and age groups. PLOS Computational Biology. 2022;18(4):1–19. doi:10.1371/journal.pcbi.1009974.

31. Grauer J, Löwen H, Liebchen B. Strategic spatiotemporal vaccine distribution increases the survival rate in an infectious disease like Covid-19. Scientific reports. 2020;10(1):1–10.

32. Svar på oppdrag 8 Vaksinasjon - Delleveranse reviderte anbefalinger for geografisk prioritering, Norwegian Institute of Public Health, Norway;. Available from: https://www.fhi.no/contentassets/1af4c6e655014a738055c79b72396de8/svar-pa-oppdrag-8-vaksinasjon---delleveranse-reviderte-anbefalinger-for-geografisk-prioritering.pdf [cited 15th December 2022].

33. Modelleringsrapport, delleveranse Oppdrag 8: Effekt av regional prioritering av covid-19 vaksiner til Oslo eller Oslo-Viken samt vaksinenes effekt på transmisjon for epidemiens videre utvikling (preliminære resultater), Norwegian Institute of Public Health, Norway;. Available from: https://www.fhi.no/contentassets/1af4c6e655014a738055c79b72396de8/modelleringsrapport_delleveranse_oppdrag8_2402.pdf [cited 15th December 2022].

34. Tilleggsanalyser til modelleringsrapport, delleveranse Oppdrag 8: Effekt av moderat regional prioritering av covid-19 vaksiner til utvalgte kommuner – 5. Mars 2021, Norwegian Institute of Public Health, Norway;. Available from: https://www.fhi.no/contentassets/1af4c6e655014a738055c79b72396de8/vedlegg-5-til-delleveranse-8.pdf [cited 15th December 2022].

35. Modelleringsrapport til Oppdrag 8, Norwegian Institute of Public Health, Norway;. Available from: https://www.fhi.no/contentassets/e6b5660fc35740c8bb2a32bfe0cc45d1/vedlegg/nasjonale-og-regionale-rapporter/oppdrag_8_2303_bfdblasio.pdf [cited 15th December 2022].

36. Emergency preparedness register for COVID-19 (Beredt C19), Norwegian Institute of Public Health, Norway;. Available from: https://www.fhi.no/en/id/infectious-diseases/coronavirus/emergency-preparedness-register-for-covid-19/ [cited 24th January 2023].

37. Norwegian Surveillance System for Communicable Diseases (MSIS), Norwegian Institute of Public Health, Norway;. Available from: https://www.fhi.no/en/hn/health-registries/msis/ [cited 31st October 2022].

38. Norsk pandemiregister, Helse Bergen, Norway;. Available from: https://helse-bergen.no/norsk-pandemiregister [cited 2nd February 2023].

39. Norwegian Intensive Care and Pandemic Registry (NIPaR), Helsedata, Norway;. Available from: https://helsedata.no/en/forvaltere/bergen-hospital-trust/norwegian-intensive-care-and-pandemic-registry/ [cited 2nd February 2023].

40. Norwegian Immunisation Registry SYSVAK, Norwegian Institute of Public Health, Norway;. Available from: https://www.fhi.no/en/hn/health-registries/norwegian-immunisation-registry-sysvak/ [cited 19th October 2021].

41. Norwegian Immunisation Registry (SYSVAK), Helsedata, Norway;. Available from: https://helsedata.no/en/forvaltere/norwegian-institute-of-public-health/norwegian-immunisation-registry-sysvak/ [cited 25th January 2023].

42. Coronavirus vaccine - information for the public, Norwegian Institute of Public Health, Norway;. Available from: https://www.fhi.no/en/id/vaccines/coronavirus-immunisation-programme/coronavirus-vaccine/#risk-groups [cited 12th April 2023].

43. Engebretsen S, Diz-Lois Palomares A, Rø G, Kristoffersen AB, Lindstrøm JC, Engø-Monsen K, et al. A real-time regional model for COVID-19: Probabilistic situational awareness and forecasting. PLOS Computational Biology. 2023;19(1):1–26. doi:10.1371/journal.pcbi.1010860.

44. Telenor;. Available from: https://www.telenor.com [cited 24th February 2023].

45. Di Ruscio F, Guzzetta G, Bjørnholt JV, Leegaard TM, Moen AEF, Merler S, et al. Quantifying the transmission dynamics of MRSA in the community and healthcare settings in a low-prevalence country. Proceedings of the National Academy of Sciences. 2019;116(29):14599–14605. doi:10.1073/pnas.1900959116.

46. R Core Team. R: A Language and Environment for Statistical Computing; 2022. Available from: https://www.R-project.org/.

47. FitzJohn R, Fischer T. odin: ODE Generation and Integration; 2022. Available from: https://github.com/mrc-ide/odin.

48. FitzJohn R, Baguelin M, Knock E, Whittles L, Lees J, Sonabend R. mcstate: Monte Carlo Methods for State Space Models; 2022. Available from: https://github.com/mrc-ide/mcstate.

49. Herrera-Esposito D, de Los Campos G. Age-specific rate of severe and critical SARS-CoV-2 infections estimated with multi-country seroprevalence studies. BMC infectious diseases. 2022;22(1):1–14.

50. Situational awareness and forecasting for Norway on 20th January 2021, Norwegian Institute of Public Health, Norway;. Available from: https://www.fhi.no/contentassets/e6b5660fc35740c8bb2a32bfe0cc45d1/vedlegg/nasjonale-og-regionale-rapporter/2021.01.20-national-and-regional-corona-report-engelsk.pdf [cited 12th December 2022].

51. Coronavirus modelling at the NIPH, Norwegian Institute of Public Health, Norway;. Available from: https://www.fhi.no/en/id/infectious-diseases/coronavirus/coronavirus-modelling-at-the-niph-fhi/ [cited 19th October 2021].

52. Thompson MG, Burgess JL, Naleway AL, Tyner H, Yoon SK, Meece J, et al. Prevention and Attenuation of Covid-19 with the BNT162b2 and mRNA-1273 Vaccines. New England Journal of Medicine. 2021;385(4):320–329. doi:10.1056/NEJMoa2107058.

53. Dagan N, Barda N, Kepten E, Miron O, Perchik S, Katz MA, et al. BNT162b2 mRNA Covid-19 Vaccine in a Nationwide Mass Vaccination Setting. New England Journal of Medicine. 2021;384(15):1412–1423. doi:10.1056/NEJMoa2101765.

54. Vaccination scenario, Norwegian Institute of Public Health, Norway;. Available from: https://www.fhi.no/en/publ/posters/vaksineringsscenario/ [cited 19th October 2021].

55. Tunheim G, Rø G[U+FFFD], Chopra A, Aase A, Kran AMB, Vaage JT, et al. Prevalence of antibodies against SARS-CoV-2 in the Norwegian population, August 2021. Influenza and Other Respiratory Viruses. 2022;16(6):1004–1013. doi:https://doi.org/10.1111/irv.13024.

56. Veneti L, Seppälä E, Larsdatter Storm M, Valcarcel Salamanca B, Alnes Buanes E, Aasand N, et al. Increased risk of hospitalisation and intensive care admission associated with reported cases of SARS-CoV-2 variants B.1.1.7 and B.1.351 in Norway, December 2020 –May 2021. PLOS ONE. 2021;16(10):1–12. doi:10.1371/journal.pone.0258513.

57. James LP, Salomon JA, Buckee CO, Menzies NA. The Use and Misuse of Mathematical Modeling for Infectious Disease Policymaking: Lessons for the COVID-19 Pandemic. Medical Decision Making. 2021;41(4):379–385. doi:10.1177/0272989X21990391.

58. Monod M, Blenkinsop A, Xi X, Hebert D, Bershan S, Tietze S, et al. Age groups that sustain resurging COVID-19 epidemics in the United States. Science. 2021;371(6536):eabe8372. doi:10.1126/science.abe8372.

59. COVID-19 Ukerapport - uke 20, Norwegian Institute of Public Health, Norway;. Available from: https://www.fhi.no/contentassets/8a971e7b0a3c4a06bdbf381ab52e6157/vedlegg/3.-alle-ukerapporter-2021/ukerapport-uke-20-17.05---23.05.21.pdf [cited 14th April 2023].

60. Advice on priority groups for coronavirus vaccination in Norway (Ethics report), Norwegian Institute of Public Health, Norway;. Available from: https://www.fhi.no/en/publ/2020/advice-on-priority-groups-for-coronavirus-vaccination-in-norway/ [cited 27th July 2023].

61. Prioriteringskriteriene ved offentlig finansiering av legemidler, Statens Legemiddelverket, Norway;. Available from: https://legemiddelverket.no/offentlig-finansiering/slik-far-legemidler-offentlig-finansiering/hva-inneberer-prioriteringskriteriene-ressursbruk-nytte-og-alvorlighet [cited 11th July 2023].

62. Amdam H, Norheim OF, Solberg CT, Littmann JR. Can Geographically Targeted Vaccinations Be Ethically Justified? The Case of Norway During the COVID-19 Pandemic. Public Health Ethics. 2023; p. phad011. doi:10.1093/phe/phad011.

63. Ajelli M, Goncalves B, Balcan D, Colizza V, Hu H, Ramasco JJ, et al. Comparing large-scale computational approaches to epidemic modeling: agent-based versus structured metapopulation models. BMC infectious diseases. 2010;10(1):1–13.

64. Zachreson C, Chang S, Harding N, Prokopenko M. The effects of local homogeneity assumptions in metapopulation models of infectious disease. Royal Society Open Science. 2022;9(7):211919. doi:10.1098/rsos.211919.

65. Folkehelseinstituttets foreløpige anbefalinger om vaksinasjon mot covid-19 og om prioritering av covid-19-vaksiner, Norwegian Institute of Public Health, Norway;. Available from: https://www.fhi.no/contentassets/d07db6f2c8f74fa586e2d2a4ab24dfdf/forelopige-anbefalinger-og-prioriteringer-1-utgave-00017622.pdf [cited 15th December 2022].

66. Folkehelseinstituttets foreløpige anbefalinger om vaksinasjon mot covid-19 og om prioritering av covid-19-vaksiner, versjon 2, Norwegian Institute of Public Health, Norway;. Available from: https://www.fhi.no/contentassets/d07db6f2c8f74fa586e2d2a4ab24dfdf/2020-12-v2-anbefalinger-og-prioriteringer-2-utgave-korrigert-forside.pdf [cited 15th December 2022].

67. Modelleringsrapport til Oppdrag 346, Norwegian Institute of Public Health, Norway;. Available from: https://www.fhi.no/contentassets/e6b5660fc35740c8bb2a32bfe0cc45d1/vedlegg/nasjonale-og-regionale-rapporter/2021-04-07-modelleringsrapport-til-oppdrag-346.pdf [cited 15th December 2022].

68. Svar på oppdrag 21 - Om gjenåpning og vaksiner, Norwegian Institute of Public Health, Norway;. Available from: https://www.fhi.no/contentassets/3596efb4a1064c9f9c7c9e3f68ec481f/svar-pa-oppdrag-21_sladdet.pdf [cited 15th December 2022].

69. Delsvar på revidert oppdrag 16. Nye vurderinger av vaksinasjonsstrategien, Norwegian Institute of Public Health, Norway;. Available from: https://www.fhi.no/contentassets/3596efb4a1064c9f9c7c9e3f68ec481f/delsvar-pa-oppdrag-16_geografisk-malrettet-fordeling-2020-04-26.pdf [cited 15th December 2022].

70. Nye vurderinger av vaksinasjonsstrategien, Norwegian Institute of Public Health, Norway;. Available from: https://www.fhi.no/contentassets/3596efb4a1064c9f9c7c9e3f68ec481f/svar-pa-oppdrag-16---nye-vurderinger-av-vaksinasjonsstrategien-inkl.-vedlegg_skjult-innhold.pdf[cited 15th December 2022].

71. Modellering av ulike vaksinasjonsscenario med og uten virusvektorvaksiner, Norwegian Institute of Public Health, Norway;. Available from: https://www.regjeringen.no/contentassets/46795a8782994e368e7a6a57d56bdf95/rapport-vorland-utvalg.pdf [cited 15th December 2022].

72. Oppdatert svar på covid-19 oppdrag fra HOD 473 – Revisjon av strategi og beredskapsplan for covid-19, Norwegian Institute of Public Health, Norway;. Available from: https://www.helsedirektoratet.no/tema/beredskap-og-krisehandtering/koronavirus/faglig-grunnlag-til-helse-og-omsorgsdepartementet-covid-19/Oppdatert%20svar%20p%C3%A5%20covid-19%20oppdrag%20fra%20HOD%20473%20%E2%80%93%20Revisjon%20av%20strategi%20og%20beredskapsplan%20for%20covid-19%20med%20vedlegg.pdf/_/attachment/inline/a5984135-8e00-4ae1-ae4c-84a63027d2e6:15480d5b07fc545ddcde199834e21a6ea651e190/Oppdatert%20svar%20p%C3%A5%20covid-19%20oppdrag%20fra%20HOD%20473%20%E2%80%93%20Revisjon%20av%20strategi%20og%20beredskapsplan%20for%20covid-19%20med%20vedlegg.pdf [cited 15th December 2022].

73. Svar på oppdrag 37 - Koronavaksinasjon av ungdom, Norwegian Institute of Public Health, Norway;. Available from: https://www.fhi.no/contentassets/3596efb4a1064c9f9c7c9e3f68ec481f/2021-07-05-oppdrag-37_vaksinasjon-av-16-17-aringer.pdf [cited 15th December 2022].

74. Modelleringsrapport til Oppdrag 45, 26. august 2021, Norwegian Institute of Public Health, Norway;. Available from: https://www.fhi.no/contentassets/3596efb4a1064c9f9c7c9e3f68ec481f/2021-09-02-oppdrag-45_ vedlegg-2_modelleringsrapport_rettet.pdf [cited 15th December 2022].

75. Revidert svar på covid-19 oppdrag fra HOD 530 – Samleoppdrag om innreiserestriksjoner, reiseråd, innreisekarantene, karantenehotell, bruk av koronasertifikat og testing ved innreise, Norwegian Institute of Public Health, Norway;. Available from: https://www.helsedirektoratet.no/tema/beredskap-og-krisehandtering/koronavirus/faglig-grunnlag-til-helse-og-omsorgsdepartementet-covid-19/Revidert%20svar%20p%C3%A5%20covid-19%20oppdrag%20fra%20HOD%20530%20%E2%80%93%20Samleoppdrag%20om%20innreiserestriksjoner,%20reiser%C3%A5d,%20innreisekarantene,%20karantenehotell,%20bruk%20av%20koronasertifikat%20og%20testing.pdf/_/attachment/inline/aeefa603-6e64-4e89-a016-795e7bed8e09:3da708d3d7ceb6ff90a36ed513628499109352e2/Revidert%20svar%20p%C3%A5%20covid-19%20oppdrag%20fra%20HOD%20530%20%E2%80%93%20Samleoppdrag%20om%20innreiserestriksjoner,%20reiser%C3%A5d,%20innreisekarantene,%20karantenehotell,%20bruk%20av%20koronasertifikat%20og%20testing.pdf [cited 15th December 2022].

76. Svar på Oppdrag 58 – Om dose to til 12-15 åringer, Norwegian Institute of Public Health, Norway;. Available from: https://www.fhi.no/contentassets/3596efb4a1064c9f9c7c9e3f68ec481f/2022-01-07-svar-pa-oppdrag-58-om-dose-to-til-12-15-aringer.pdf [cited 15th December 2022].

77. Merow C, Urban MC. Seasonality and uncertainty in global COVID-19 growth rates. Proceedings of the National Academy of Sciences. 2020;117(44):27456–27464. doi:10.1073/pnas.2008590117.

78. Kamineni M, Engø-Monsen K, Midtbø JE, Forland F, de Blasio BF, Frigessi A, et al. Effects of non-compulsory and mandatory COVID-19 interventions on travel distance and time away from home, Norway, 2021. Eurosurveillance. 2023;28(17). doi:https://doi.org/10.2807/1560-7917.ES.2023.28.17.2200382.

79. Davies NG, Klepac P, Liu Y, Prem K, Jit M, Eggo RM. Age-dependent effects in the transmission and control of COVID-19 epidemics. Nature medicine. 2020;26(8):1205–1211.

80. Rozhnova G, van Dorp CH, Bruijning-Verhagen P, Bootsma MC, van de Wijgert JH, Bonten MJ, et al. Model-based evaluation of school-and non-school-related measures to control the COVID-19 pandemic. Nature communications. 2021;12(1):1–11.

81. Viana J, van Dorp CH, Nunes A, Gomes MC, van Boven M, Kretzschmar ME, et al. Controlling the pandemic during the SARS-CoV-2 vaccination rollout. Nature communications. 2021;12(1):1–15.

82. Jing QL, Liu MJ, Zhang ZB, Fang LQ, Yuan J, Zhang AR, et al. Household secondary attack rate of COVID-19 and associated determinants in Guangzhou, China: a retrospective cohort study. The Lancet Infectious Diseases. 2020;20(10):1141–1150. doi:https://doi.org/10.1016/S1473-3099(20)30471-0.

83. Goldstein E, Lipsitch M, Cevik M. On the Effect of Age on the Transmission of SARS-CoV-2 in Households, Schools, and the Community. The Journal of Infectious Diseases. 2020;223(3):362–369. doi:10.1093/infdis/jiaa691.

84. Maier BF, Burdinski A, Rose AH, Schlosser F, Hinrichs D, Betsch C, et al. Potential benefits of delaying the second mRNA COVID-19 vaccine dose; 2021. Available from: https://arxiv.org/abs/2102.13600.

85. Matrajt L, Eaton J, Leung T, Dimitrov D, Schiffer JT, Swan DA, et al. Optimizing vaccine allocation for COVID-19 vaccines shows the potential role of single-dose vaccination. Nature communications. 2021;12(1):1–18.

86. Saad-Roy CM, Morris SE, Metcalf CJE, Mina MJ, Baker RE, Farrar J, et al. Epidemiological and evolutionary considerations of SARS-CoV-2 vaccine dosing regimes. Science. 2021;372(6540):363–370. doi:10.1126/science.abg8663.

87. Classification of statistical tract and basic statistical unit, Statistics Norway, Norway;. Available from: https://www.ssb.no/en/klass/klassifikasjoner/1 [cited 25th January 2023].

88. Hinch R, Probert WJM, Nurtay A, Kendall M, Wymant C, Hall M, et al. OpenABM-Covid19—An agent-based model for non-pharmaceutical interventions against COVID-19 including contact tracing. PLOS Computational Biology. 2021;17(7):1–26. doi:10.1371/journal.pcbi.1009146.

89. Rypdal M, Rypdal V, Jakobsen PK, Ytterstad E, Løvsletten O, Klingenberg C, et al. Modelling suggests limited change in the reproduction number from reopening Norwegian kindergartens and schools during the COVID-19 pandemic. PLOS ONE. 2021;16(2):1–13. doi:10.1371/journal.pone.0238268.

90. Hoertel N, Blachier M, Blanco C, Olfson M, Massetti M, Rico MS, et al. A stochastic agent-based model of the SARS-CoV-2 epidemic in France. Nature medicine. 2020;26(9):1417–1421.

91. Fumanelli L, Ajelli M, Manfredi P, Vespignani A, Merler S. Inferring the Structure of Social Contacts from Demographic Data in the Analysis of Infectious Diseases Spread. PLOS Computational Biology. 2012;8(9):1–10. doi:10.1371/journal.pcbi.1002673.

92. Statistics Norway;. Available from: https://www.ssb.no/en [cited 31st October 2022].

93. Andrews N, Tessier E, Stowe J, Gower C, Kirsebom F, Simmons R, et al. Duration of Protection against Mild and Severe Disease by Covid-19 Vaccines. New England Journal of Medicine. 2022;386(4):340–350. doi:10.1056/NEJMoa2115481.

94. Risiko ved covid-19-epidemien og ved omikronvarianten i Norge, Norwegian Institute of Public Health, Norway;. Available from: https://www.fhi.no/contentassets/c9e459cd7cc24991810a0d28d7803bd0/vedlegg/risikovurdering-2021-12-13.pdf [cited 19th April 2023].

95. Modelling scenarios for the SARS-CoV-2 Omicron VOC (B.1.1.529) in Norway during the winter 2021—2022, Norwegian Institute of Public Health, Norway;. Available from: https://www.fhi.no/contentassets/e6b5660fc35740c8bb2a32bfe0cc45d1/vedlegg/nasjonale-og-regionale-rapporter/omicron_modelling_report_2021_12_22.pdf [cited 19th April 2023].

96. Modelling scenarios for the SARS-CoV-2 Omicron VOC (B.1.1.529) in Norway, January-February 2022, Norwegian Institute of Public Health, Norway;. Available from: https://www.fhi.no/contentassets/e6b5660fc35740c8bb2a32bfe0cc45d1/vedlegg/nasjonale-og-regionale-rapporter/modelling-scenarios-for-the-sars-cov-2-omicron-voc-b.1.1.529-in-norway-january-february-2022.pdf [cited 19th April 2023].

97. Modelling scenarios for the SARS-CoV-2 Omicron VOC (B.1.1.529) in Norway, gradual reopening in February-March 2022, Norwegian Institute of Public Health, Norway;. Available from: https://www.fhi.no/contentassets/e6b5660fc35740c8bb2a32bfe0cc45d1/vedlegg/nasjonale-og-regionale-rapporter/modelling-scenarios-for-the-sars-cov-2-omicron-voc-26.01.2022.pdf [cited 19th April 2023].

98. Ferretti L, Wymant C, Kendall M, Zhao L, Nurtay A, Abeler-Dörner L, et al. Quantifying SARS-CoV-2 transmission suggests epidemic control with digital contact tracing. Science. 2020;368(6491):eabb6936. doi:10.1126/science.abb6936.

99. Carpenter B, Gelman A, Hoffman MD, Lee D, Goodrich B, Betancourt M, et al. Stan: A probabilistic programming language. Journal of statistical software. 2017;76(1).

100. Stan Development Team. RStan: the R interface to Stan; 2022. Available from: http://mc-stan.org/.

101. Grint DJ, Wing K, Houlihan C, Gibbs HP, Evans SJW, Williamson E, et al. Severity of Severe Acute Respiratory System Coronavirus 2 (SARS-CoV-2) Alpha Variant (B.1.1.7) in England. Clinical Infectious Diseases. 2021;75(1):e1120–e1127. doi:10.1093/cid/ciab754.

102. Pascall DJ, Vink E, Blacow R, Bulteel N, Campbell A, Campbell R, et al. The SARS-CoV-2 Alpha variant is associated with increased clinical severity of COVID-19 in Scotland: a genomics-based retrospective cohort analysis. MedRxiv. 2022; p. 2021–08.

103. Bager P, Wohlfahrt J, Fonager J, Rasmussen M, Albertsen M, Michaelsen TY, et al. Risk of hospitalisation associated with infection with SARS-CoV-2 lineage B.1.1.7 in Denmark: an observational cohort study. The Lancet Infectious Diseases. 2021;21(11):1507–1517. doi:https://doi.org/10.1016/S1473-3099(21)00290-5.

104. Funk T, Pharris A, Spiteri G, Bundle N, Melidou A, Carr M, et al. Characteristics of SARS-CoV-2 variants of concern B.1.1.7, B.1.351 or P.1: data from seven EU/EEA countries, weeks 38/2020 to 10/2021. Eurosurveillance. 2021;26(16). doi:https://doi.org/10.2807/1560-7917.ES.2021.26.16.2100348.

105. Tunheim G, Rø G[U+FFFD], Tran T, Kran AMB, Andersen JT, Vaage EB, et al. Trends in seroprevalence of SARS-CoV-2 and infection fatality rate in the Norwegian population through the first year of the COVID-19 pandemic. Influenza and Other Respiratory Viruses. 2022;16(2):204–212. doi:https://doi.org/10.1111/irv.12932.

106. Seroprevalence of SARS-CoV-2 in the Norwegian population measured in residual sera collected in January 2021, Norwegian Institute of Public Health, Norway;. Available from: https://www.fhi.no/en/publ/2021/seroprevalence-of-sars-cov-2-in-the-norwegian-population-measured-in-residu/ [cited 10th October 2022].

107. Seroprevalence of SARS-CoV-2 in the Norwegian population measured in residual sera collected in April/May 2020 and August 2019, Norwegian Institute of Public Health, Norway;. Available from: https://www.fhi.no/en/publ/2020/seroprevalence-of-sars-cov-2-in-the-norwegian-population--measured-in-resid/ [cited 10th October 2022].

108. Anda EE, Braaten T, Borch KB, Nøst TH, Chen SL, Lukic M, et al. Seroprevalence of antibodies against SARS-CoV-2 virus in the adult Norwegian population, winter 2020/2021: pre-vaccination period. medRxiv. 2021;.

109. Cori A, Ferguson NM, Fraser C, Cauchemez S. A New Framework and Software to Estimate Time-Varying Reproduction Numbers During Epidemics. American Journal of Epidemiology. 2013;178(9):1505–1512. doi:10.1093/aje/kwt133.

110. Fraser C. Estimating Individual and Household Reproduction Numbers in an Emerging Epidemic. PLOS ONE. 2007;2(8):1–12. doi:10.1371/journal.pone.0000758.

111. Wallinga J, Teunis P. Different Epidemic Curves for Severe Acute Respiratory Syndrome Reveal Similar Impacts of Control Measures. American Journal of Epidemiology. 2004;160(6):509–516. doi:10.1093/aje/kwh255.

